# DISCRIMINATOR: Assigning Cohort-Wide Provisional Pathogenicity Classifications to CNVs

**DOI:** 10.1101/2022.09.26.22278681

**Authors:** Alyssa S. Wetzel, Heather Major, Mrutyunjaya Parida, J. Robert Manak, Benjamin W. Darbro

## Abstract

The interpretation of clinical chromosomal microarrays (CMAs) has historically relied on the relevance of identified copy number variants (CNVs) to the clinical phenotype. New interpretation guidelines are focused on standardizing pathogenicity classifications based on genomic location, gene content, and previous publications, rather than the immediate clinical relevance. Here we report on DISCRIMINATOR, which was developed to assign provisional pathogenicity classifications based on genomic location by integrating information on putative benign and pathogenic loci in the human genome. However, its application extends beyond that of a simple classifier. The novel utility of DISCRIMINATOR is its ability to operate on a cohort-level and easily integrate updated definitions of benign and pathogenic regions of the human genome. We used DISCRIMINATOR to assign provisional pathogenicity classifications (‘Benign’, ‘Secondary’, ‘Primary’ or ‘Non-Coding’) to 87,808 CNVs in 3,362 cases ascertained through clinical CMA testing. The majority of identified CNVs were provisionally classified as ‘Benign’ or ‘Non-Coding’ and consistent with their prevalence rates, 15q11.2, 16p11.2, and 22q11.2 were the most common ‘Primary’ CNVs detected. Targeted re-analysis led to the identification of several cases where DISCRIMINATOR identified a ‘Primary’ CNV within a case that had a non-Abnormal CMA test result, and several cases where only benign and/or non-coding CNVs were identified in reports with a ‘VUS’ CMA test result. Together these results show the utility of large-scale re-analysis of CMA data and how DISCRIMINATOR addresses this long-standing challenge.

## Introduction

Clinical chromosomal microarray (CMA) is a first-tier diagnostic test in the evaluation of patients with non-syndromic developmental delay, intellectual disability, autism spectrum disorders and multiple congenital anomalies not specific to a well-delineated genetic syndrome ^2–7^. CMA has allowed for the identification of the genetic cause for many distinct clinical syndromes, and recurrent copy number variants (CNVs) have been used to establish novel clinically recognizable syndromes ^8–16^. However, these recurrent pathogenic CNVs represent a subset of the CNVs present in the human genome. To date, over 500,000 unique CNVs have been found in healthy individuals ^17^, and the Clinical Genome Resource (ClinGen) has classified nearly 11,000 CNVs as VUS (variants of uncertain significance), and an additional 5,000 CNVs as *likely* pathogenic or *likely* benign ^18–20^. Despite the ever-increasing numbers of VUS, and attempts to develop interpretative guidelines, there is limited consistency in how these CNVs are classified and reported ^21–25^.

Recurrent benign and pathogenic CNVs are more straightforward to classify and interpret, as by definition, both have been observed previously in either control or affected patient populations, respectively. In addition, guidelines from professional organizations have proposed criteria that allow a CNV to be interpreted as pathogenic: multiple independent reports of similarly sized CNVs in patients with overlapping phenotypic features, an inheritance pattern within families that supports segregation of the CNV with affected status, and gene-phenotype correlations ^21–25^. In contrast, individual VUS CNVs are so rare that they may have only ever been seen in a very small handful of individuals or even just one family. As such, clinical interpretation of a VUS to a given patient is a manual and subjective process which relies heavily on the genes present within the CNV, what those genes are known or believed to do, and the phenotype of the patient. While expert consensus criteria for benign and pathogenic CNVs help to prevent against insufficiently characterized CNVs from being called pathogenic, or CNVs with no clinical relevance from being reported clinically, this leaves a majority of non-recurrent and/or rare CNVs classified as VUS ^7,26–28^, or with conflicting pathogenicity classifications ^21,26^. Recently, to reduce the amount of discordance in CNV pathogenicity classifications, a joint ACMGG and ClinGen working group created a series of recommendations for the classification of CNVs, namely by uncoupling the immediate clinical relevance from the pathogenicity classification of the variant ^24,25^. However, it remains to be seen whether the new pathogenicity guidelines will significantly shift our ability to classify an otherwise VUS CNV.

Here we report on the development and performance of DISCRIMINATOR, a computational program that was created to address the limitations of current methods in non-phenotype driven discrimination of pathogenic, benign, and VUS CNVs. Namely, this program assigns provisional pathogenicity classifications (benign, primary, secondary, or non-coding) to sample CNVs. These provisional classifications were created to guide interpretation rather than directly attribute pathogenicity, given that expert consensus relies on more than just genomic location in the classification of CNV pathogenicity. In addition to its function as a CNV classifier, DISCRIMINATOR uniquely provides summary metrics on the number of primary, secondary, benign, and non-coding CNVs identified within a given dataset, and annotates each primary CNV with the associated known microdeletion and microduplication syndromes (MMS), and all coding CNVs with the affected genes. Within the clinical context, DISCRIMINATOR can be utilized in real-time to prioritize CNVs identified by CMA for interpretation and/or to perform retrospective cohort analyses and identify CMAs for re-analysis and/or re-interpretation. In this paper, we report on three cohorts of CMA testing data obtained between 2011-2018 and provide descriptive metrics on the types of CNVs that were identified as well as the utility of DISCRIMINATOR in the reanalysis of these cohorts.

## Methods

### Case Cohort

Our study population consists of chromosomal microarray (CMA) tests performed at the University of Iowa’s Shivanand R. Patil Cytogenetics and Molecular Laboratory between 2011 and 2018 (*N* = *3524*). The Institutional Review Board of the University of Iowa has determined that this work does not meet the regulatory definition of human subjects research and does not require review. CMAs performed between 2011 and 2012 underwent testing with NimbleGen 385K (*n* = 415) or 720K (*n* = 665) feature comparative genomic hybridization (CGH) microarrays (Cohort #1), while CMAs performed between 2012 and 2018 underwent testing with the Affymetrix CytoscanHD copy number and single nucleotide polymorphism CGH microarrays (*n* = 2444). The Affymetrix CytoscanHD cases were further subdivided into two relatively equal-sized cohorts (Cohort #2 (2012-2015; n=1293); Cohort #3 (2015-2018; n=1158)) to investigate differences that might exist following the relocation of the testing lab to a new facility. CMA results were reported out as “Normal” if only presumed benign CNV(s) were detected, “Abnormal” if one or more known or presumed pathogenic CNV(s) were identified, or “VUS” (variant(s) of unknown significance) if one or more CNV(s) were detected that could not be classified as either benign or pathogenic. Finally, if the only detectable lesion was a region of homozygosity larger than 5 Mb and not observed in prior cases, the CMA result was reported out as “Region of Homozygosity” (ROH). All CNVs reported out on the clinical CMA report were recorded and broken down into two groups: CNVs which correspond to a known MMS (‘MMS CNVs’) and the remaining pathogenic, likely pathogenic, and VUS CNVs (‘non-MMS CNVs’; *e*.*g*. large *de novo* duplications or deletions).

### Microarray Data Curation

CMA quality was assessed using three metrics obtained from Nexus Copy Number software (Versions 4.0-9.0; BioDiscovery Inc): quality score, total number of CNVs, and ratio of copy number gains to losses as previously described ^29^ (**Supplemental Figure 1**). Arrays that scored four standard deviations from the cohort mean for any parameter were manually inspected to determine if the array was of poor quality (excluded) or skewed due to an autosomal aneuploidy (retained). Arrays containing whole chromosome or chromosomal arm aneuploidies of the X and Y chromosome were excluded from further analysis. Next, a two-standard deviation from the cohort mean cutoff was utilized to obtain the highest quality arrays. Arrays were removed from the cohort if (1) all three parameters exceeded the cutoff, (2) two parameters exceeded the cutoff including the quality score, and (3) two parameters exceeded the cutoff and manual inspection revealed subjectively noisy data. A total of 156 arrays (4.42%) were excluded for low quality (*n* = 89), duplications or deletion of the X-chromosome (*n* = 34), duplicate arrays (*n* = 30), or gender mismatch (*n* =3). For cases who had two microarrays run (duplicate arrays), the microarray with the newest and highest quality data was used.

We removed all CNVs that did not meet a minimum size threshold of 20kb. CNVs on the Y-chromosome or within the pseudo-autosomal regions of the X-chromosome were removed unless the CNVs were previously determined by clinical cytogenetics interpretation to be clinically relevant (*e*.*g*. CNVs involving the *SHOX* gene). UCSC’s Lift-Over tool ^30^ was used to convert CNV coordinates from the NimbleGen microarrays from hg18 to hg19. The Affymetrix microarrays were not gender matched CGH arrays, therefore, sex chromosome CNVs which occurred at a high frequency in females (>5%; X-chromosome) and males (>1%; X-chromosome) were excluded. Globally, individual CNV calls from a single sample were merged with respect to direction if they were within 30kb of each other. To account for the segmental duplication content of the 1q21.1, 15q11.2, and 22q11.2 MMS regions and ensure that large deletions and duplications across these regions were not segmented, the merging criteria for these specific regions was relaxed (**Supplementary Methods**). CNVs in these regions were merged if the region separating two CNV calls was less than 100kb, predominantly overlapped by a segmental duplication region (>90%), or located in a 500kb region surrounding the 22q11.2 segmental duplication at breakpoint “B” (chr22:20249559-20744436). The final CNV dataset consisted of 87,808 CNVs (**Supplementary Figure 1**).

### Overview of DISCRIMINATOR

DISCRIMINATOR is a CNV classifier which utilizes user-supplied benign and pathogenic intervals to assign provisional pathogenicity classifications to the input/sample CNV(s) in a stepwise manner: primary, benign, or secondary, respectively (**Figure 1; Supplemental Figure 2**). Provisional classifications were created as additional information beyond genomic location is required by expert consensus in the classification of CNV pathogenicity. Sample CNV(s) are classified as primary if the Jaccard similarity index score between the sample CNV and a pathogenic region exceeds a user-defined threshold (default set to 40%; see **Supplementary Methods**). If more than one primary CNV exceeds this threshold, DISCRIMINATOR will classify using the Primary CNV with the largest Jaccard value. Sample CNV(s) which do not overlap a known protein coding gene (see **Supplementary Methods**) are classified as non-coding. Coding CNVs are classified as benign if the percent overlap between the sample CNV and a benign region exceeds the 50% threshold. All remaining CNV(s) not classified as primary or benign are classified as secondary. All non-benign (*i*.*e*. primary and secondary) sample CNV(s) are then annotated with either the genes contained within the CNV interval or the genes whose exons are contained within the CNV interval. For our purposes, DISCRIMINATOR utilized the output of Benign-Ex (Wetzel *et al*.; manuscript in preparation ^31^) to define benign regions, and the manually curated list of MMS (Wetzel and Darbro; in press ^32^) to define pathogenic regions (see **Supplementary Methods**). However, DISCRIMINATOR has the flexibility to use any benign and/or pathogenic interval list provided they meet the formatting requirements.

**Figure 1:**
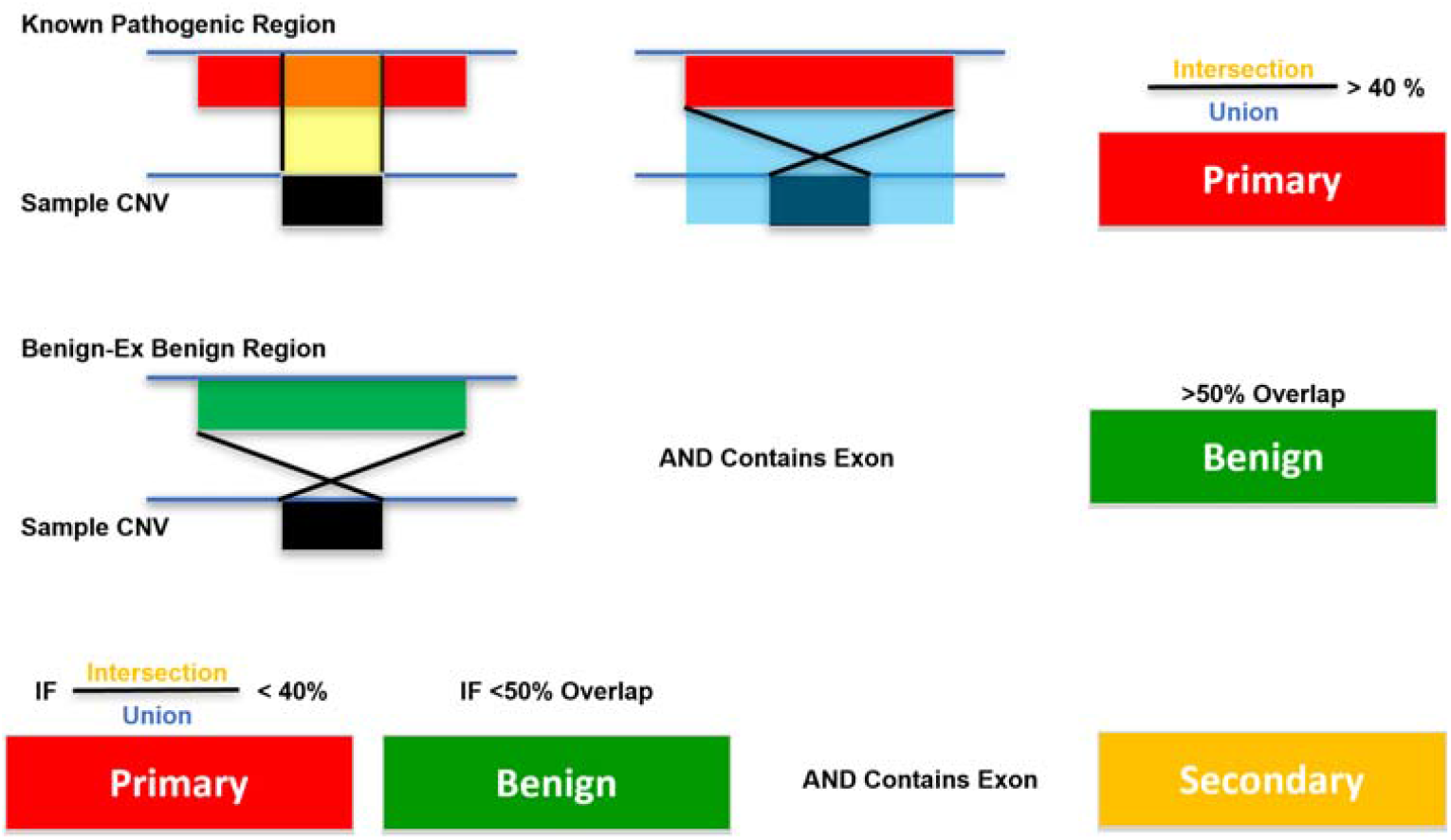
Overview of DISCRIMINATOR. Provisional pathogenicity classifications are assigned in a step-wise manner: primary, benign, or secondary. Sample CNV(s) are classified as primary if the Jaccard similarity index score between the sample CNV and a pathogenic region exceeds a user-defined threshold (default set to 40%; **Supplementary Methods**). Sample CNV(s) which intersect one or more protein-coding gene(s) are classified as benign if the percent overlap between the sample CNV and a benign region exceeds the 50% overlap threshold. All remaining CNV(s) which intersect protein coding gene(s) not classified as primary or benign are classified as secondary. CNV(s) which do not intersect protein-coding gene(s) are classified as non-coding.

### DISCRIMINATOR Optimization

To ensure that DISCRIMINATOR was accurately classifying known pathogenic CNV(s) as Primary CNV(s) from a set of sample CNV(s), we compared the number of primary CNV(s) identified by DISCRIMINATOR for each Jaccard similarity threshold (range: 0.1-0.9; 0.1 intervals) to clinical records for nine loci: 1q21.1, 2p15-p16.1, 15q11.2, 16p11.2, 17p11 (Potocki-Lupski/Smith Magenis/Yuan-Harel-Lupski), 17q21 (Koolen de Vries), 22q11.2 and Xp22.31 (steroid sulfatase; STS), Xp22.33 (Leri-Weill dyschondrosteosis/SHOX haploinsufficiency; SHOX; see **Supplementary Methods**). Nexus Copy Number software and clinical test reports were utilized for the ascertainment of the true number of primary CNVs at each locus for each cohort (**Supplemental Figure 3**). These primary CNV(s) were chosen for analysis because they represent both common MMS CNVs, rare recurrent MMS CNVs, and intervals with complexity. The 1q21.1, 15q11.2 and 22q11.2 regions are complex, with multiple recurrent CNV intervals; therefore, in addition to examining gains and losses individually, a total of 27 distinct primary CNV(s) were investigated across these 9 loci.

### Targeted Re-Analysis of CMA Result Classifications

We compared the provisional CNV classifications obtained from DISCRIMINATOR to the reported clinical CMA result to identify cases for which targeted re-analysis may be necessary; specifically, cases where DISCRIMINATOR identified (1) only benign and/or non-coding CNVs, and the CMA was reported out as a ‘VUS’ or ‘Abnormal’, and (2) one or more primary CNVs, and the CMA was not reported out as ‘Abnormal’. Once identified, the data from these cases underwent re-analysis.

### Statistical Methods

Statistical analyses (Kruskal-Wallis one-way-analysis of variance and Wilcoxon Rank Sum test) were performed with R v4.1.2.

## Results

### Cohort and Diagnostic Yield of CMA

Our study population consists of unique chromosomal microarray (CMA) tests performed at the University of Iowa’s Shivanand R. Patil Cytogenetics and Molecular Laboratory between April 2011 and January 2018 (*n* = 3362). Both diagnostic and familial testing CMAs were included (familial testing refers to the subgroup of cases who underwent targeted CMA testing to either aid in variant interpretation of the proband, or for confirmation of a variant in a family member). Consistent with the reported diagnostic rate ^2,4,6,27^, a clinical diagnosis was reached in 16.78% of cases (*n* = 564; **Figure 2**). Of the 564 abnormal cases, 59 cases (10.5%) represent focused studies for targeted analysis of familial CNVs. A total of 334 recognizable microduplication/microdeletion syndrome (MMS) CNVs were identified by array CGH (9.04% of all cases studied, 53.9% of abnormal cases), and an additional 442 CNVs were reported. These additional reported CNVs (non-MMS) represent both VUS CNVs and non-MMS pathogenic CNVs (*e*.*g*., large *de novo* duplications or deletions). The bulk of identified MMS CNVs (n=208; 62.3%) were found within the 1q21.1 (n=27), 15q11.2 (n=93), 16p11.2 (n=18), 16p13.11 (n=15), and 22q11.2 (n=55) regions (**Supplemental Table 1**). Overall, more pathogenic deletions were reported than duplications (59.5% vs 40.5%; p<0.001), and there was a significant association between the CNV directionality (deletion vs duplication) and CNV subtype (MMS vs other; p=0.002; **Supplemental Figure 4**). Reported MMS CNVs were more likely to be deletions (65.9%; 95% CI: 0.61, 0.71); while there was no significant difference in the proportion of deletions and duplications for non-MMS CNVs. On average, there were 1.56±0.88 CNVs reported per Abnormal CMA (0.92±0.73 deletions; 0.64±0.80 duplications) with no significant difference in the average number of CNVs reported per Abnormal CMA between the three cohorts (Gain: p=0.948; Loss: p=0.923; Total: p=0.773; **Supplemental Figures 5-6**).

**Figure 2:**
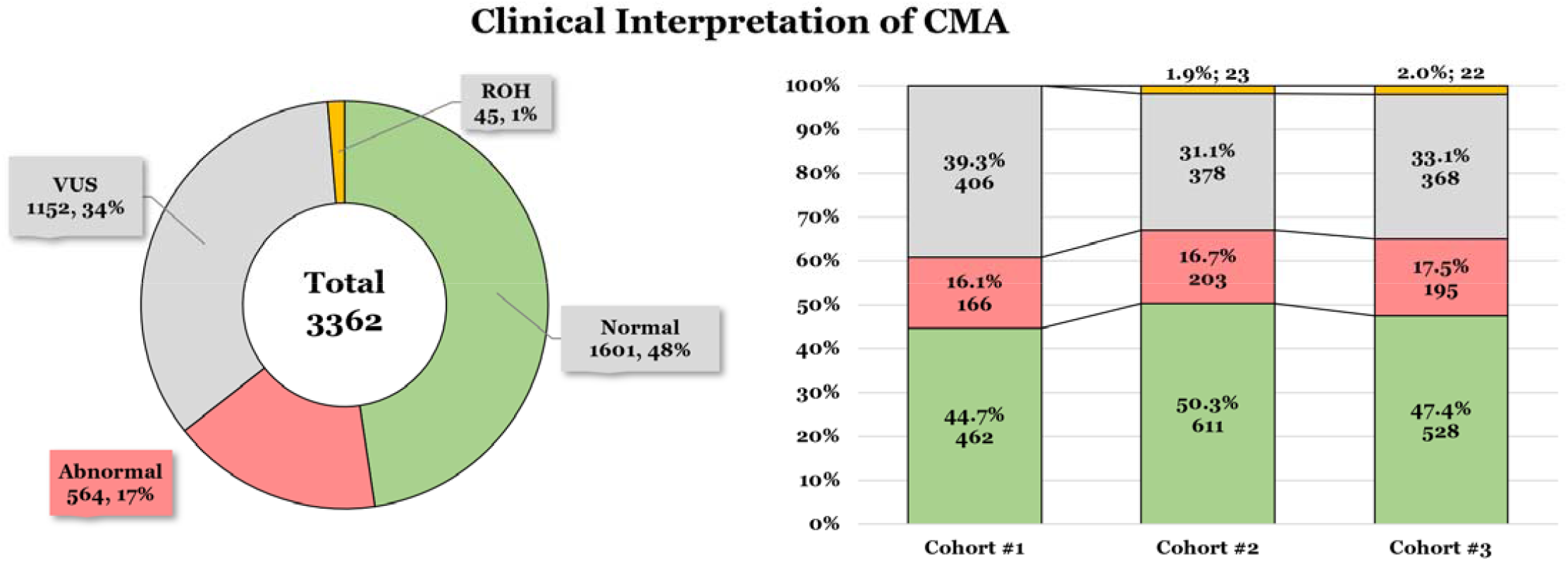
Clinical Interpretation of CMA Results. Left: Pie chart depicting the total number and percentage of cases with a Normal (green), Abnormal (red), VUS (grey), or regions of homozygosity (ROH; yellow) CMA Result. Right: The total number and percentage of cases with Normal, Abnormal, VUS, and ROH CMA results for each individual cohort. ROH events were not reported until the Cohorts #2 and #3 because the NimbleGen 385K and 720k platforms did not have the ability to detect these events.

### CNV Dataset Characteristics

The final CNV dataset consisted of 87,808 CNVs (55,016 deletions and 32,792 duplications; **Table 1**) located in 2,536 CNV regions (CNVRs; 2853 deletion CNVRs and 1519 duplication CNVRs; **Supplemental Figure 7A**) and which accounted for 57.7% of the genome. Of those, 27,414 CNVs were unique (15,180 deletions and 13,194 duplications). The size of CNVs ranged from 20kb to 146 Mb (20,002 – 146,137,722 bp) with a median size of 57.6 kb (56.4-58.3; 95% CI). The length of duplication CNVs (91.0 kb [88.6-92.9]; 95% CI) was significantly longer than deletion CNVs (48.9 kb [48.1-48.8]; 95% CI; χ^2^=5255.9, p<0.001; **Supplemental Figure 7B-C**). There was also a statistically significant difference in CNV length by direction between cohorts (Gain: χ^2^=1365.8, p<0.001; Loss: χ^2^ =13,355, p<0.001; **Table 1; Supplemental Figure 8C-D**). Finally, there was a statistically significant difference in the average number of CNVs (gains, losses, total) identified per case between cohorts (**Table 1; Supplementary Figure 9A**; p<0.001).

**Table 1:**
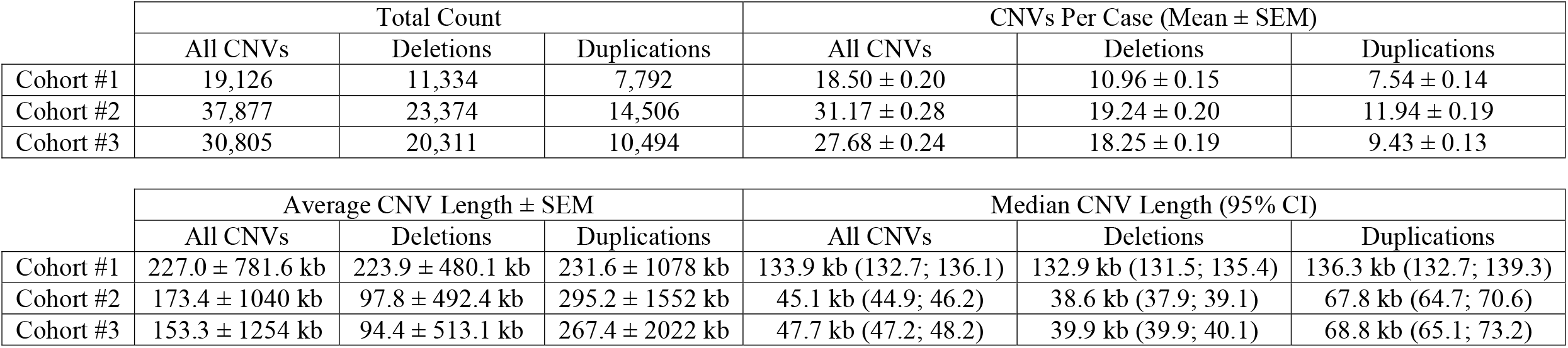
Descriptive CNV Metrics.

### DISCRIMINATOR Optimization

The total number of Primary CNV(s) identified by DISCRIMINATOR at each Jaccard similarity threshold across the three cohorts ranged from 3,655 (Jaccard = 0.1) to 155 (Jaccard = 0.9; **Supplementary Figure 10**). DISCRIMINATOR accurately identified both gain and loss primary CNVs across each of the 9 loci at the 0.40 and 0.41 Jaccard threshold levels (**Supplementary Figures 11**). As the total number of Primary CNVs identified by DISCRIMINATOR was the same for 0.40 and 0.41 across these loci, the Jaccard similarity threshold of 0.40 was chosen for the final value.

### Provisional CNV Classifications

The majority of identified CNVs were classified as ‘BENIGN’ (n=42,824; 48.8%) or ‘NON-CODING’ (n=39,009; 44.4%; **Figure 3**; **Supplemental Figure 12; Supplemental Table 2**). Six percent of CNVs were classified as Secondary CNVs (n=5,356), and 0.7% were classified as PRIMARY CNVs (n=619). Consistent with their prevalence rates, 15q11.2, 16p11.2, and 22q11.2 were the most common primary CNVs detected by DISCRIMINATOR (**Figure 4**). The 619 Primary CNVs were identified across 557 cases, with the vast majority having a single Primary CNV (*n*=499; 89.6%). There were a handful of situations in which mulitple Primary CNVs were identified in the same case (2 Primary CNVs: *n*=54; 3 Primary CNVs: *n*=4). In addition, there were 864 cases in which CNVs were provisionally classified as either Benign or Non-Coding CNVs only (25.7%). There was no significant difference in the average number of Primary CNVs identified per case by cohort (p=0.06), but there was a significant difference in the average number of Secondary, Benign, and Non-Coding CNVs identified per case by cohort (Secondary: χ^2^=535.73, p<0.001; Benign: χ^2^=67.295, p<0.001; Non-Coding: X^2^= 1984.6, p<0.001; **Supplementary Figure 13**). The average number of Primary CNVs, Secondary CNVs, Benign CNVs, and Non-Coding CNVs per case by cohort can be found in **Supplementary Table 3**.

**Figure 3:**
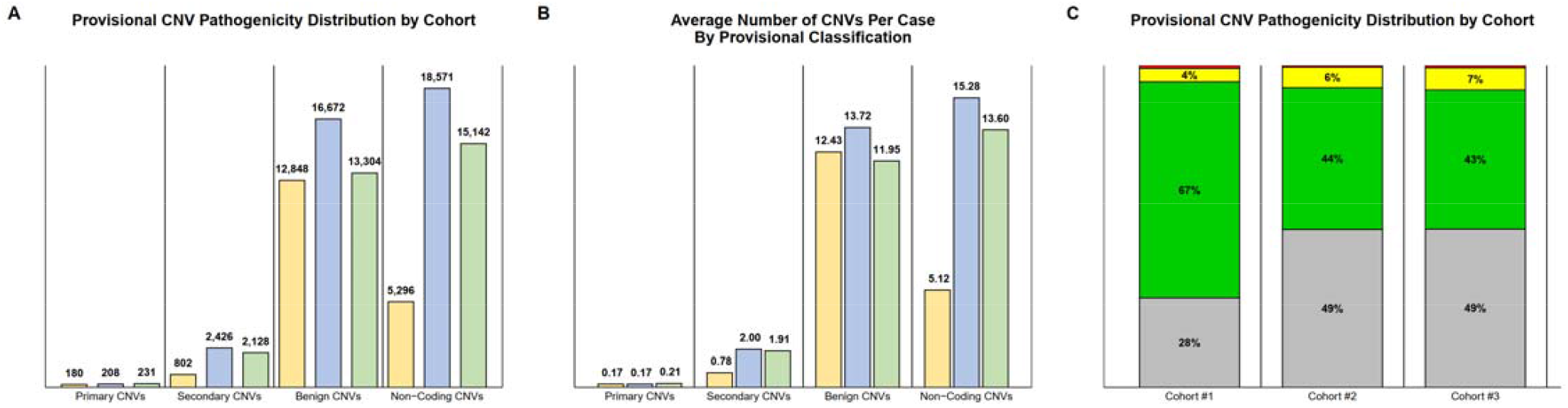
DISCRIMINATOR Provisional Pathogenicity Classifications. (A) Distribution of Provisional CNV Pathogenicity by Cohort. (B) Average Number of CNVs of each Provisional Classification by Cohort. (C) Relative Distribution of Provisional CNV Classifications by Cohort. CNVs identified in each cohort are color coded (Cohort #1: yellow, Cohort #2: blue, and Cohort #3: green). Provisional pathogenicity classifications are color coded (grey: non-coding; green: benign; yellow: secondary; red: primary).

**Figure 4:**
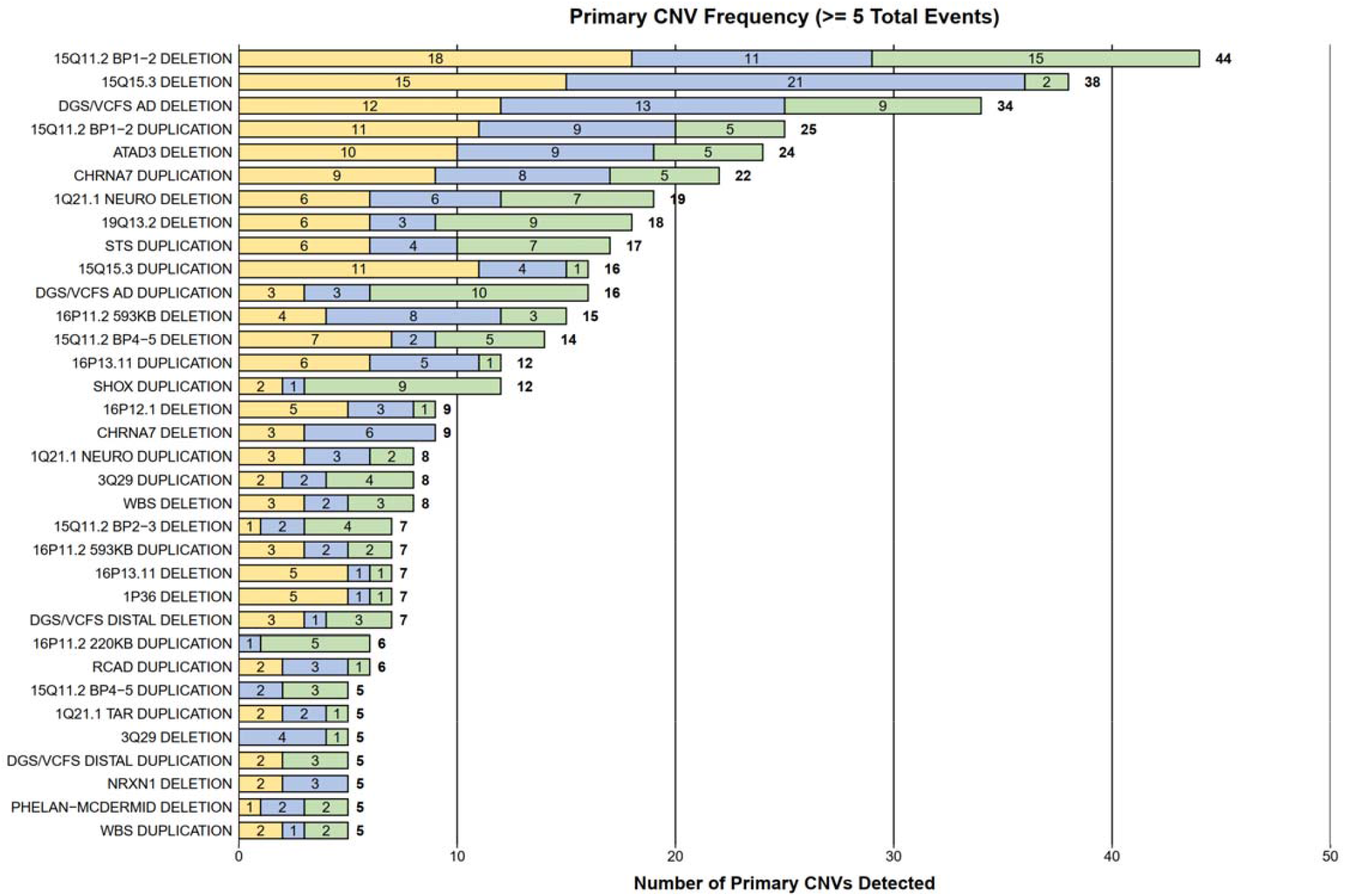
Primary CNVs Identified by DISCRIMINATOR in 5+ Cases. This graph depicts all Primary CNVs identified in at least five cases, broken down by the cohort in which the Primary CNV was detected. Consistent with their prevalence rates, 22q11.2, 15q11.2, and 16p11.2 primary CNVs were among the most common Primary CNVs detected by DISCRIMINATOR. CNVs identified in each cohort are color coded (Cohort #1: yellow, Cohort #2: blue, and Cohort #3: green). Abbreviations: DGS/VCFS: DiGeorge syndrome/Velocardiofacial syndrome; WBS: Williams-Beuren syndrome; RCAD: renal cysts and diabetes (RCAD) syndrome.

### Re-analysis of CMA Results

In total, 102 CMA cases (3.03%) underwent re-analysis based on discrepancies between the reported CMA result and DISCRIMINATOR CNV provisional classification(s). A total of 85 CMA cases (83.3%) were identified that were *not* reported out as ‘Abnormal’ but contained one or more Primary CNVs (12 ‘Normal’, 14.1%; 73 ‘VUS’ 85.9%), and 17 CMAs that were reported out as ‘VUS’ but contained no Primary or Secondary CNVs (16.7%; **Table 2**). There were no CMA cases in which DISCRIMINATOR missed a Primary CNV in an ‘Abnormal’ case – indicating that DISCRIMINATOR identified all Primary CNVs in the examined cohorts.

**Table 2:** Targeted Re-Analysis of CMA Results.

| DISCRIMINATOR Provisional Classification Findings |  |  |
| --- | --- | --- |
| CMA Clinical Report | 1+ Primary CNV Identified | No Primary or Secondary CNVs<br>(*i.e.* only Benign/Non-Coding) |
| Abnormal | --- | 0 |
| Normal | 12 | -- |
| VUS | 73 (18) | 17 (10) |

There were 12 CMA cases which were reported out clinically as ‘Normal’, but in which DISCRIMINATOR identified a Primary CNV (**Table 2**). These cases fall into three categories: (1) familial testing (*n*=4), (2) false positives (*n*=4), and (3) clinical vs non-clinical results (*n*=4). However, given that clinical guidelines for reported CNVs are stricter than the thresholds used in this quality improvement study, there are no true cases in which a ‘Normal’ clinical CMA report contained a Primary CNV, and therefore, no cases that are candidates for clinical re-classification. Four cases represent focused familial testing wherein the Primary CNV identified by DISCRIMINATOR is not the CNV that was being investigated clinically. For completeness, we chose to include all reciprocal deletion and/or duplication events within the Primary CNV list supplied to DISCRIMINATOR regardless of whether these reciprocal events have been observed or associated with a clinically reported disorder. The four false positive calls that fall into this category are all duplication events that are either thought to be benign, or regions in which triplosensitivity has not been evaluated. Lastly, in four cases, the clinical calling thresholds differed from those used in this study (e.g. log2 threshold of 0.3 vs 0.25), and therefore the Primary CNV was not reported clinically.

Similarly, there were 73 CMA cases that were reported out clinically as a ‘VUS’, but in which DISCRIMINATOR identified a Primary CNV. Excluding the CMA cases which are ineligible for reclassification (Familial Testing (*n*=17); False Positives (*n*=35; *CHRNA7: n*=17; *SHOX*: *n*=7; *STS*: *n*=7); Non-Clinical Results (*n*=3)), there were 18 CMA cases that represent true cases of CNVs whose pathogenicity interpretation may require reclassification from VUS to Pathogenic or Likely Pathogenic. Of these, nearly all cases involve duplication events (*n*=15; **Supplemental Table 4**) and Primary CNVs identified in the 15q11.2 (*n*=6), 16p11.2 (*n*=4), 16p12.1 (n=3), or 20p12.1 (*n*=2) regions.

Lastly, there were 17 cases that were reported out clinically as a ‘VUS’ and in which DISCRIMINATOR classified all identified CNVs as either ‘benign’ or ‘non-coding’. Seven of these cases were focused familial studies and thus excluded from further analysis. The remaining cases were primarily from Cohort #1 (*n*=9), representing duplication events (*n*=8), and all reported VUS CNVs were considered benign or likely benign variants upon re-analysis (*n*=10).

## Discussion

DISCRIMINATOR was developed to aid in cohort analysis and re-analysis of CMA results and the prioritization of CNVs. Here we demonstrate that DISCRIMINATOR can reliably detect known pathogenic CNVs and provisionally classify them as ‘Primary’, thus identifying a subset of cases in which CMA re-analysis may be warranted. In addition, we also present a cohort of 3362 cases who underwent CMA testing and the results of that testing.

Our data shows that while there was no significant difference in the number of CNVs that were reported out clinically, there was a significant difference between all three cohorts with regards to the size and number of CNVs identified. The difference in array platform including probe design and coverage (NimbleGen 385K, 720K vs Affymetrix CytoscanHD) likely explains the differences seen between Cohort #1 and Cohorts #2/#3. While there were differences in the average number of Benign, Secondary, and Non-Coding provisional classifications between all three cohorts, the difference between Cohort #1 and Cohort #2/#3 appears greatest for Non-coding and Secondary CNVs (**Supplemental Figure 13**). This can be attributed to the increased resolution of the Affymetrix CytoscanHD array, as the number of small CNVs (<50kb) is increased in Cohorts #2/#3 compared to Cohort #1 (**Supplemental Figure 8B**). Further, the distribution of secondary and non-coding CNVs (particularly deletion events) is right-skewed towards smaller variants (**Supplemental Figures 12B**).

Cohorts #2 and #3 were run on the same array platform (Affymetrix CytoscanHD) but in two different facilities, and this change in facilities and subsequent reduction of background noise is likely responsible for the observed differences between these cohorts. The move to a newer facility allowed for better control and standardization of the ambient temperature and humidity, and these are factors that have been shown to influence microarray quality ^33–35^. There was a significant difference in the ‘noise’ or quality of the CMAs that were performed between Cohort #2 and Cohort #3 (**Supplemental Figure 14A**), with significantly less noise across Cohort #3. Temporal mapping of noise also revealed this cohort-level effect (**Supplemental Figure 14B**). Cohort #2 and Cohort #3 were found to be significantly different in terms of the average number of deletions and duplications identified per case on average, as well as the average length of deletion CNVs. In addition, there were more duplication and deletion (and therefore total) CNVs identified in Cohort #2 compared to Cohort #3 (**Supplemental Figure 9A**). These findings therefore are consistent with a reduction of background noise and spurious calls in Cohort #3.

The provisional classifications provided by DISCRIMINATOR require further investigation before ‘Pathogenic’ or ‘Likely Pathogenic’ labels can be applied to clinically identified CNVs for a few main reasons. First, the overlap of a case CNV with gene(s) and/or region(s) of haploinsufficiency is only a single criterion by which Riggs *et al*. propose evaluating CNVs ^24,25^. We also used a more lenient set of criteria for calling CNVs than is utilized clinically and chose to include all reciprocal events for all identified MMS. Finally, the classifications provided by DISCRIMINATOR are dependent on the set of user-defined benign and pathogenic intervals and gene coordinates. We chose to limit the overlap of genes to those that are ‘known protein coding’; therefore, users could generate a different distribution of benign, non-coding, and secondary CNVs if the ‘known protein coding’ limitation is relaxed. There will, however, be no change in the identification of Primary CNVs unless the pathogenic interval file is changed, as those are assessed independent of genic content.

Lastly, we identified several cases where DISCRIMINATOR identified a ‘Primary’ CNV within a case that had a ‘VUS’ CMA test result, and several cases where DISCRIMINATOR identified only benign and/or non-coding CNVs in cases with a ‘VUS’ CMA test result. Targeted re-analysis of these cases demonstrates the need for regular re-interpretation of ‘VUS’ CMAs as the guidelines for interpretation are updated. Uncoupling the clinical phenotype with the classification of variant pathogenicity allows for more consistent interpretations across and within laboratories, and retrospective analysis can lead to the identification of cases with inconsistent variant pathogenicity assessments. Targeted re-analysis in our cohorts led to a few major findings: First, when undergoing cohort-level analysis of CNV data, there will be a percentage of secondary findings identified in cases that underwent focused familial testing. Second, as research progresses and we are better able to interpret deletions and duplications in the human genome, our ability to classify variants improves. This is exemplified by (1) the observation that many of the cases that were reported out as ‘VUS’ and harbored a Primary CNV were within the 15q and 16p regions and (2) the majority of the cases that were reported out as a ‘VUS’ and only contain benign and/or non-coding CNVs by DISCRIMINATOR originated in Cohort #1. Cohort #1 was interpreted in a time (2011-2012) at which we had the least medical knowledge about CNV consequence; thus, was likely to exhibit higher levels of reclassification.

The ACMGG has no formal guidelines on the frequency of re-analysis of CMA data and instead suggests that individual clinical laboratories establish regular re-analysis as part of their protocol ^36^. Further, they suggest that upon the publication of new control databases and/or gene-disease relationships, a clinical laboratory’s entire variant set should undergo partial review to identify a subset of variants for re-analysis. DISCRIMINATOR could be used in these circumstances to quickly and efficiently identify if any clinically identified variants exist which may need to undergo further review.

DISCRIMINATOR has been optimized to identify Primary CNVs from *a specific list of pathogenic CNVs/regions*. It is possible that an alternative pathogenic region list will not perform as well at the 0.4 Jaccard threshold level, and therefore, further testing might be necessary for re-optimization. We chose to expand our analysis to variants below our clinical threshold (50kb); therefore, it is important to distinguish between re-analysis for clinical and non-clinical purposes. Lastly, DISCRIMINATOR is also dependent on the quality of the CNV calls that are supplied.

The value of DISCRIMINATOR is not limited to what we have shown here. The cohort-wide metrics provided by DISCRIMINATOR, including the number of identified CNVs at different size thresholds, could be used in quality control monitoring. By monitoring the relative proportions of Benign, Secondary, and/or Non-Coding CNVs, the relative proportions of deletions and deletions, or the relative proportions of CNVs at different size thresholds across CMA batches, a metric could be created to assess for drift or other indicators of shifts in clinical CMA quality. The set of benign and pathogenic intervals could be modified for the validation of a new CNV detection assay, or to quickly scan large cohorts for a specific list of candidate pathogenic CNVs or regions of interest. Additionally, due to the significant time and effort involved, re-analysis of clinical CMA data on a large scale is not routinely performed and is a longstanding challenge. However, because DISCRIMINATOR can operate on the cohort level, processing thousands of CNVs concurrently, and employ any benign or pathogenic interval list, DISCRIMINATOR can be used to continually re-evaluate and re-analyze clinical CMA data with the most up-to-date versions of what is considered benign and/or pathogenic.

## Data Availability

DISCRIMINATOR is publicly available on GitHub at https://github.com/aswetzel/DISCRIMINATOR. All non-case data produced and analyzed in the present study are included in this published article and its supplementary information. Case data is available upon reasonable request to the authors.

## List of abbreviations

CMA: clinical chromosomal microarray
CNVs: copy number variants
MMS: microdeletion and microduplication syndromes

## Acknowledgements

This study makes use of data generated by the DECIPHER community. A full list of centres who contributed to the generation of the data is available from http://decipher.sanger.ac.uk and via email from. Funding for the DECIPHER project was provided by Wellcome ^1^. We would also like to thank the members of the University of Iowa’s Shivanand R. Patil Cytogenetics & Molecular laboratory including its past directors Drs. Val Sheffield and Shivanand Patil.

Literature Cited

