## Supplemental Materials for "DISCRIMINATOR: Assigning Cohort-Wide Provisional Pathogenicity Classifications to CNVs"

**Supplementary Methods:**

**Supplemental Tables:**

**Supplemental Figures:**

Supplementary Methods:

**1 Microarray Data Curation**

: The NimbleGen 385K microarray is comprised of 385,000 copy number probes with an average probe spacing of 6,270bp, and the NimbleGen 720K microarray is comprised of 720,412 copy number probes with an average probe spacing of 2,500bp. Copy number variants for Cohort #1 (NimbleGen 385K and NimbleGen 720K microarray data) were identified from raw microarray data with Nexus Copy Number software (Versions 4.0-9.0; BioDiscovery Inc) using the proprietary FASST2 segmentation algorithm. The Affymetrix CytoscanHD microarray is comprised of ~2.7 million copy number (1,953,246) and SNP (743,304) probes, with an average probe spacing of 880bp. Copy number variants for Cohorts #2 and #3 (Affymetrix CytoscanHD microarray data) were identified from raw microarray data with Chromosome Analysis Suite (Versions 7-9) using their proprietary CNV detection algorithm**.** Nexus Copy Number software (Versions 4.0-9.0; BioDiscovery Inc) was used for visualization and curation of microarray data.

Likely due to the unique nature of the probe coverage distribution of the NimbleGen arrays, we identified several cases that had 22q11.2 A-D duplications and/or deletions, but whose duplication/deletion was segmented into two unique CNVs (22q11.2 A-B and 22q11.2 B-D; **Supplemental Figure 15**). We expanded our analysis to several other recurrent CNV regions looking for segmented CNV calls and identified 25 cases across the three different array platforms where two distinct primary CNV events were being called (22q11.2 locus: n=20; 15q11.2 locus: n=3; 1q21.1 locus: n=1; 17p12 locus: n=1). Therefore, we devised a series of criteria which relaxed the merging constraints exclusively for these regions (**Supplemental Figure 16**). To be merged, sample CNVs need to overlap the Primary CNV regions of interest (1q21.1, 15q11.2, 17q12, and 22q11.2) but *not* completely overlap segmental duplications (100% identity). The Primary CNV region of interest was defined as the whole interval from the most proximal to the most distal coordinate of the DISCRIMINATOR Primary CNVs in the region. For the 22q11.2 interval, this corresponds to the start coordinate of the DGS/VCFS A-B deletion/duplication to the end coordinate of the DGS/VCFS D-F deletion/duplication. To facilitate removal of CNVs by segmental duplication content, the Segmental Duplication (SegDup) track (last updated: 2011-09-26)^1,2^ was downloaded from the UCSC Table Browser ^3,4^ and individual segdup calls were merged if they were within 50kbp of one another. Next, we required consecutive sample CNVs to be less than 1 Mb apart, and in the same direction. Finally, sample CNVs were recursively merged only if the region between the two CNVs was (a) less than 100kbp, (b) overlapped by a SegDup cluster (>90% identity) or (c) overlapped the 22q11.2 "B" SegDup cluster (>90% identity; chr22:20249559-20744436 (hg19)). This supplemental merging step ('recurrent CNV merging') is performed by DISCRIMINATOR and users can modify the regions of interest(s) and/or turn off this merging step.

**2 DISCRIMINATOR Input Datasets**

**:** DISCRIMINATOR will accept any set of benign interval file(s), primary CNV interval file(s), and gene and/or exon file for assigning provisional pathogenicity classifications and determining gene/exon overlap provided those files meet the formatting requirements.

Coordinates for the gene and exon files were downloaded from UCSC Table Browser ^3,4^ using the GENCODE V39lift37 Basic track (hg19.wgEncodeGencodeBasicV39lift37; 2022-01-16 release) ^5,6^. We restricted our list to known protein-coding genes by employing the filters transcriptClass="coding" and geneType="protein_coding" against the linked attributes table (wgEncodeGencodeAttrsV39lift37).

Coordinates for the benign interval files were generated using the 2020-02-25 release of the Database of Genomic Variants (DGV) ^7^ and the 2021-04-01 release of the Clinical Genome Resource (ClinGen) ^8^ in conjunction with Benign-Ex (Wetzel *et al.*; manuscript in preparation ^9^) with the parameter settings optimized against the included Primary CNV List: DGV Gain (2_30_GAIN_0.004_N_2009), DGV Loss (2_30_LOSS_0.006_N_2009), ClinGen Gain (1_B_USLB_GAIN) and ClinGen Loss (1_B_USLB_LOSS). The set of benign intervals obtained using DGV consists of those variants which were observed two or more times with an allele frequency (AF) greater than 0.004% (Gain Events) or 0.006% (Loss Events). Each of these 2+ variants must also come from a study published in or after 2009 with a sample size greater than 30. The set of benign intervals obtained using ClinGen were all variants with a 'benign' or 'likely benign' classification.

Coordinates for the Primary CNV interval file were derived and modified from a manually curated list of microdeletion and microduplication syndromes (Wetzel and Darbro; in press ^10^). We utilized both the set of reported CNVs (Dataset 2 of Wetzel and Darbro) and a set of nine representative loci (see **DISCRIMINATOR Optimization**) to optimize the coordinates of the primary CNV intervals for DISCRIMINATOR.

**3 Detailed DISCRIMINATOR Methodology**

**:** DISCRIMINATOR is a python-based program which assigns provisional pathogenicity classifications (Primary, Secondary, Benign, Non-Coding) to sample CNVs. DISCRIMINATOR runs from the wrapper file 'INTERFACE_DISCRIMINATOR.py' which calls each of the sub-scripts (**Supplemental Figure 17**). First, DISCRIMINATOR reads the input files (COHORT_FILES, BENIGN_FILES, PRIMARY_CNV_FILES, JACCARD_INDEX) to determine which cohort files to process, which files contain the benign and pathogenic region intervals, and determine the user defined Jaccard Index Threshold(s). Next, DISCRIMINATOR will process the set of benign files provided by BENIGN_FILES to generate a singular benign file which consists of the merged benign regions.

For each sample CNV, DISCRIMINATOR will determine if (a) it overlaps any protein-coding gene(s) and/or exon(s), and (b) if it intersects with any benign and/or primary interval(s) (**Supplemental Figure 2**). For each identified intersection, DISCRIMINATOR will calculate the percent overlap (sample CNV vs benign interval) and/or Jaccard Index (sample CNV vs primary interval). Sample CNV(s) which meet or exceed the user-defined Jaccard Index threshold are classified as 'Primary' and annotated with the name of the Primary CNV. If a sample CNV intersects multiple primary intervals, DISCRIMINATOR will annotate the sample CNV with the Primary CNV which has the greatest Jaccard Index value. Any non-primary sample CNV(s) which do not overlap a protein-coding gene are classified as 'Non-Coding'. Sample CNV(s) which meet or exceed the user-defined percent overlap are classified as 'Benign', while all remaining CNV(s) which overlap a protein-coding gene and do not meet the Jaccard index or percent overlap thresholds are classified as 'Secondary'. All Primary, Secondary, and Benign CNVs are also annotated with the set of gene(s) and exon(s) which it contains/overlaps.

After establishing the classifications for each sample CNV, DISCRIMINATOR will generate a descriptive cohort-wide metrics file. Notably, this file contains the (1) total number of Primary, Secondary, Benign, and Non-Coding CNVs identified across the cohort, (2) the average number of Primary, Secondary, Benign, and Non-Coding CNVs per sample across the cohort, and (3) the total number of samples with each identified Primary CNV in the cohort. Additional information includes details on co-occurring primary and secondary CNVs for each primary CNV, and the total number and average number of CNVs in each classification for each direction (i.e. gain, loss) and a number of different size thresholds (>50kb, >100kb, >500kb, >1Mb).

**4** **DISCRIMINATOR Optimization**

**:** The number of Primary CNV(s) identified by DISCRIMINATOR for each Jaccard Index Threshold was compared to clinical records for nine loci: 1q21.1 (**Supplemental Figure 18**), 2p15-p16.1, 15q11.2 (**Supplemental Figure 19**), 16p11.2 (**Supplemental Figure 20**), 17p11 (Potocki-Lupski/Smith Magenis/Yuan-Harel-Lupski; **Supplemental Figure 21**), 17q21 (Koolen de Vries; **Supplemental Figure 22**), 22q11.2 (**Supplemental Figure 23**), Xp22.33 (Leri-Weill dyschondrosteosis/SHOX haploinsufficiency; SHOX; **Supplemental Figure 24**), and Xp22.31 (steroid sulfatase; STS; **Supplemental Figure 25**). There were no cases with CNVs in the 2p15-p16 CNV region, and DISCRIMINATOR did not detect any 2p15-p16 Primary CNVs at any Jaccard Threshold.

The total number of Primary CNV(s) identified by DISCRIMINATOR ranged from 3,655 (Jaccard = 0.1) to 155 (Jaccard = 0.9; **Supplemental Figure 10**. The Jaccard threshold of 0.4 performed the best across all loci and sub-intervals. To confirm this finding, we repeated this analysis using a narrower Jaccard similarity threshold range with more frequent steps (0.30-0.50; 0.01 intervals; **Supplemental Figure 11; Supplemental Figures 18-25**) to optimize the Jaccard threshold value. DISCRIMINATOR identified the correct number of Primary CNVs for each sub-interval in the nine loci for the 0.40 and 0.41 Jaccard thresholds. Of these two thresholds, we chose 0.40 as the final Jaccard threshold value.

**Supplemental Table 1: Reported MMS CNVs**

**.**

| **CNV Region** | **Deletions** | **Duplications** | **Total** |  | **CNV Region** | **Deletions** | **Duplications** | **Total** |
| --- | --- | --- | --- | --- | --- | --- | --- | --- |
| **1p36** | 5 | 1 | 6 |  | **16p11.2** | 16 | 2 | 18 |
| **1q21.1** | 19 | 8 | 27 |  | **16p12.1** | 4 | 0 | 4 |
| **2p21** | 1 | 0 | 1 |  | **16p11.2-p12.2** | 0 | 1 | 1 |
| **NPHP1** | 8 | 3 | 11 |  | **16p13.11** | 5 | 10 | 15 |
| **3p** | 4 | 0 | 4 |  | **ATR-16** | 1 | 0 | 1 |
| **3q29** | 3 | 7 | 10 |  | **PTLS/SMS** | 1 | 1 | 2 |
| **Wolf-Hirshorn** | 2 | 0 | 2 |  | **CMT1A/HNPP** | 1 | 0 | 1 |
| **5p** | 2 | 0 | 2 |  | **Yuan-Harel-Lupski** | 1 | 3 | 4 |
| **Cri-du-Chat** | 1 | 0 | 1 |  | **NF1** | 2 | 0 | 2 |
| **5q35** | 1 | 0 | 1 |  | **17q12** | 3 | 5 | 8 |
| **WBS** | 8 | 5 | 13 |  | **RCAD** | 0 | 1 | 1 |
| **8p23.1** | 0 | 2 | 2 |  | **Koolen de Vries** | 1 | 1 | 2 |
| **9p(-)** | 1 | 2 | 3 |  | **18p** | 5 | 2 | 7 |
| **10q22-q23** | 4 | 0 | 4 |  | **Cat-Eye** | 5 | 2 | 7 |
| **10q26** | 2 | 0 | 2 |  | **22q11.2** | 39 | 16 | 55 |
| **ELP4** | 1 | 0 | 1 |  | **Xp11.22-p11.23** | 0 | 1 | 1 |
| **Jacobsen** | 1 | 0 | 1 |  | **STS** | 2 | 8 | 10 |
| **15q11.2** | 64 | 29 | 93 |  | **SHOX** | 7 | 2 | 9 |
| **15q26** | 0 | 1 | 1 |  | **MECP2** | 0 | 1 | 1 |

**Supplemental Table 2: Average Number of CNVs Per CMA by Provisional Pathogenicity Classifications and Cohort**

**.** Mean CNVs Per CMA ± SEM

|  | **Cohort #1** | **Cohort #2** | **Cohort #3** |
| --- | --- | --- | --- |
| **Primary** | 0.17±0.01 | 0.17±0.01 | 0.21±0.01 |
| **Secondary** | 0.78±0.05 | 2.00±0.06 | 1.91±0.1 |
| **Benign** | 12.43±0.14 | 13.72±0.15 | 11.95±0.12 |
| **Non-Coding** | 5.12±0.08 | 15.28±0.14 | 13.6±0.11 |

**Supplemental Table 3: Total Number of CNVs for each Provisional Pathogenicity Classification by Cohort**

**.**

|  | **Cohort #1** | **Cohort #2** | **Cohort #3** | **Total** |
| --- | --- | --- | --- | --- |
| **Primary** | 180 | 208 | 231 | 619 |
| **Secondary** | 802 | 2,426 | 2,128 | 5,356 |
| **Benign** | 12,848 | 16,672 | 13,304 | 42,824 |
| **Non-Coding** | 5,296 | 18,571 | 15,142 | 39,009 |
| **Total** | 19,126 | 37,877 | 30,805 | 87,808 |

**Supplemental Table 4: CNV Cases with 1+ Primary CNV Identified by DISCRIMINATOR and a 'VUS' Clinical Report**

**.**

|  | **Deletion** | **Duplication** | **Total** |
| --- | --- | --- | --- |
| **MYTIL** | 0 | 1 | 1 |
| **4q21** | 0 | 1 | 1 |
| **15q11.2 BP1-2** | 1 | 3 | 4 |
| **15q11.2 BP4-5** | 0 | 2 | 2 |
| **16p11.2 220kb** | 0 | 3 | 3 |
| **16p11.2 593kb** | 0 | 1 | 1 |
| **16p12.1** | 1 | 2 | 3 |
| **20p12.3** | 0 | 2 | 2 |
| **20p13** | 1 | 0 | 1 |

**Supplemental Figure 1: Flowchart depicting workflow for curating microarray data**

**. PAR: pseudoautosomal region.**

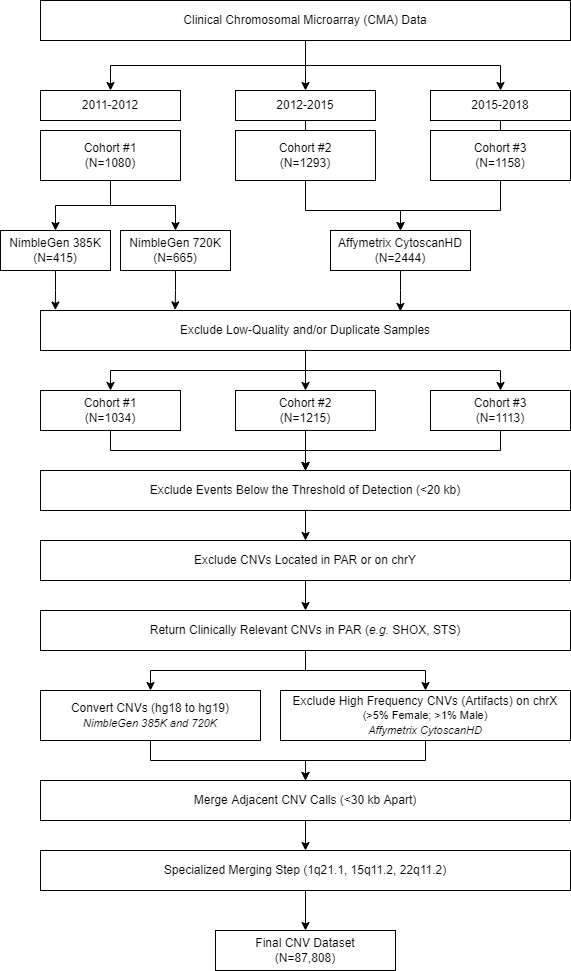

**Supplemental Figure 2: DISCRIMINATOR Provisional Classification Schematic**

**.**

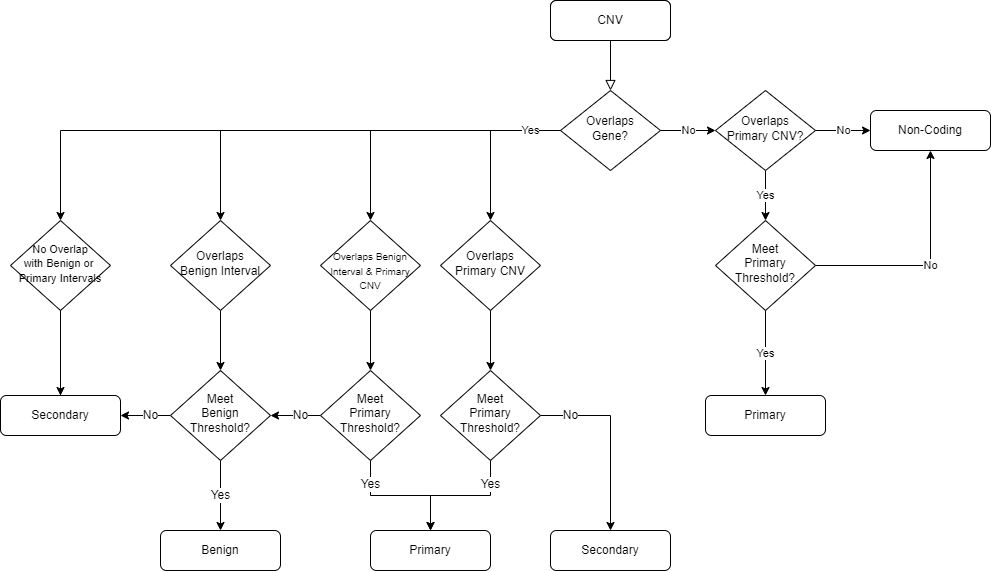

**Supplemental Figure 3: Identifying the Optimal Jaccard Threshold for Calling Primary CNVs in DISCRIMINATOR**

**.** Depicted are the number of CNVs DISCRIMINATOR identified at each Jaccard value (range: 0.1-0.9) in each cohort for the following Primary CNVs: (a) 22q11.2 Microduplication, (b) 16p11.2 Microdeletion, (c) 1q21.1 Neuro Deletion, (d) 22q11.2 Microdeletion, (e) Smith-Magenis Microdeletion, and (f) Koolen de Vries Microduplication. The true number of primary CNVs is depicted on the far right of each graph. CNVs identified in each cohort are color coded (Cohort #1: yellow, Cohort #2: blue, and Cohort #3: green).

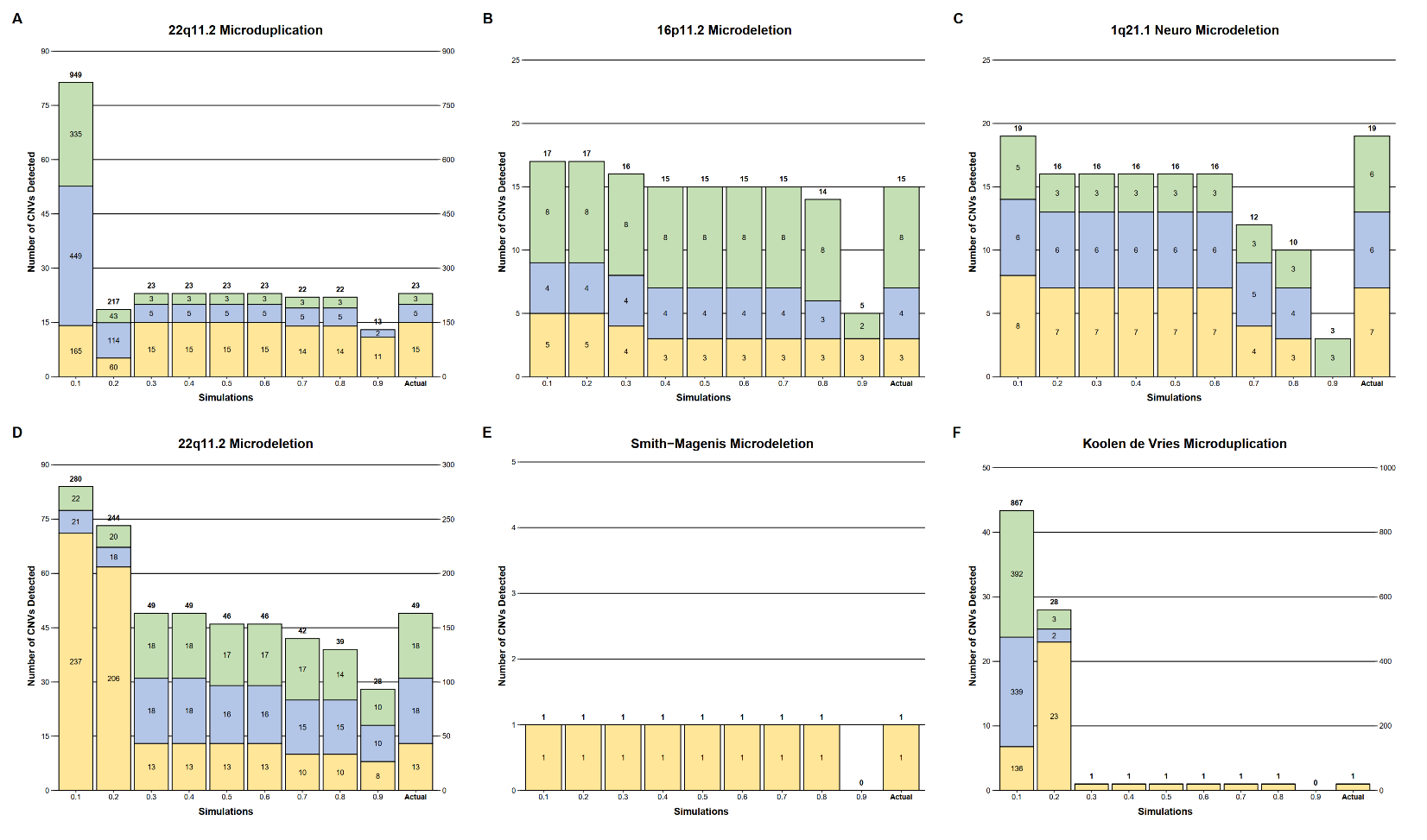

**Supplemental Figure 4: Proportion of Deletions and Duplications for Reported MMS and Non-MMS CNVs in Abnormal Array Results**

**.** The total number of duplications (blue) and deletions (red) reported on Abnormal CMA results which were classified as either MMS CNVs or Non-MMS CNVs (e.g. large de novo pathogenic CNVs and VUS CNVs). The ratio of deletion and duplication CNVs which were reported is different between MMS and non-MMS CNVs (χ^2^= 9.304, p=0.0023) as assessed by Two-Proportions Z-Test. Non-MMS CNVs were equally likely to be deletions or duplications, while there was a significant larger proportion of deletion MMS CNVs compared to duplication MMS CNVs (p<0.001) as assessed by One-Proportions Z-Test with Bonferroni multiple-test correction.

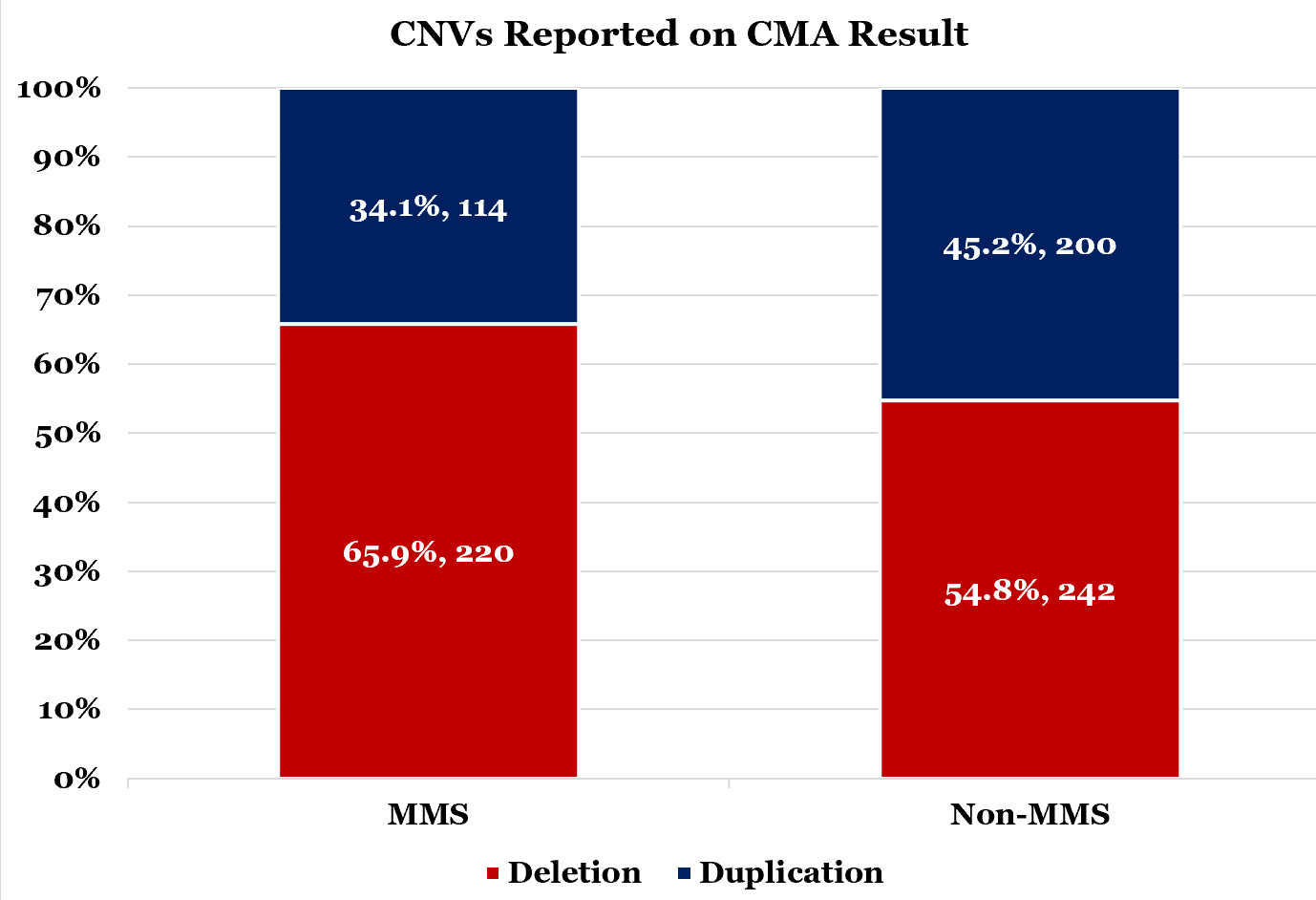

**Supplemental Figure 5: Average Number of Reported CNVs Per Case for Abnormal CMA Results**

**.** Bar plot depicting the average number of CNVs reported (deletions, duplications, and total) per case for CMAs that had Abnormal results in each cohort (Cohort #1: yellow; Cohort #2: blue; Cohort #3: green). Error bars show standard error of the mean. There were no significant differences in the average number of reported deletions, duplications, or total CNVs between cohorts as assessed by Kruskal-Wallis rank sum test with Benjamini & Hochberg multiple-test correction (Gain: p=0.948; Loss: p=0.923; Total: p=0.773).

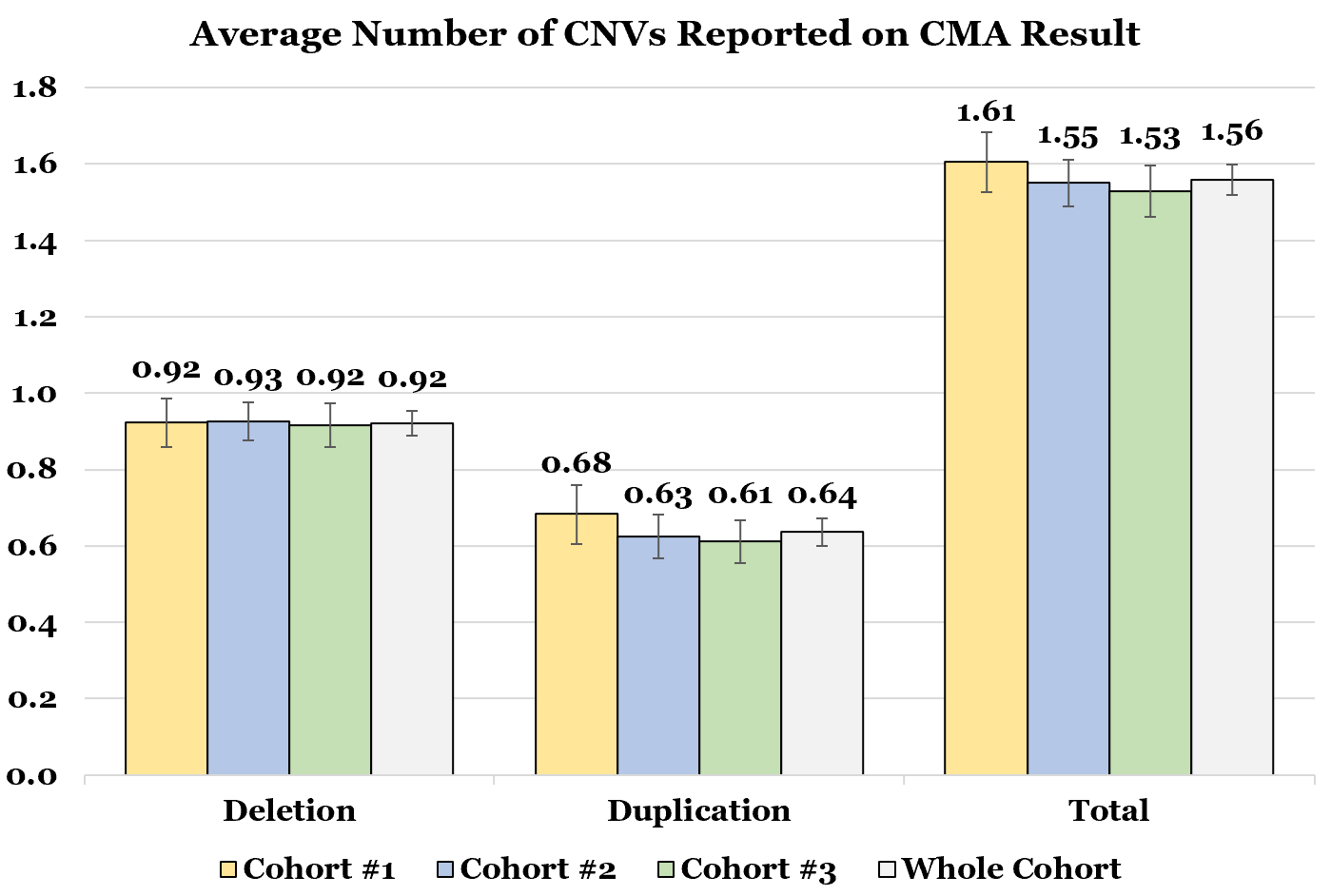

**Supplemental Figure 6: Proportion of Deletions and Duplications Identified by Cohort**

**.** The total number and percentage of duplications (blue) and deletions (red) reported on all clinical CMA reports for each cohort.

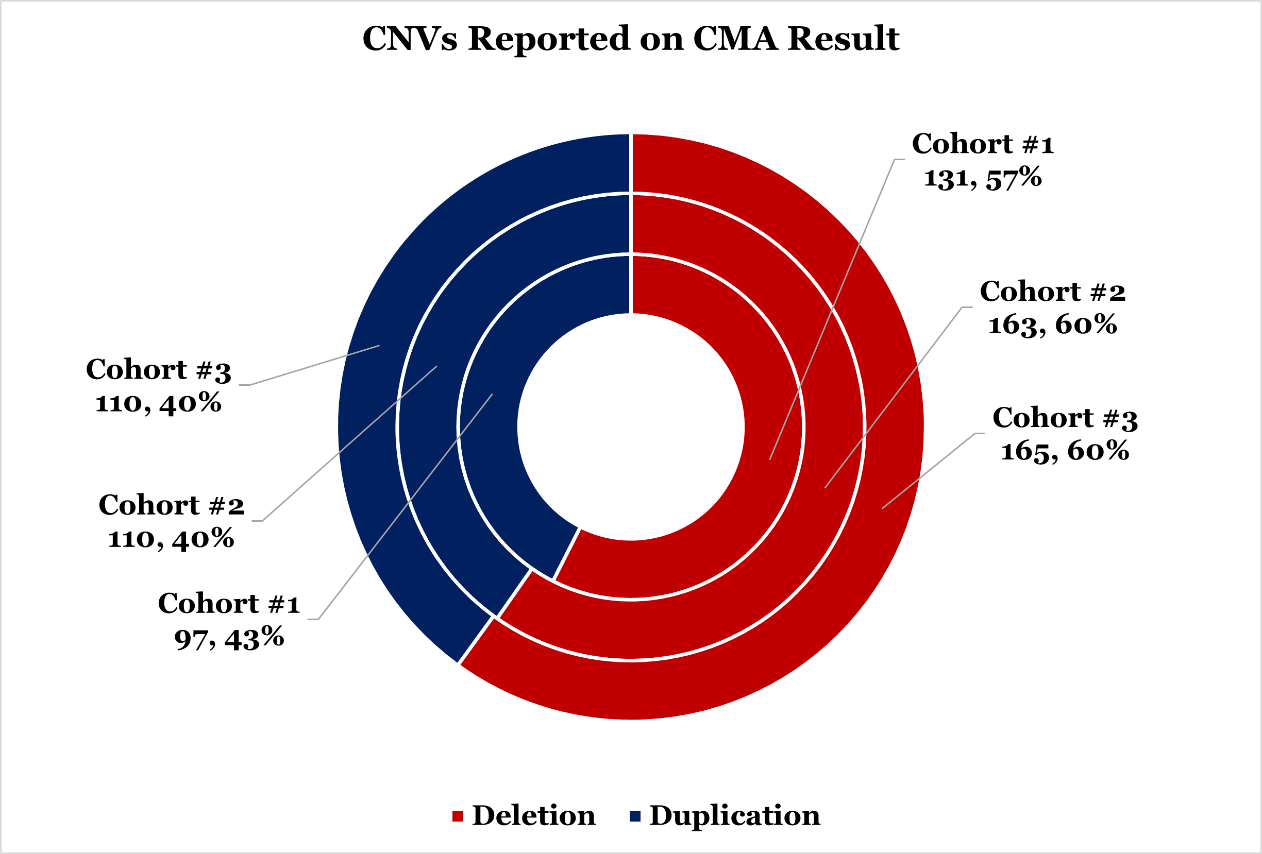

**Supplemental Figure 7: Size Distribution of CNVs**

**.** (A) Frequency of CNVs at each length bin size for all cohorts by direction (Gain: blue; Loss: red; Total: black). (B) Boxplot of Log10 Normalized CNV lengths with outliers for each direction. (C) Median CNV length with 95% confidence intervals by direction. Kruskal Wallis H-Test determined that the median CNV duplication length was significantly longer than the median CNV deletion length (χ^2^=5255.9, p<0.001).

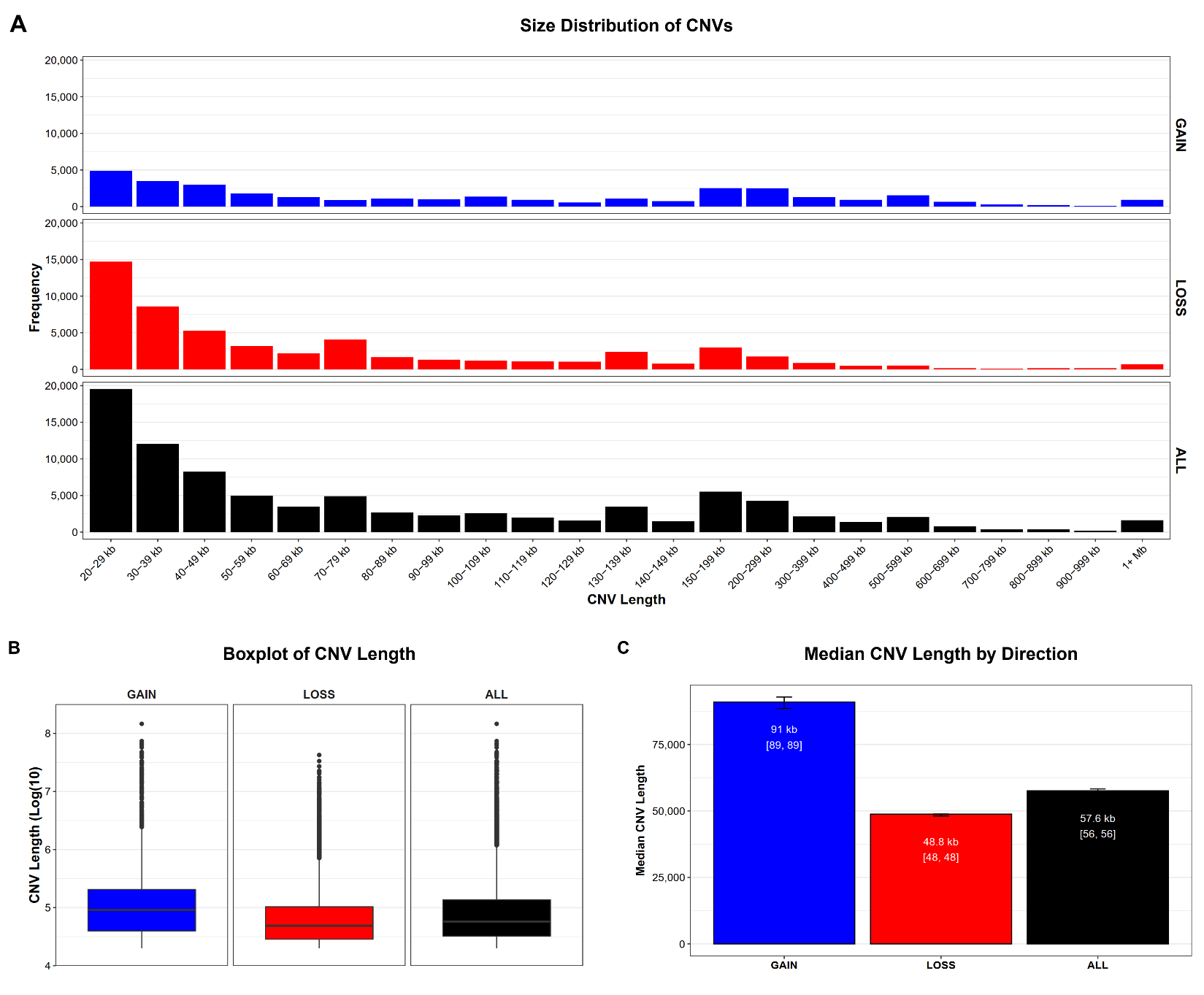

**Supplemental Figure 8: Size Distribution of CNVs by Cohort**

**.** (A) Frequency of CNVs at each length bin size for each cohort (Cohort #1: yellow; Cohort #2: blue; Cohort #3: green). (B) Frequency of CNVs at each length bin size for each cohort (deletions and duplications separate). (C) Boxplot of Log10 Normalized CNV lengths with outliers for each cohort. (D) Bar plot of median CNV length with 95% Confidence Intervals by Cohort and direction. Two Kruskal Wallis H-Tests determined that the median length of duplication (χ^2^=1365.8, p<0.001) and deletion (χ^2^=13,355, p<0.001) CNVs differed by cohort. Pairwise Wilcoxon Signed Rank Tests identified that all three cohorts were significantly different in terms of deletion length (p<0.001), and Cohort #1 was significantly different from Cohort #2 and Cohort #3 in terms of duplication length (p<0.001).

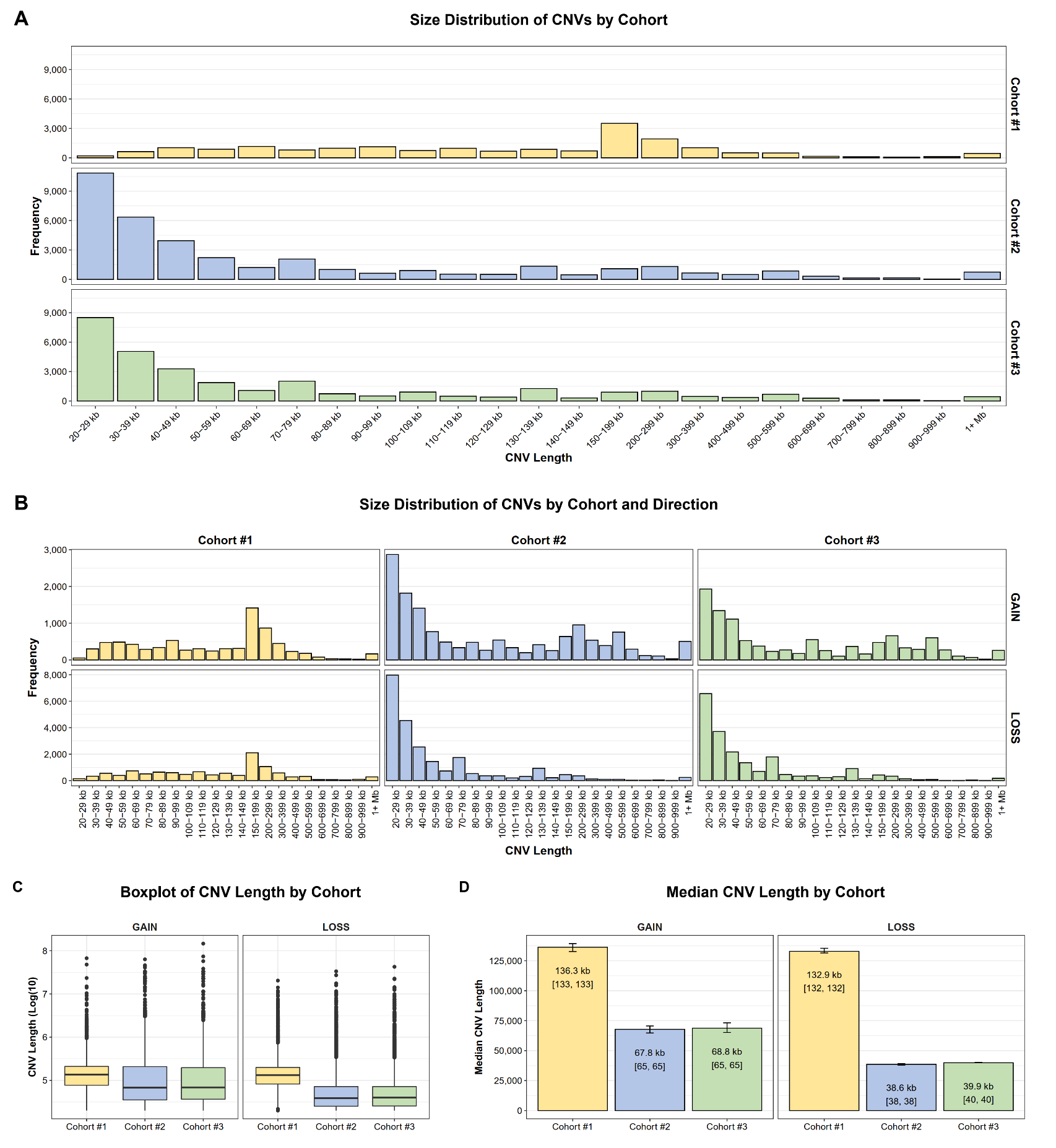

**Supplemental Figure 9: Average Number of CNVs Detected by Microarray per Case by Cohort**

**.** (A) Average number of CNVs (gains, losses, and total) identified per case by cohort (Cohort #1: yellow; Cohort #2: blue; Cohort #3: green). Error bars show standard error of the mean. (B) Box plot of the average number of CNVs identified per case by Cohort and direction. There was a significant difference in the average number of CNVs identified by direction and cohort as assessed by Kruskal-Wallis rank sum test with Benjamini & Hochberg multiple-test correction (Gain: p<0.001; Loss: p<0.001; Total: p<0.001).

A

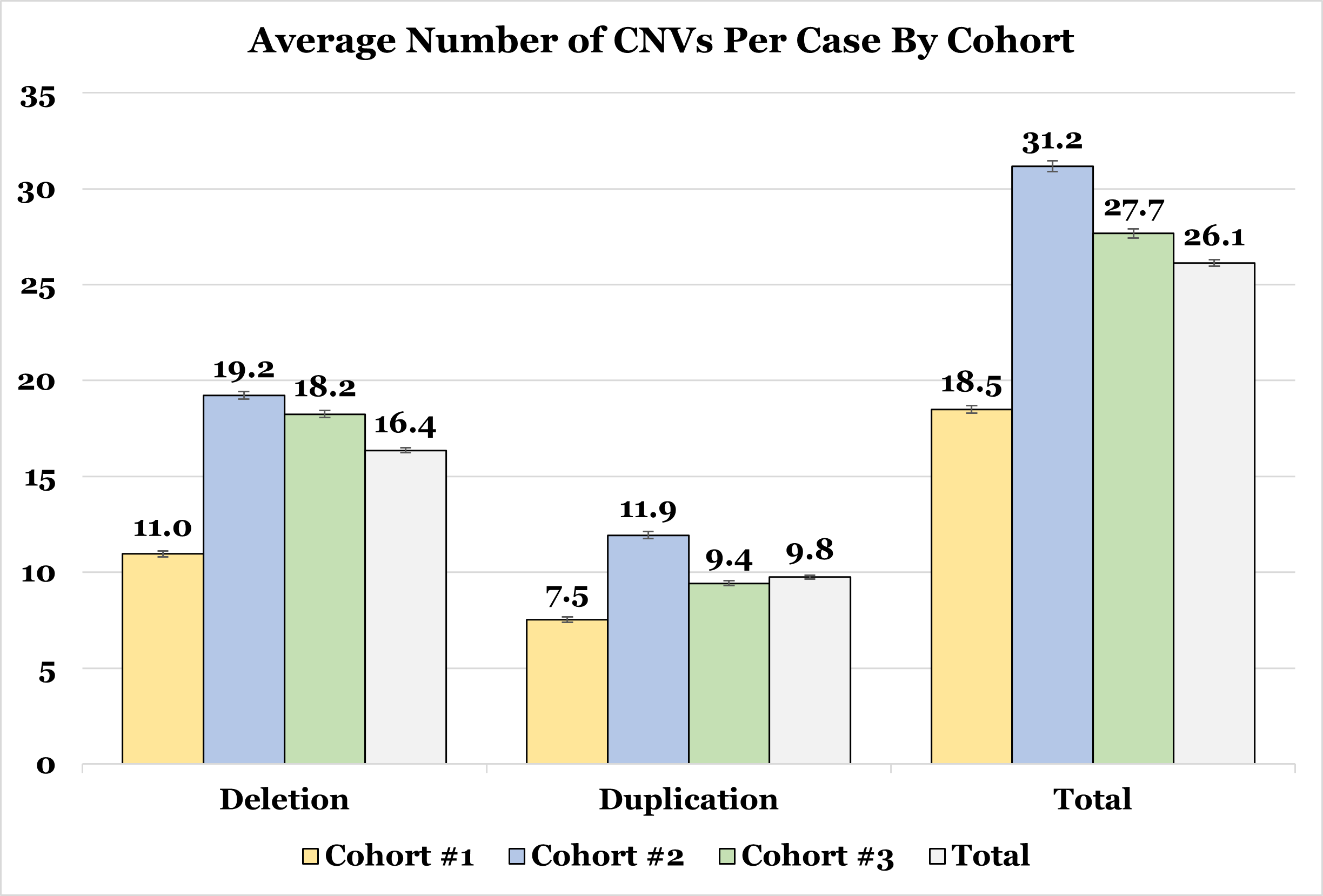

B

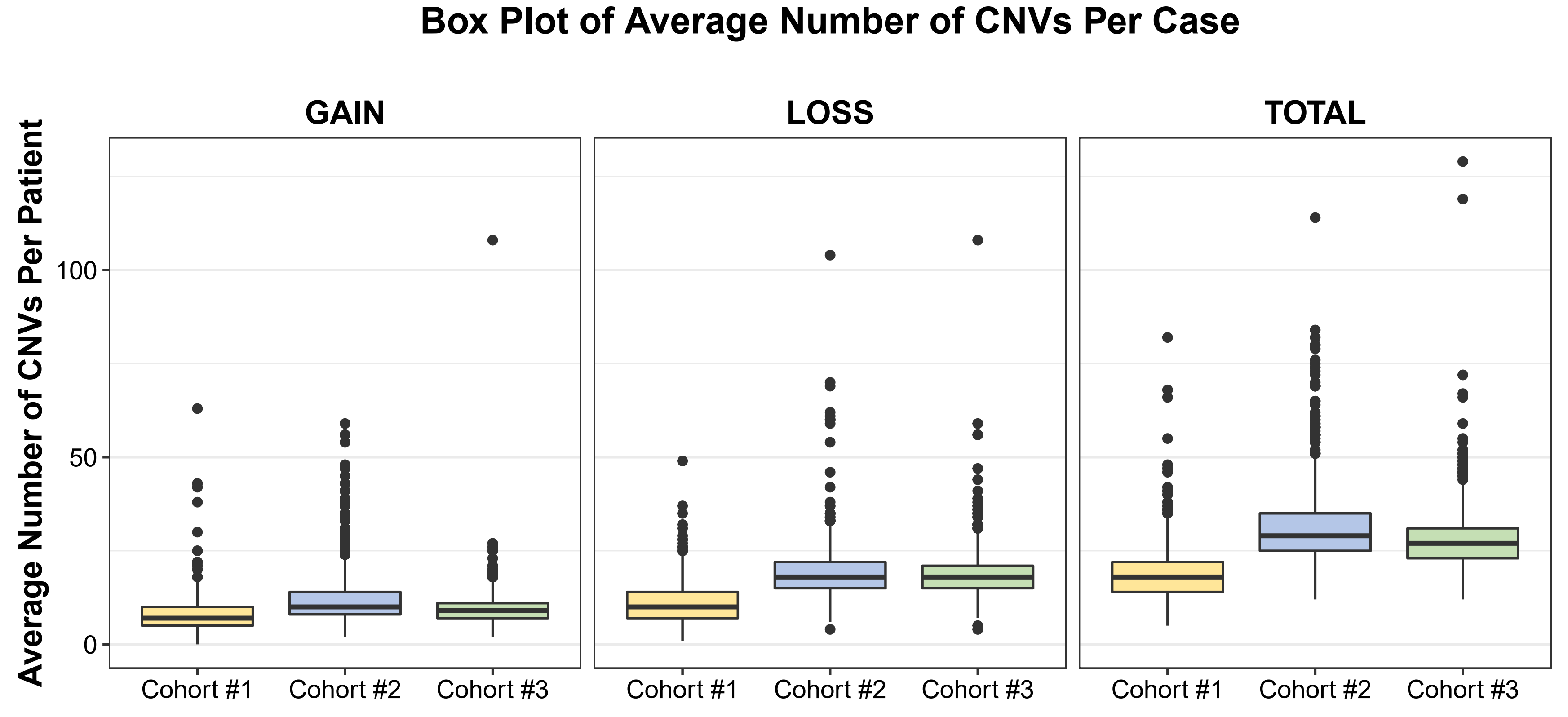

**Supplemental Figure 10: Total number of Primary CNVs Identified by DISCRIMINATOR for Each Jaccard Value**

**.** The total number of Primary CNVs identified for each cohort at each Jaccard Threshold (Cohort #1: yellow; Cohort #2: blue; Cohort #3: green).

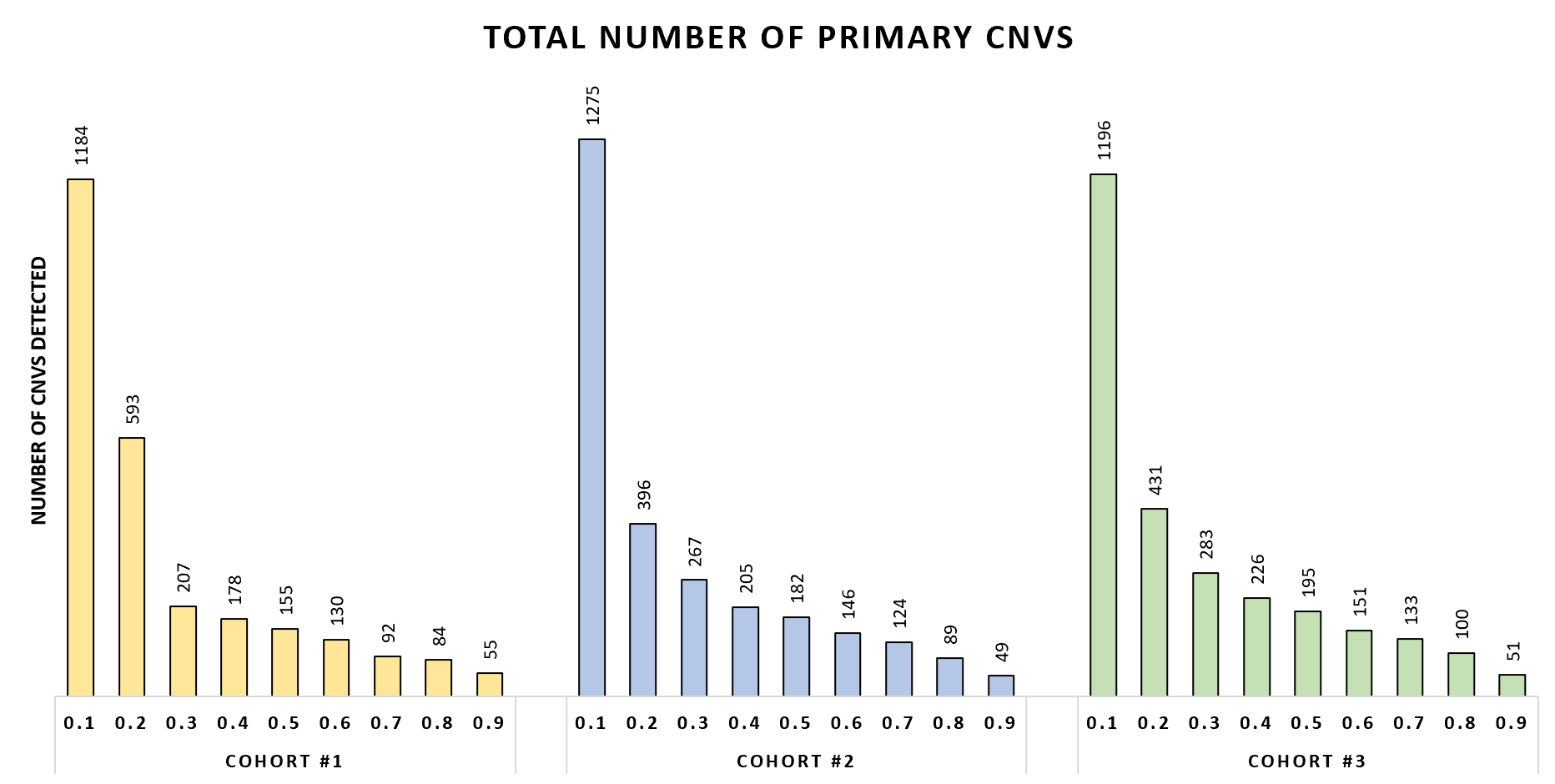

**Supplemental Figure 11: Number of Primary CNVs Identified by DISCRIMINATOR for Each Jaccard Value (0.3-0.5)**

**.** For each cohort and Primary CNV locus evaluated, the number of Primary CNVs detected was compared to the actual number of CNVs for each Primary CNVs within that locus. A colored box indicates that at that Jaccard Threshold, all Primary CNV intervals within the region are being called correctly. CNVs identified in each cohort are color coded (Cohort #1: yellow, Cohort #2: blue, and Cohort #3: green). Across all cohorts, and all evaluated Primary CNVs, the optimal Jaccard Threshold value was: 0.40-0.41.

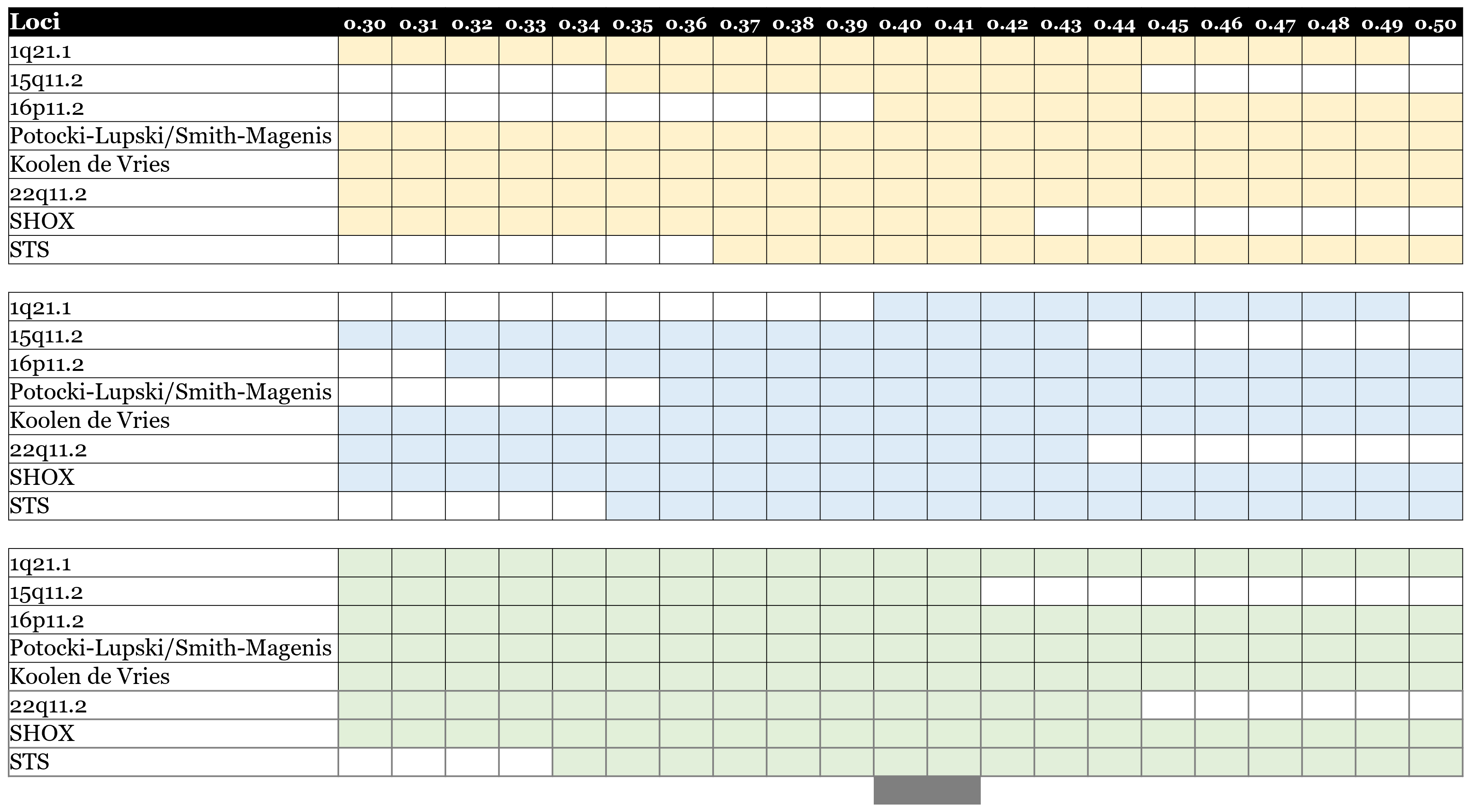

**Supplemental Figure 12: Size Distribution of Provisional CNV Classifications**

**.** (A) Frequency of CNVs at each length bin size for each provisional classification (Primary: red; Secondary: yellow; Benign: green; Non-Coding: grey). (B) Frequency of CNVs at each length bin size for each provisional classification and direction. (B) Frequency of CNVs at each length bin size for each provisional classification and cohort.

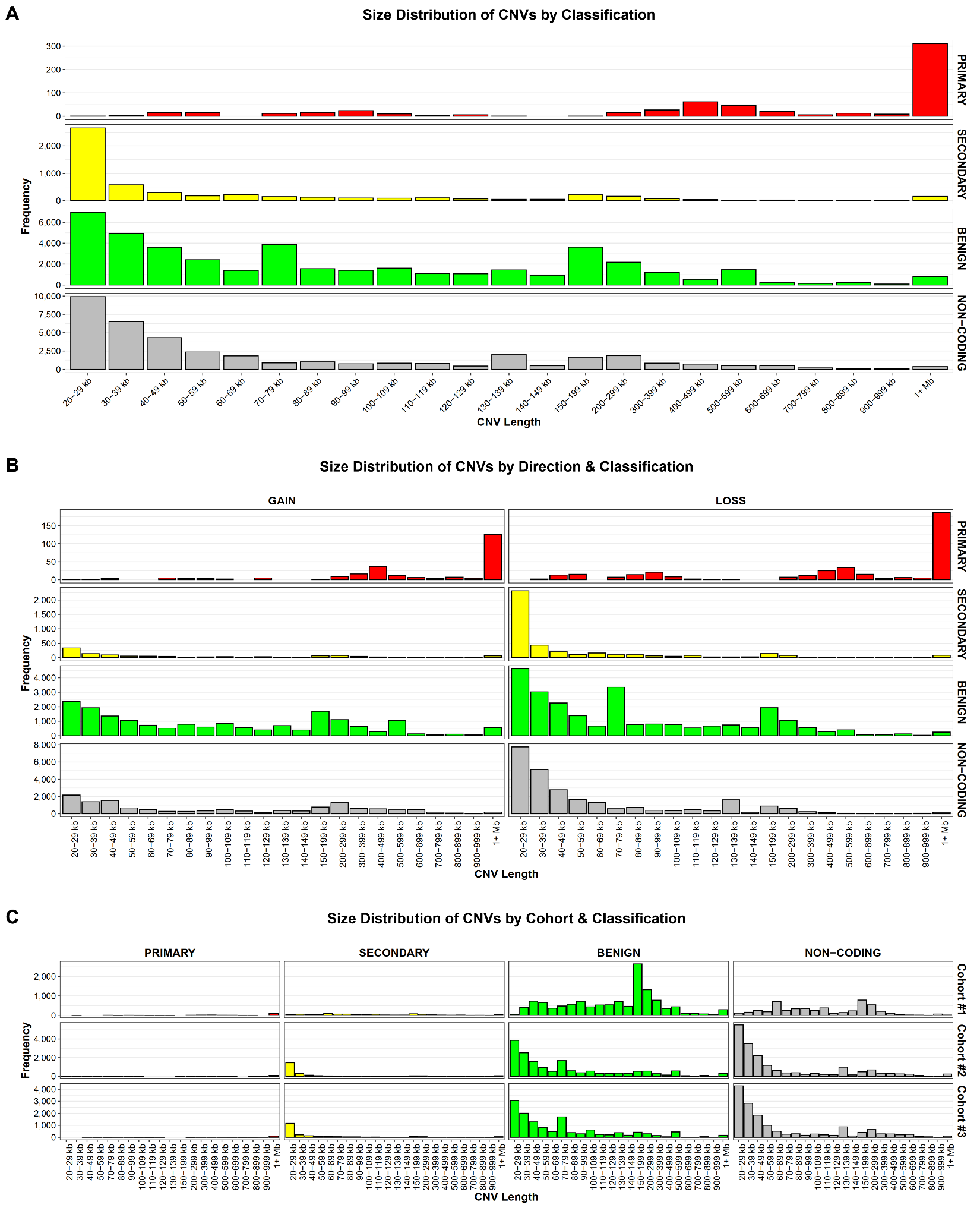

**Supplemental Figure 13: Average Number of CNVs Per Case By Classification By Cohort**

A

**.** (A) The plot displays the average number of deletions, duplications, and total CNVs per case broken down by provisional classification in each cohort, and cohort wide (Cohort #1: yellow; Cohort #2: blue; Cohort #3: green; Cohort-Wide: grey). Error bars show standard error of the mean. (B) Box plot of the average number CNVs identified per case broken down by classification and cohort. There was no significant difference in the average number of identified Primary CNVs between cohorts as assessed by Kruskal-Wallis rank sum test (p=0.06442). There was a significant difference in the average number of identified Secondary, Benign, and Non-Coding CNVs between cohorts (Secondary: p<0.001; Benign: p<0.001; Non-Coding: p<0.001).

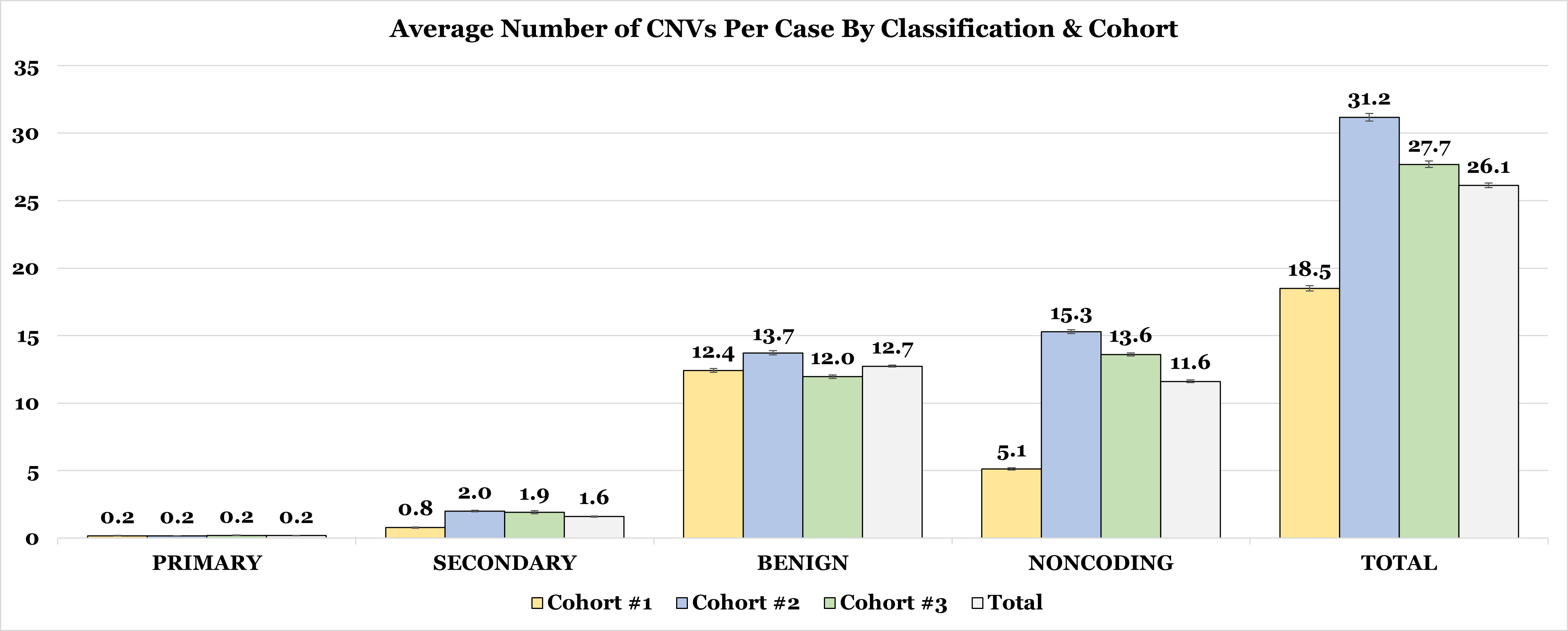

B

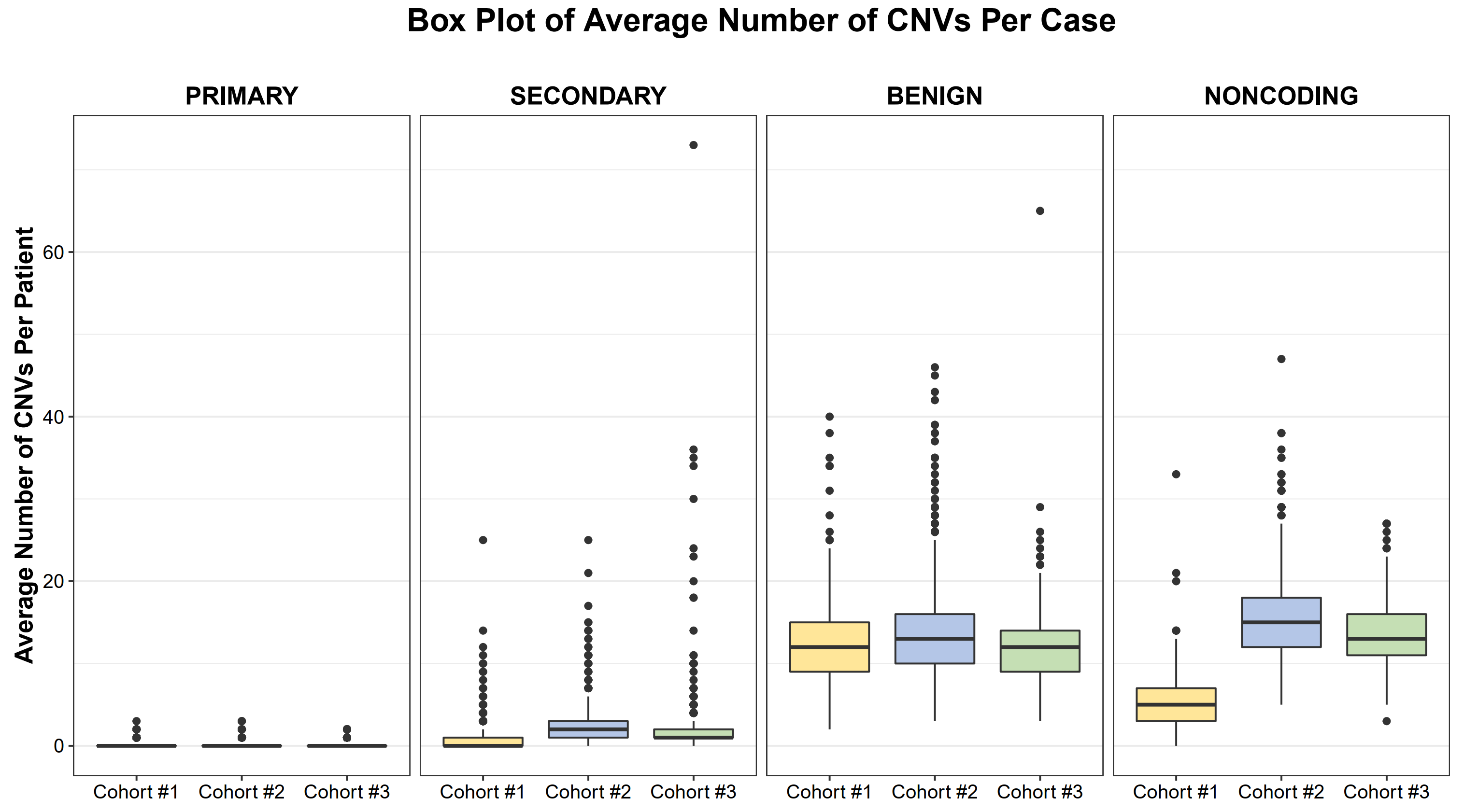

**Supplemental Figure 14: Comparison of Affymetrix CytoscanHD Quality Score ('Noise') Between Cohorts**

**.** (A) Box plot of quality scores for each cohort with the quality score value plotted for each individual clinical CMA. (B) Dot plot of individual CMA quality scores plotted over time. Quality score values are color coded by cohort (Cohort #2: blue, and Cohort #3: green). There was a significant difference in the average quality score value between cohorts as assessed by Kruskal-Wallis rank sum test (p<0.001).

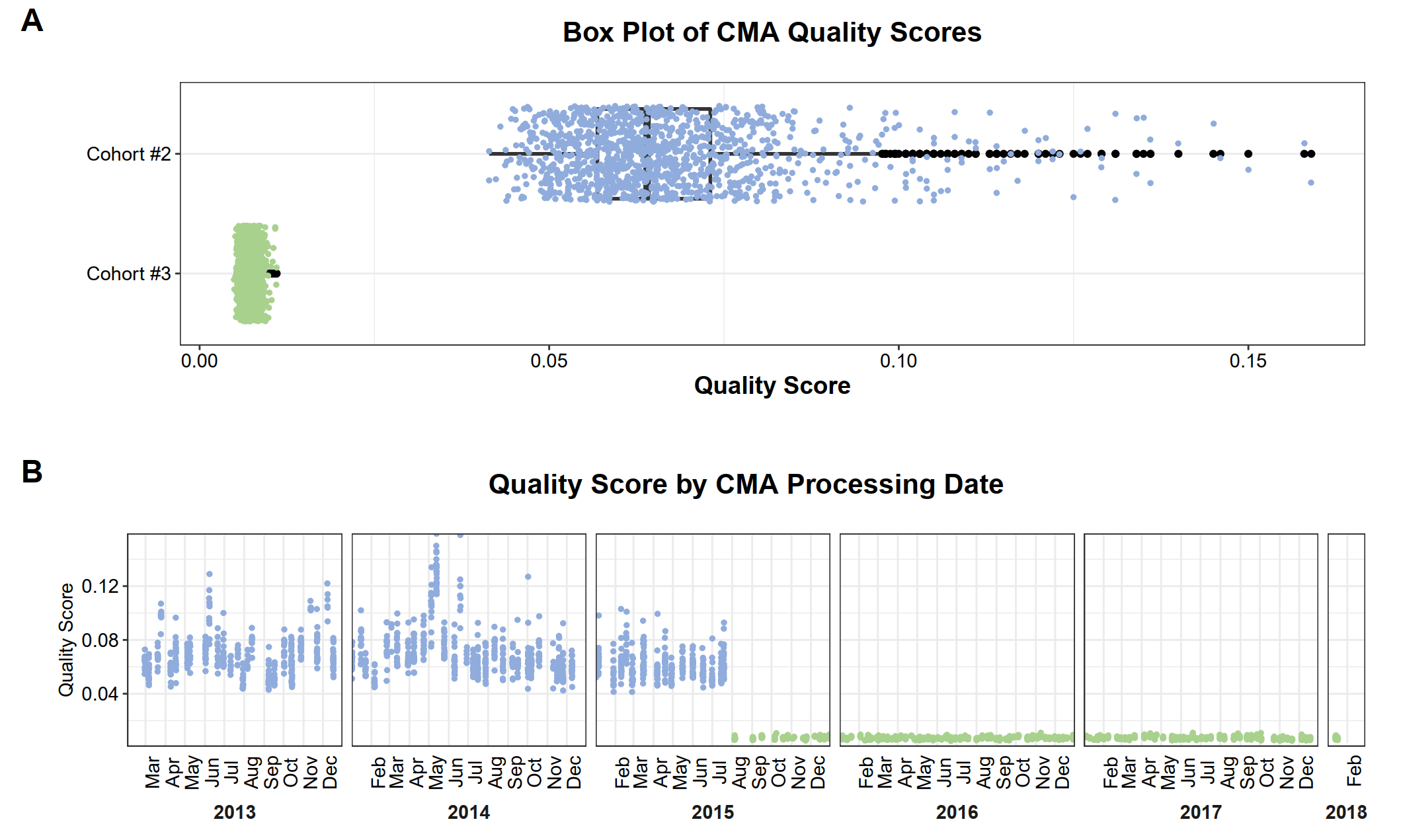

**Supplemental Figure 15: 22q11.2 Region with Segmented Primary CNV Calls**

**.** UCSC Genome Browser View (http://genome.ucsc.edu) of the 22q11.2 region. From top to bottom, tracks shown represent individual sample level calls from cases with segmented 22q11.2 A:D CNVs, chromosome banding pattern, segmental duplications, and DISCRIMINATOR Primary CNVs. Blue highlighted regions represent the segmental duplication clusters/breakpoints which mediate rearrangement and are labeled from A-G.

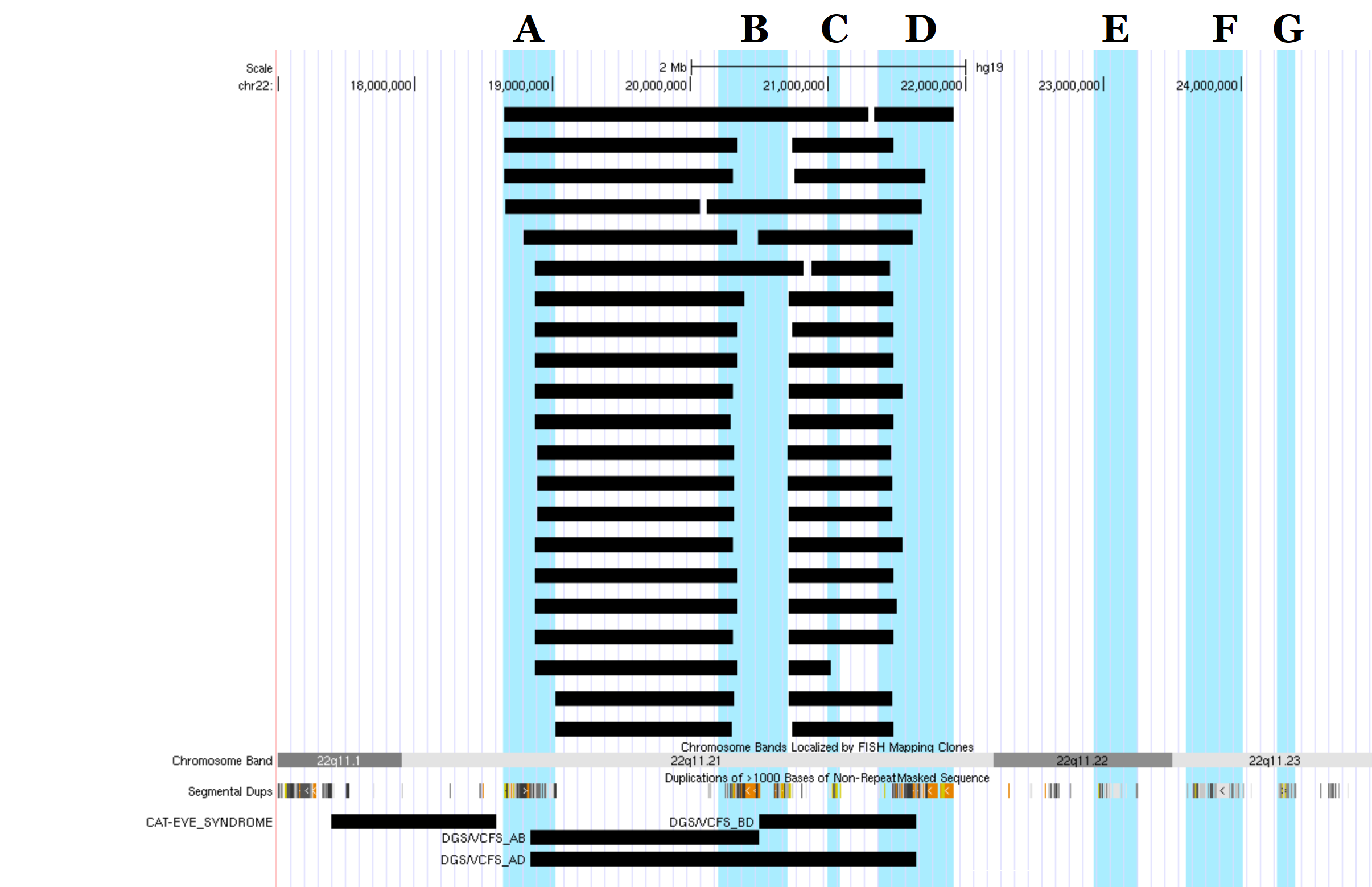

**Supplemental Figure 16: Relaxed Merging Criteria for Specified Regions**

**.**

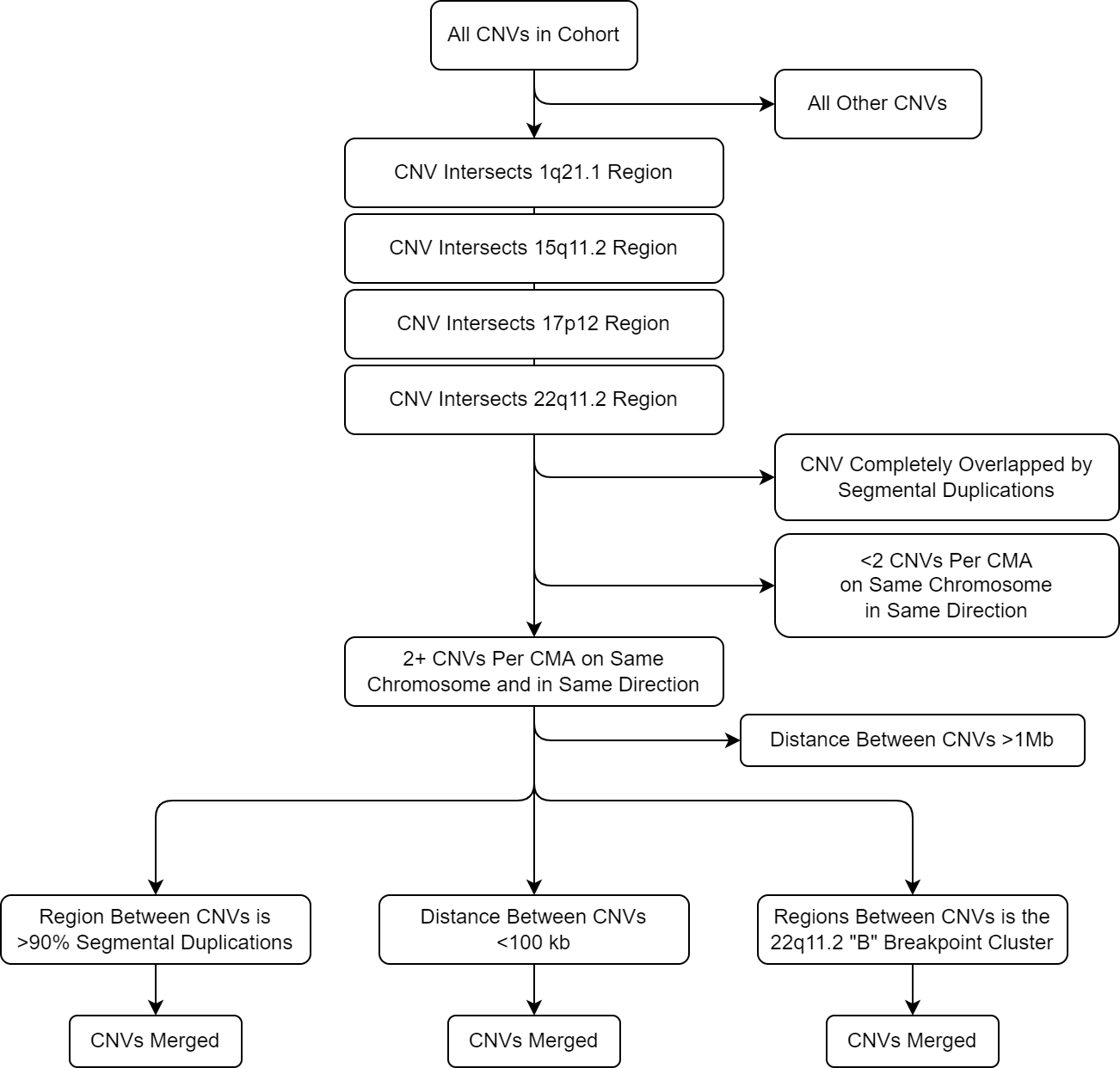

**Supplemental Figure 17: DISCRIMINATOR Code Structure**

**.** DISCRIMINATOR reads the BENIGN_FILES, PRIMARY_FILES, COHORT_FILES, and JACCARD_INDEX files to determine which cohort files to process, which files contain the benign and pathogenic region intervals, and determine the user-defined Jaccard Index Threshold(s). The set of supplied benign interval files are merged, and each cohort file undergoes a merging step. The DISCRIMINATOR_ALG.py and STATISTICS_AND_OUTPUT.py scripts are run for each [ Jaccard Index * Primary Interval file * Cohort file ] combination. Each combination will yield two output files: COMPLETE_OUTPUT and COLLAPSED_ANNATIONS. DISCRIMINATOR will also create a merged benign file for the set of benign intervals and a merged cohort file for all cohort files supplied.

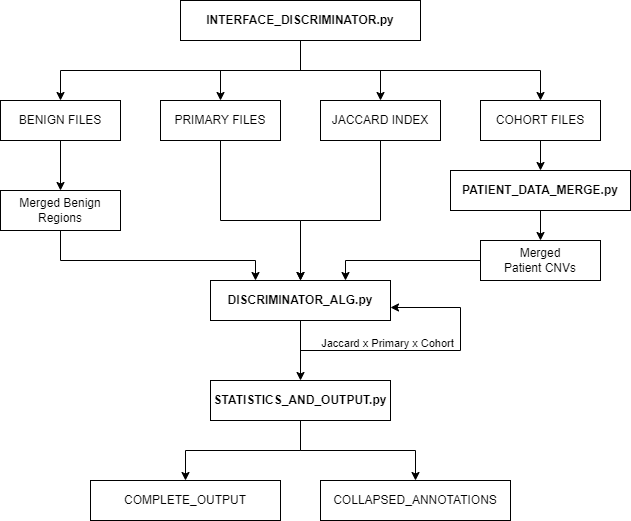

**Supplemental Figure 18: Number of Primary CNVs Identified by DISCRIMINATOR for Each Jaccard Value (1q21.1)**

**.** (A) UCSC Genome Browser View (http://genome.ucsc.edu) of the 1q21.1 region (chr1:144,000,000-149,000,000; hg19). The different DISCRIMINATOR regions for the 1q21.1 TAR, 1q21.1 Neuro, and 1q21.1 TAR Neuro CNV Intervals are shown as black bars, genes in this region are shown by dark blue bars, and the breakpoint regions are highlighted in blue. (B) The number of Primary CNVs called within each cohort for each Jaccard threshold value (0-1-0.9) and Primary CNV (1q21.1 TAR, 1q21.1 Neuro, and 1q21.1 TAR Neuro) for gain events. The true number of primary CNVs is depicted on the far right of each graph. CNVs identified in each cohort are color coded (Cohort #1: yellow, Cohort #2: blue, and Cohort #3: green). (C) The number of Primary CNVs called within each cohort for each Jaccard threshold value (0-1-0.9) and Primary CNV (1q21.1 TAR, 1q21.1 Neuro, and 1q21.1 TAR Neuro) for loss events. (D) The number of Primary CNVs called within each cohort for each Jaccard threshold value (0.3-0.5) for the 1q21.1 TAR Duplication. The number of Primary CNVs identified for remainder of the Primary CNVs in the 1q21.1 region was unchanged across the 0.3-0.5 Jaccard threshold range.

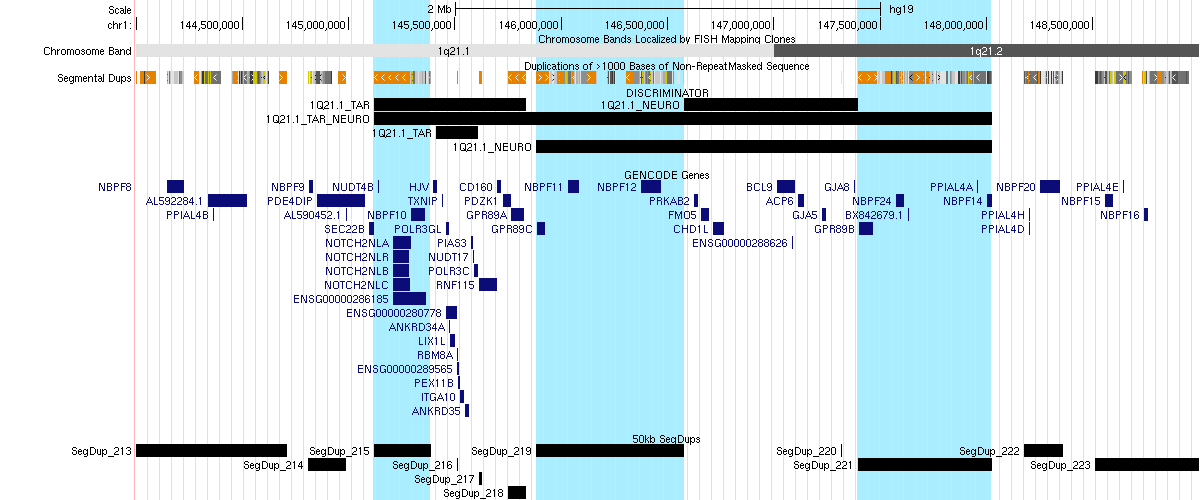

A

B

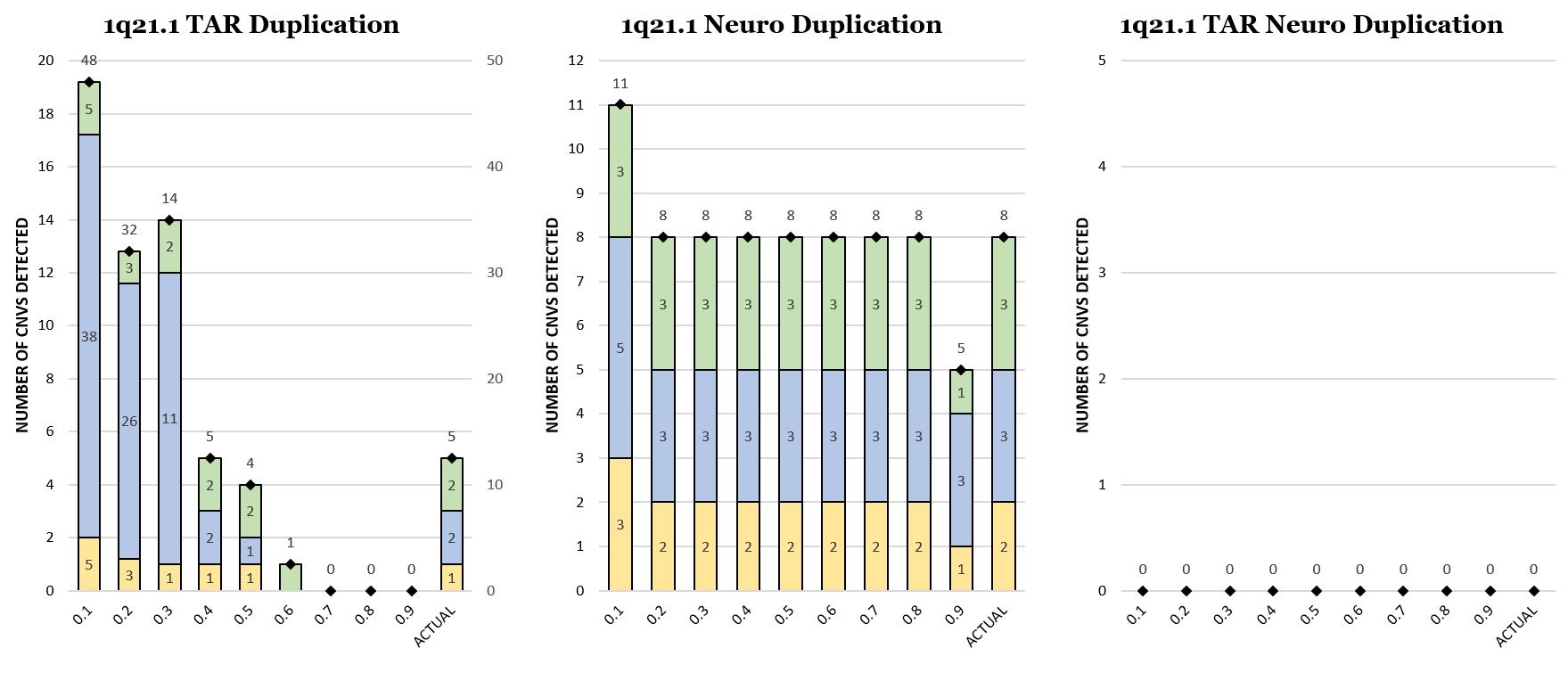

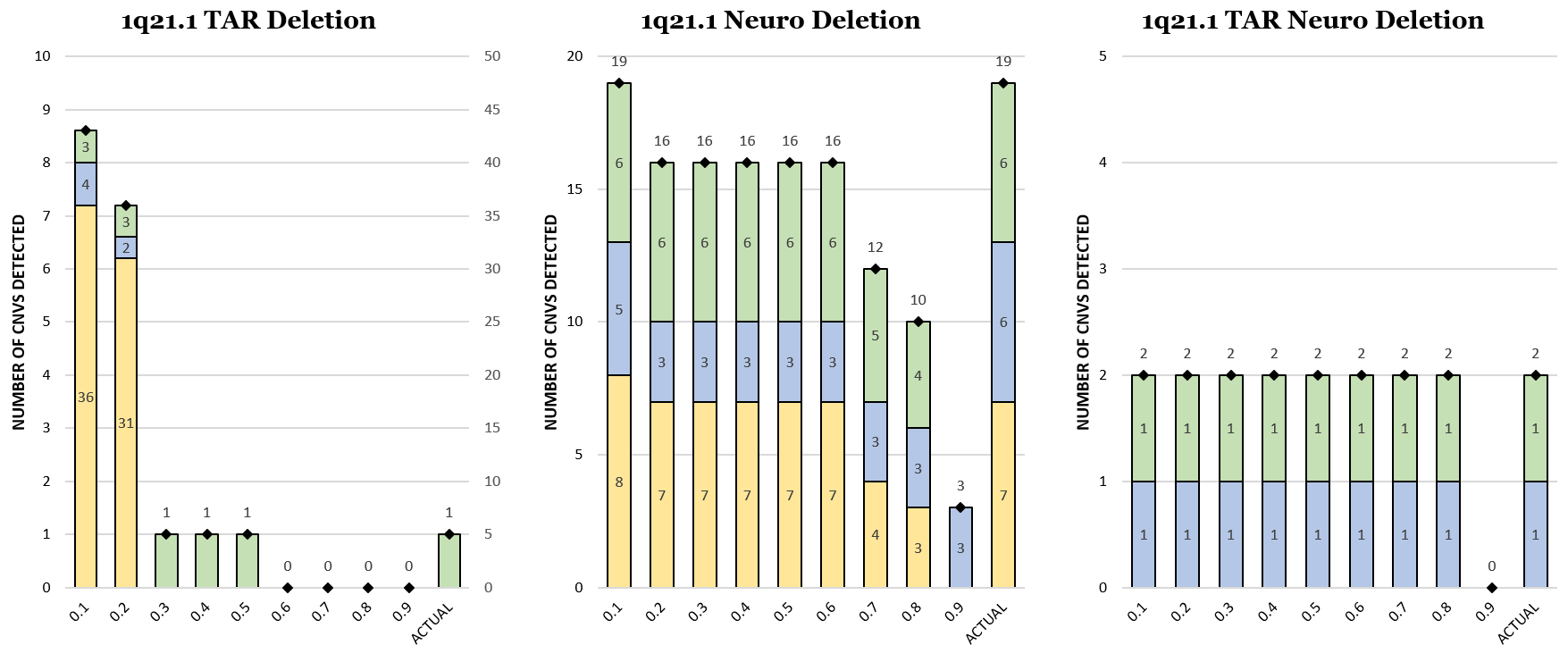

C

D

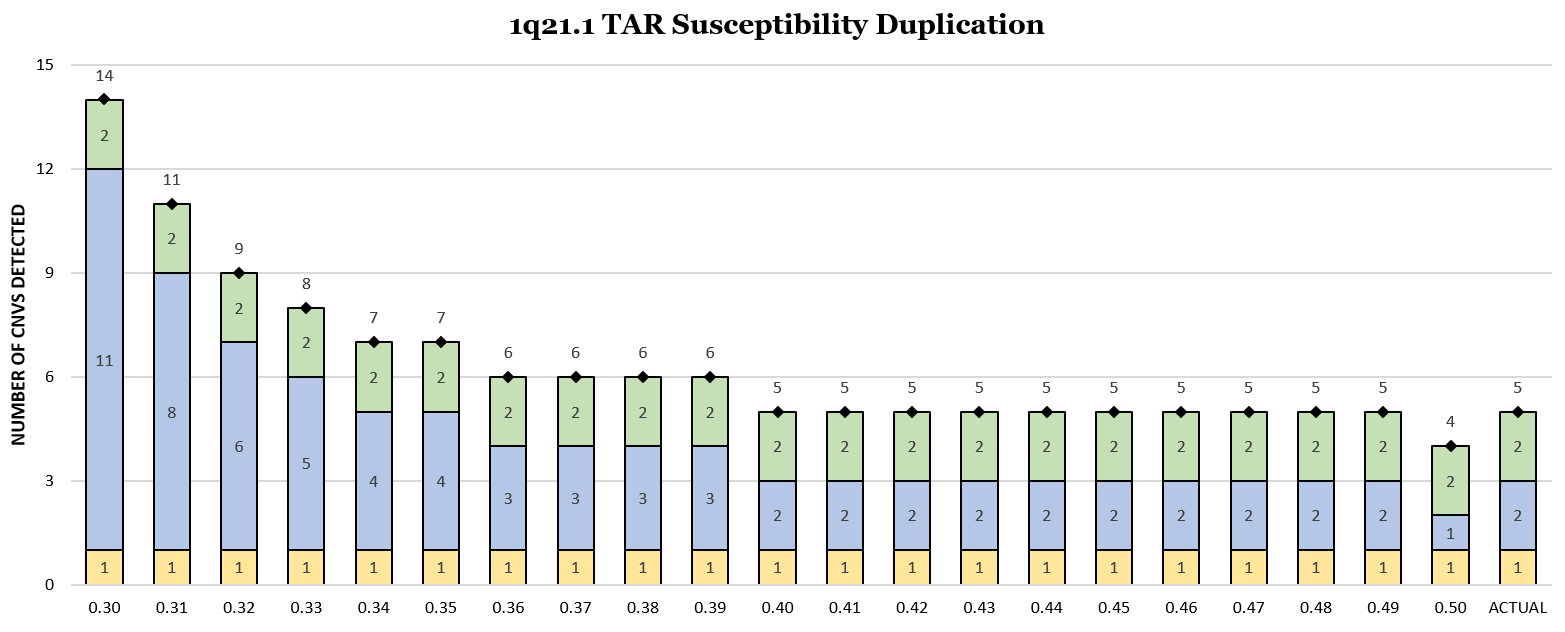

**Supplemental Figure 19: Number of Primary CNVs Identified by DISCRIMINATOR for Each Jaccard Value (****15q11.2)**

**.** (A) UCSC Genome Browser View (http://genome.ucsc.edu) of the 15q11.2 region (chr15:20,000,000-34,000,000; hg19). The different DISCRIMINATOR regions for the 15q11.2 Primary CNV Intervals are shown as black bars, genes in this region are shown by dark blue bars, and the breakpoint regions are labeled and highlighted in blue. (B) The number of Primary CNVs called within each cohort for each Jaccard threshold value (0-1-0.9) and Primary CNV within the 15q11.2 locus for gain events. The true number of primary CNVs is depicted on the far right of each graph. CNVs identified in each cohort are color coded (Cohort #1: yellow, Cohort #2: blue, and Cohort #3: green). (C) The number of Primary CNVs called within each cohort for each Jaccard threshold value (0-1-0.9) and Primary CNV within the 15q11.2 locus for loss events. (D) The number of Primary CNVs called within each cohort for each Jaccard threshold value (0.3-0.5) for the 15q11.2 BP1-2 Duplication, 15q11.2 BP1-2 Deletion, and 15q11.2 BP3-4 Deletion. The number of Primary CNVs identified for remainder of the Primary CNVs in the 15q11.2 region was unchanged across the 0.3-0.5 Jaccard threshold range.

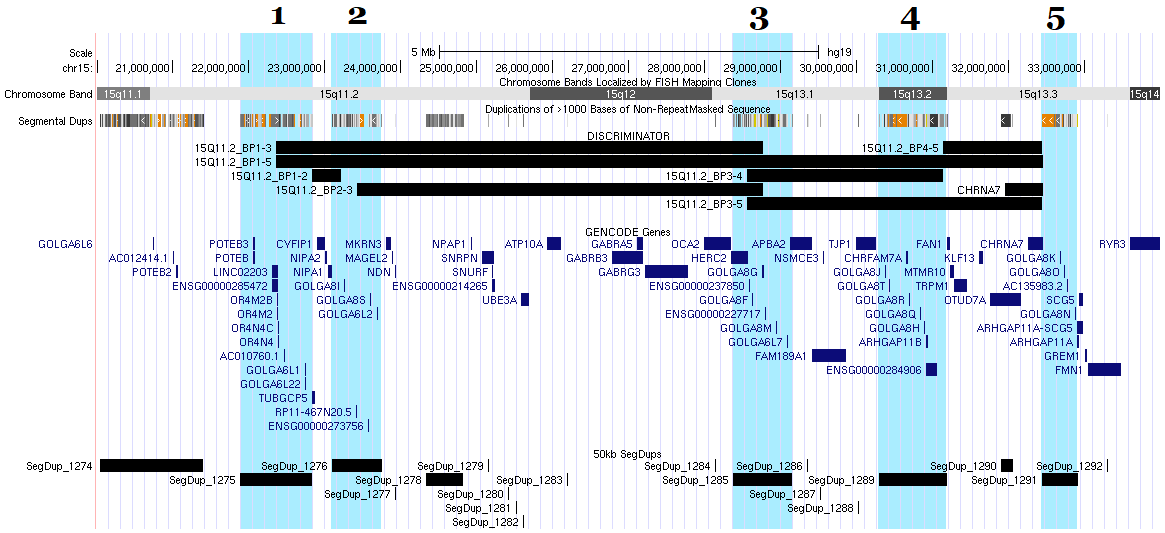

A

B

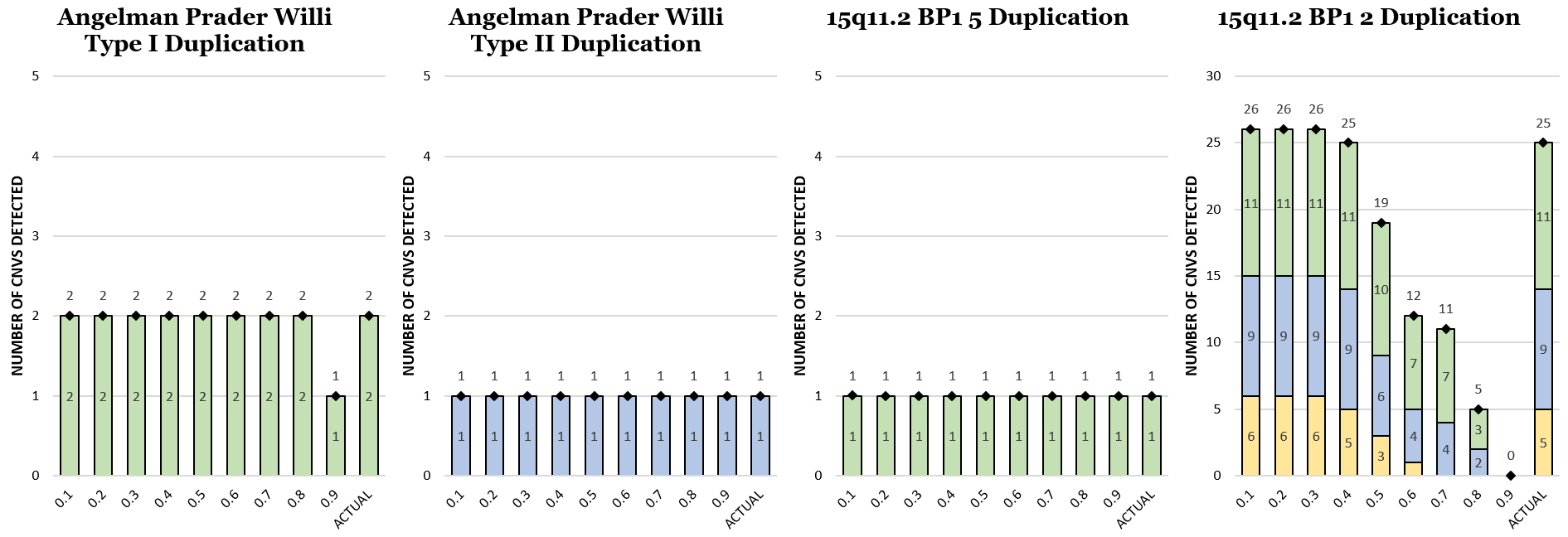

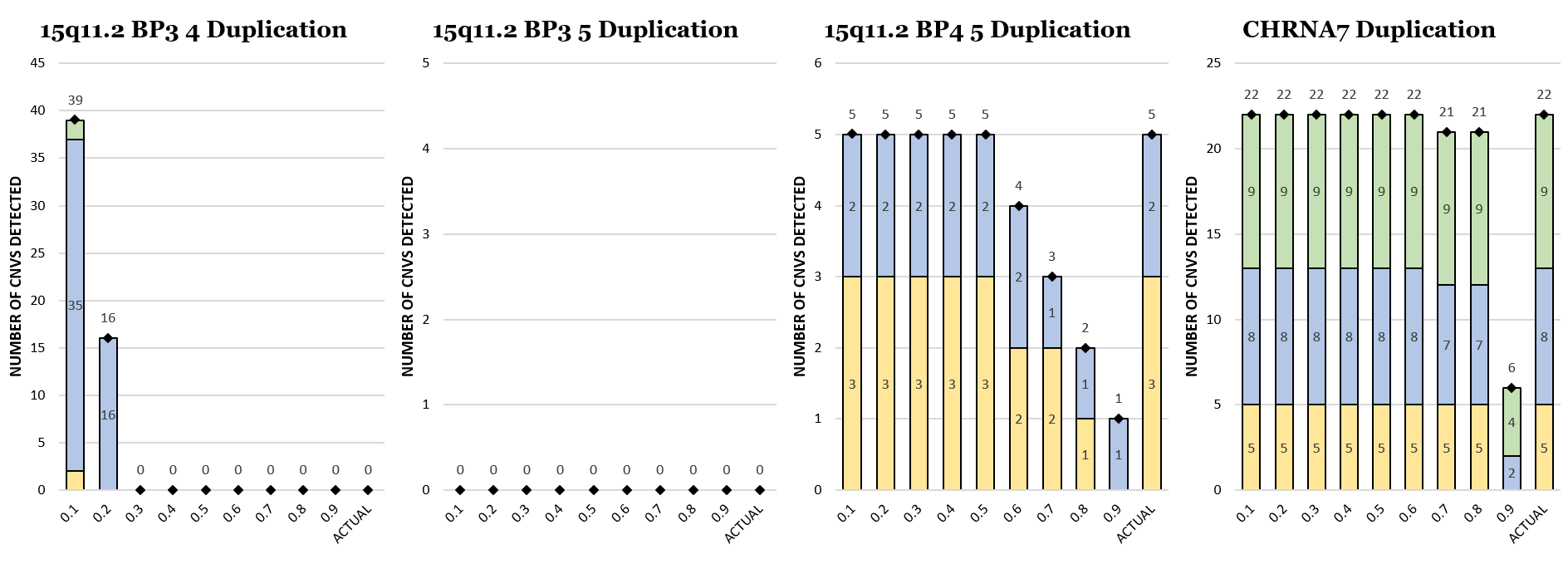

B

C

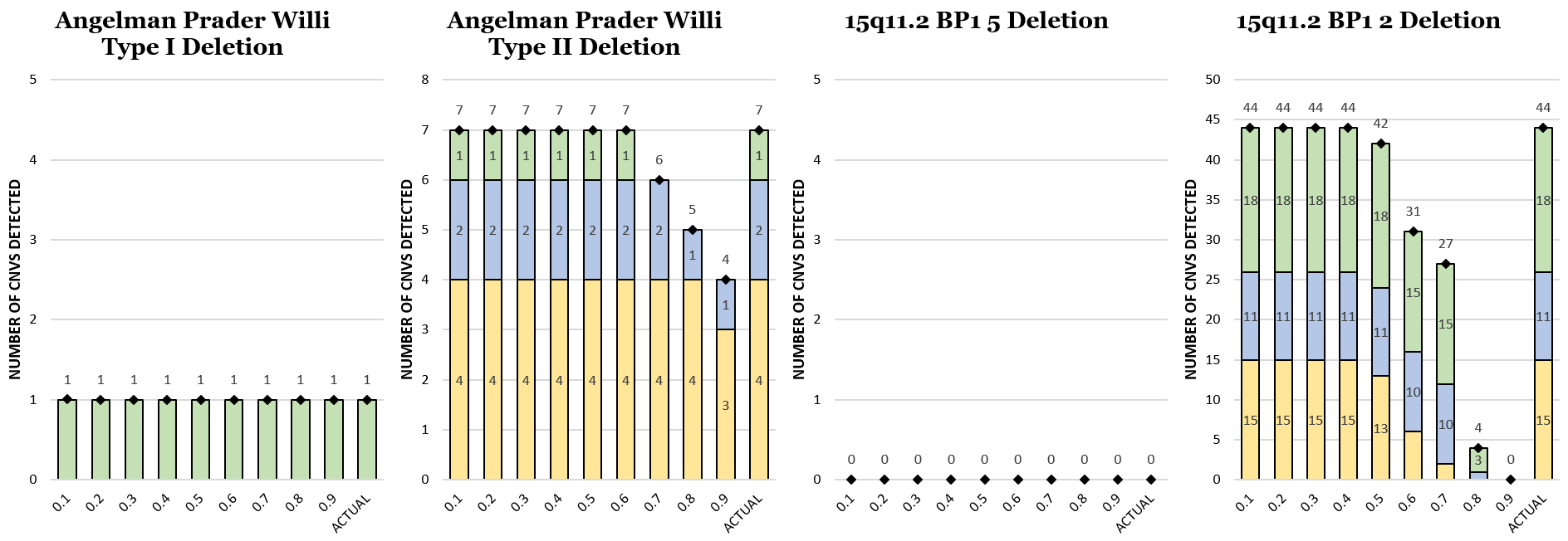

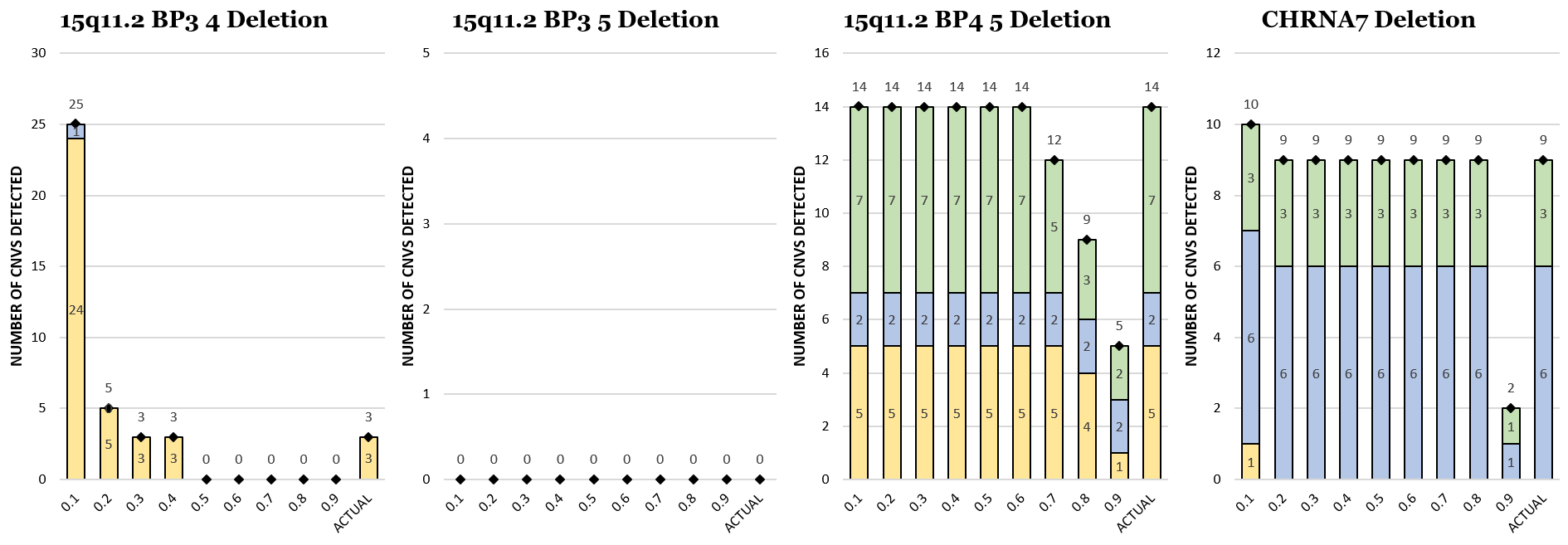

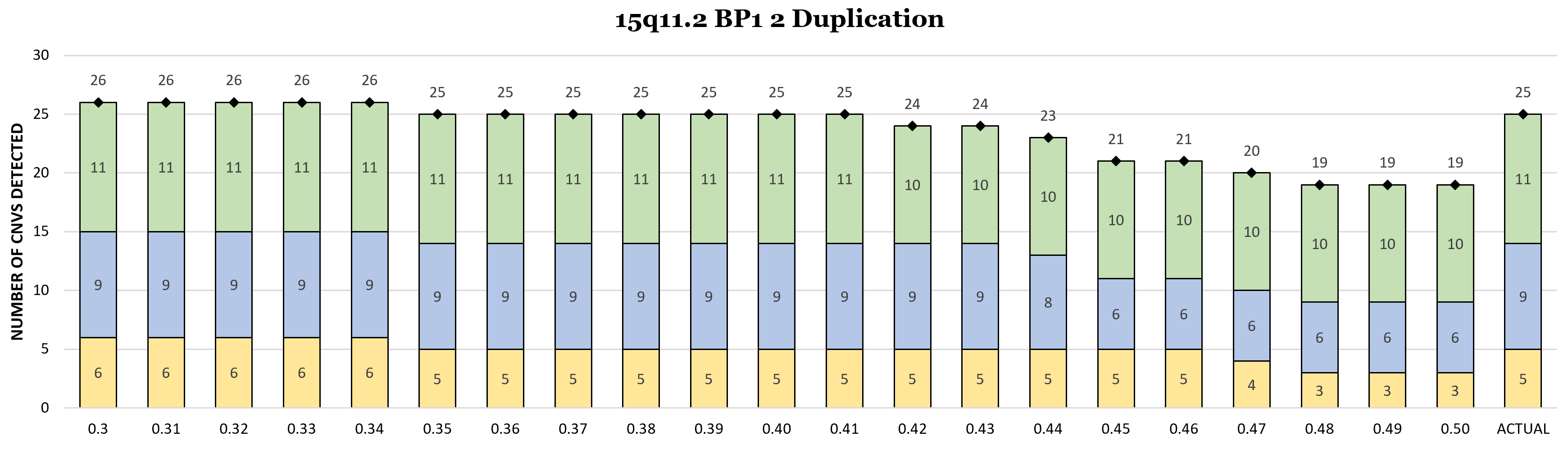

D

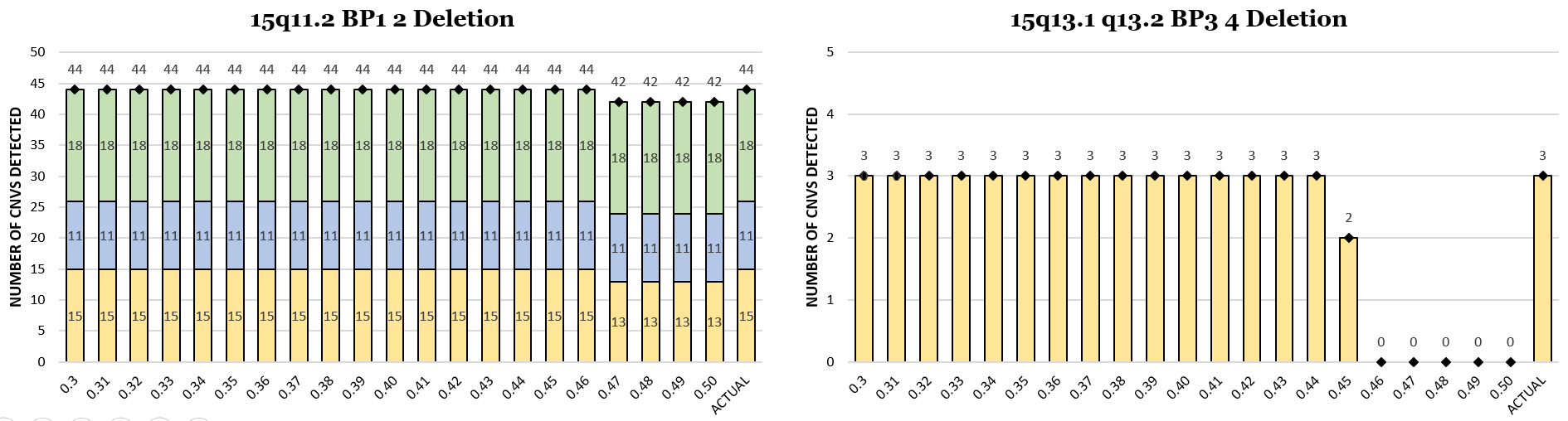

**Supplemental Figure 20: Number of Primary CNVs Identified by DISCRIMINATOR for Each Jaccard Value (16p11.2)**

**.** (A) UCSC Genome Browser View (http://genome.ucsc.edu) of the 16p11.2 region (chr16:28,500,000-30,500,000; hg19). The different DISCRIMINATOR regions for the 16p11.2 Primary CNV Intervals are shown as black bars, genes in this region are shown by dark blue bars, and the breakpoint regions are labeled and highlighted in blue. (B) The number of Primary CNVs called within each cohort for each Jaccard threshold value (0-1-0.9) and Primary CNV within the 16p11.2 locus for both gain and loss events. The true number of primary CNVs is depicted on the far right of each graph. CNVs identified in each cohort are color coded (Cohort #1: yellow, Cohort #2: blue, and Cohort #3: green). (C) The number of Primary CNVs called within each cohort for each Jaccard threshold value (0.3-0.5) for the 16p11.2 Duplication and Deletion.

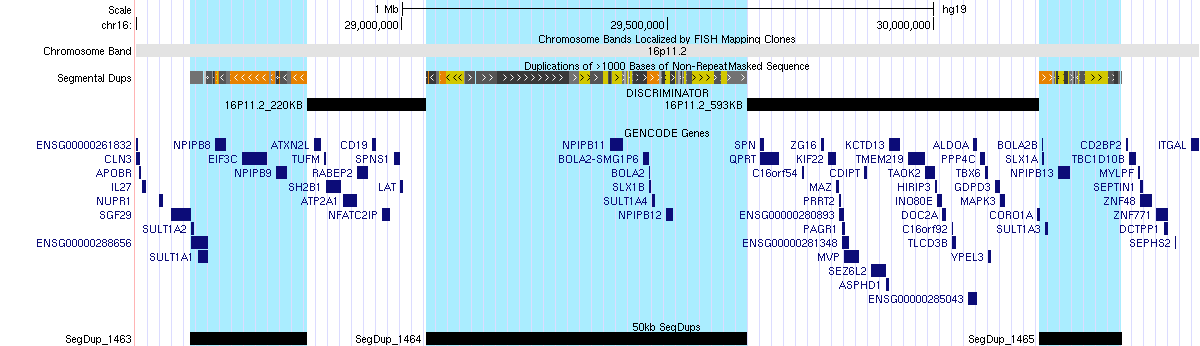

A

B

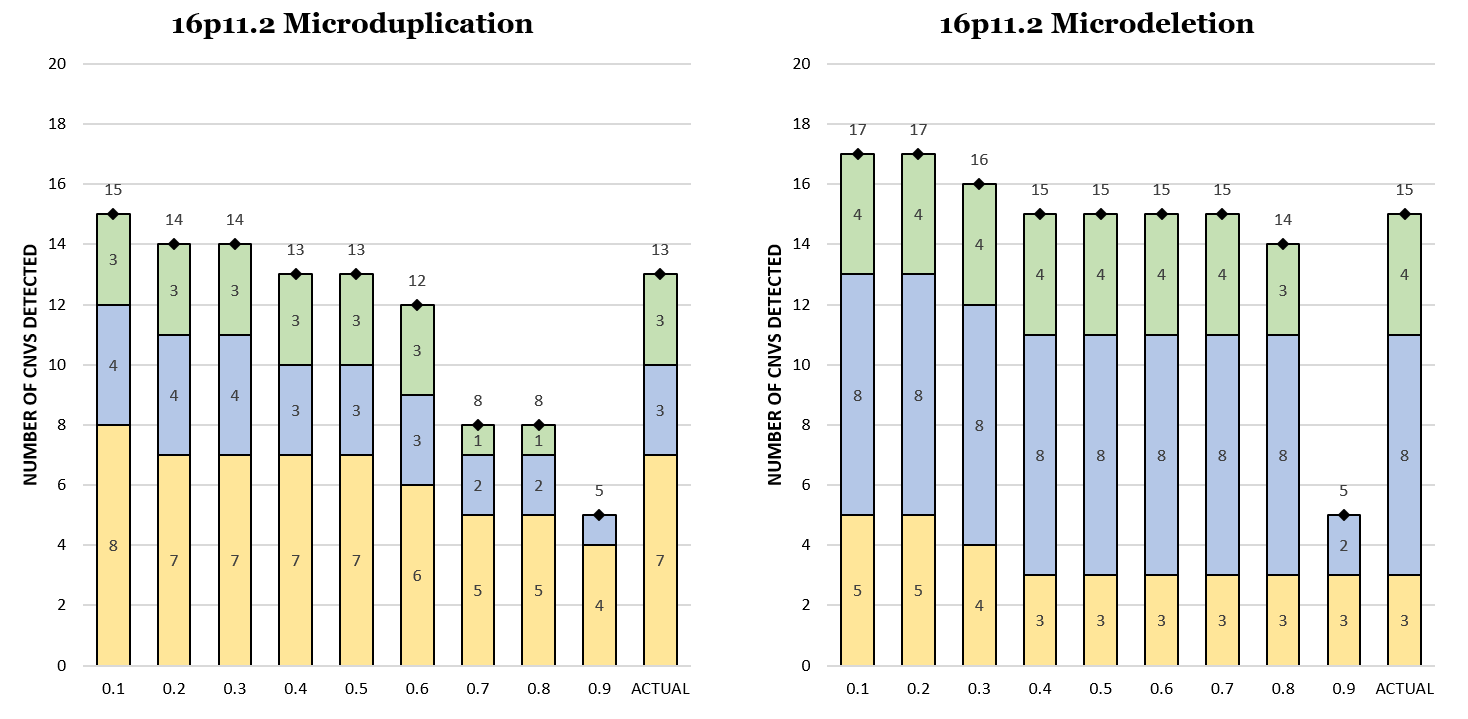

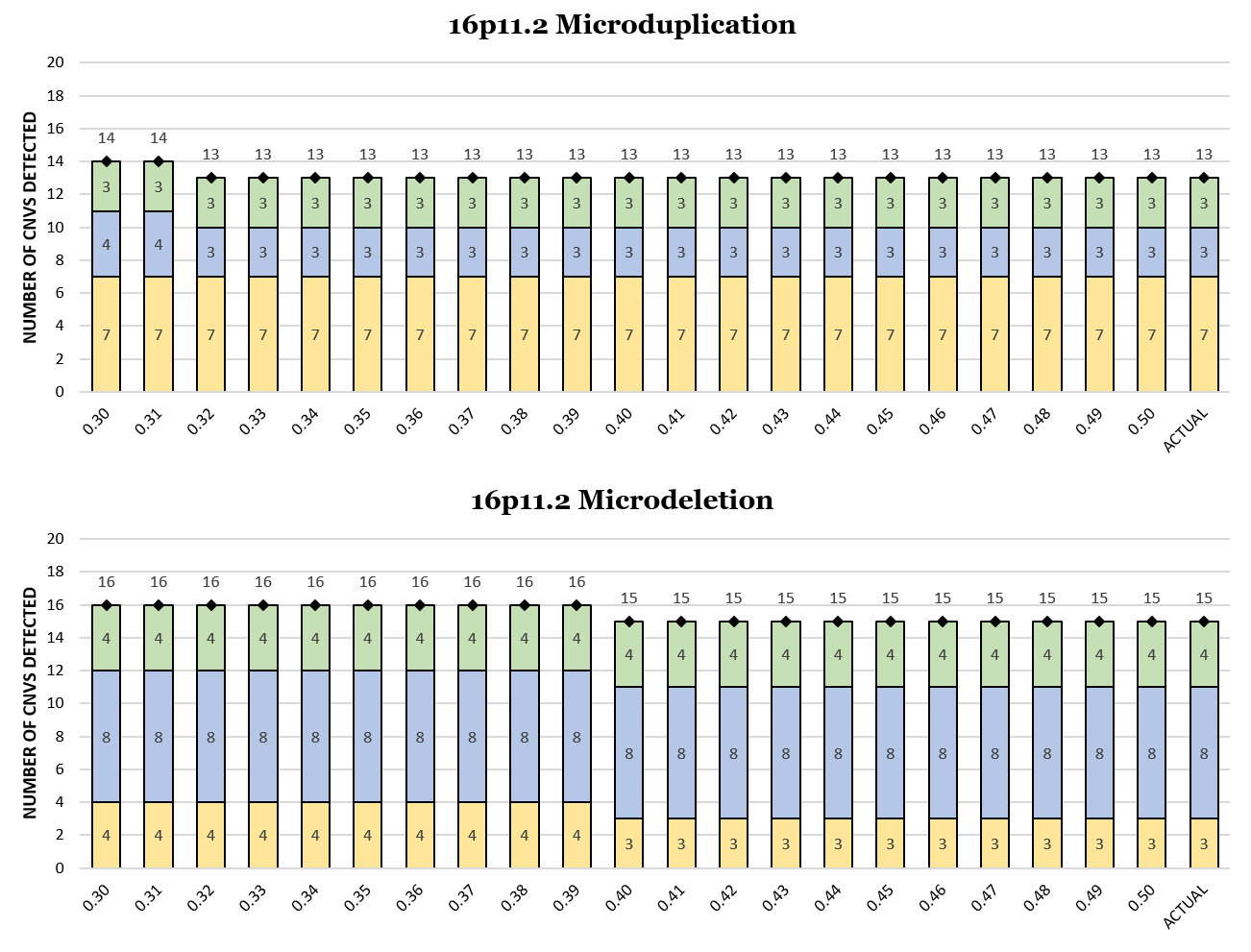

C

**Supplemental Figure 21: Number of Primary CNVs Identified by DISCRIMINATOR for Each Jaccard Value (17p12-p11.2)**

**.** (A) UCSC Genome Browser View (http://genome.ucsc.edu) of the 17p12-p11.2 region (chr17:10,500,000-22,000,000; hg19). The different DISCRIMINATOR regions for the CMT1A/HNPP, PTLS/SMS, and Yuan-Harel-Lupski Primary CNV Intervals are shown as black bars, genes in this region are shown by dark blue bars, and the breakpoint regions are labeled and highlighted in blue and pink. (B) The number of Primary CNVs called within each cohort for each Jaccard threshold value (0-1-0.9) and Primary CNV (CMT1A, Potocki-Lupski, and Yuan-Harel-Lupski) for gain events. The true number of primary CNVs is depicted on the far right of each graph. CNVs identified in each cohort are color coded (Cohort #1: yellow, Cohort #2: blue, and Cohort #3: green). (C) The number of Primary CNVs called within each cohort for each Jaccard threshold value (0-1-0.9) and Primary CNV (Hereditary Neuropathy, Smith-Magenis, and Yuan-Harel-Lupski) for loss events. (D) The number of Primary CNVs called within each cohort for each Jaccard threshold value (0.3-0.5) for the Yuan-Harel-Lupski Microduplication. The number of Primary CNVs identified for remainder of the Primary CNVs in this region was unchanged across the 0.3-0.5 Jaccard threshold range.

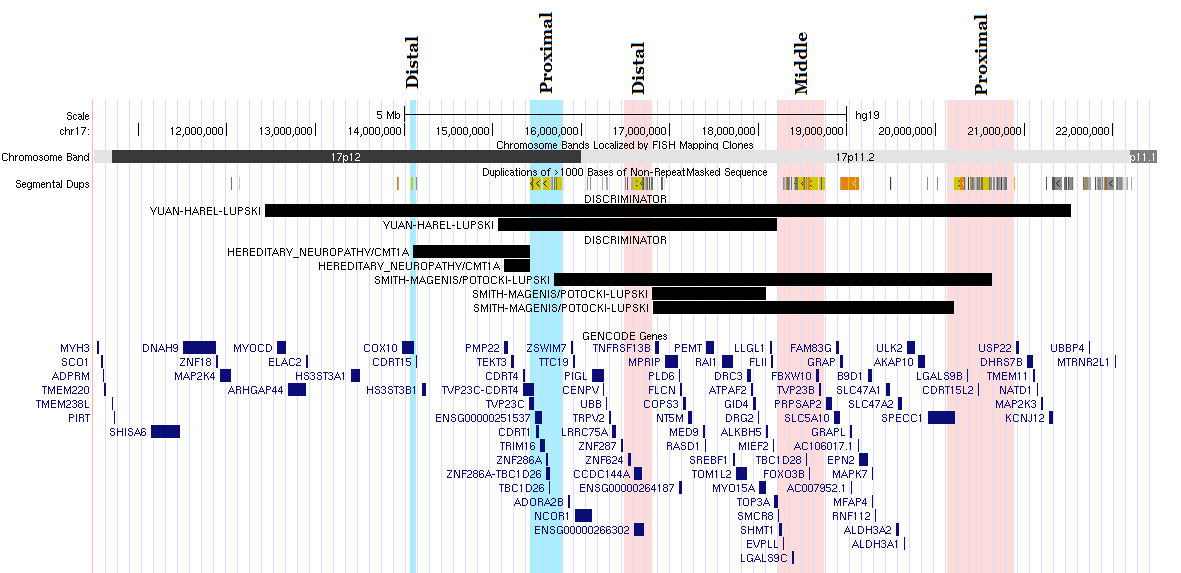

A

B

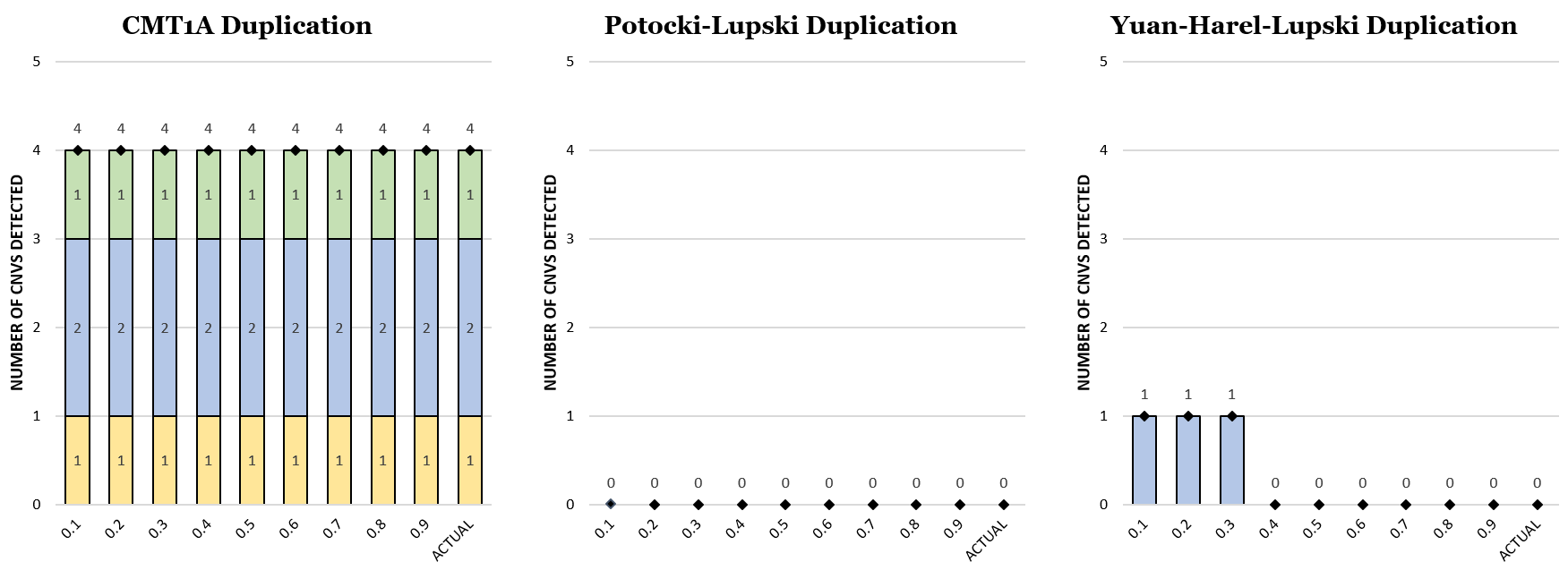

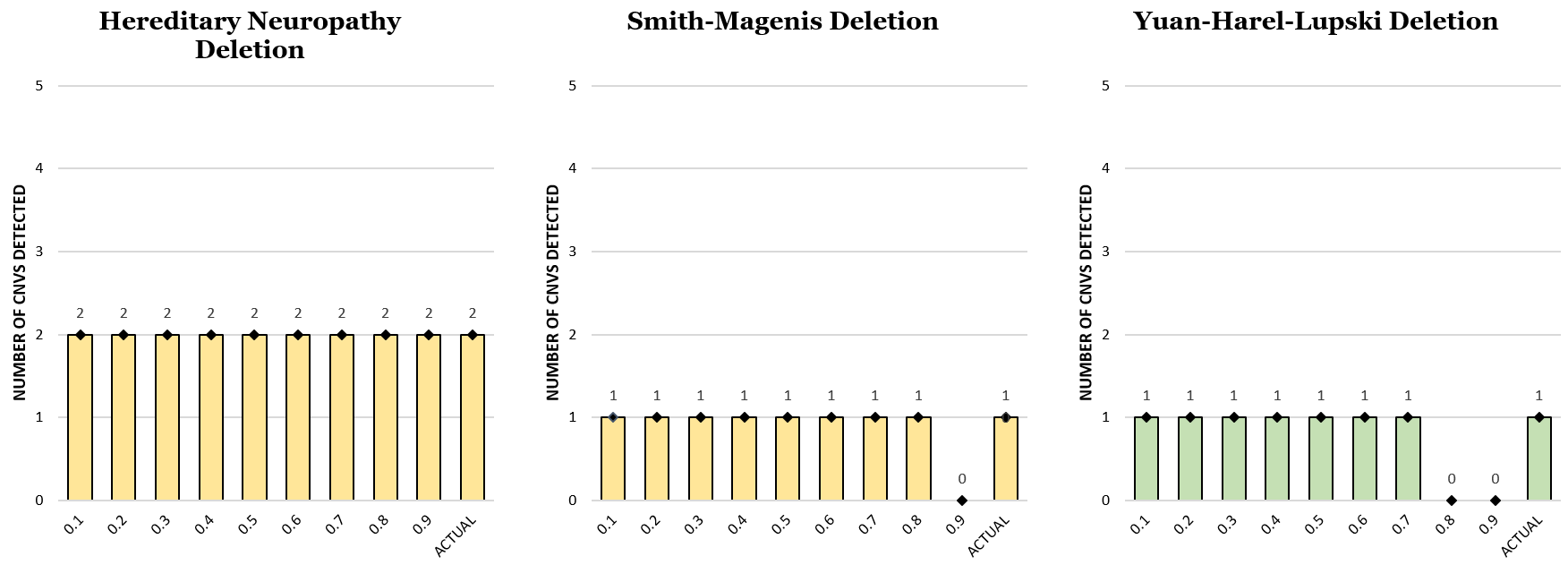

C

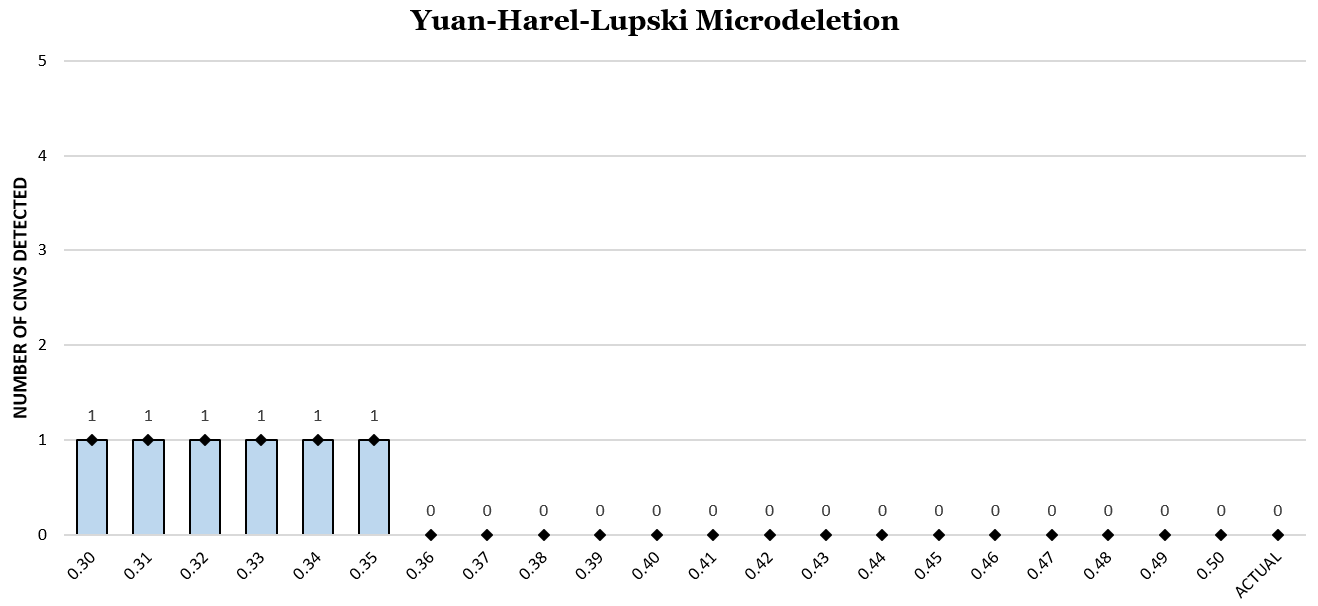

D

**Supplemental Figure 22: Number of Primary CNVs Identified by DISCRIMINATOR for Each Jaccard Value (****17q21.31)**

**.** (A) UCSC Genome Browser View (http://genome.ucsc.edu) of the 17q21.31 region (chr17:43,000,000-45,000,000; hg19). The DISCRIMINATOR region for the Koolen de Vries Primary CNV Interval is shown as a black bar, genes in this region are shown by dark blue bars, and the flanking segmental duplications are highlighted in blue. (B) The number of Primary CNVs called within each cohort for each Jaccard threshold value (0-1-0.9) and Primary CNV within the 17q21.31 locus for both gain and loss events. The number of Primary CNVs identified for the Koolen de Vries Deletion and Duplication was unchanged across the 0.3-0.5 Jaccard threshold range. The true number of primary CNVs is depicted on the far right of each graph. CNVs identified in each cohort are color coded (Cohort #1: yellow, Cohort #2: blue, and Cohort #3: green).

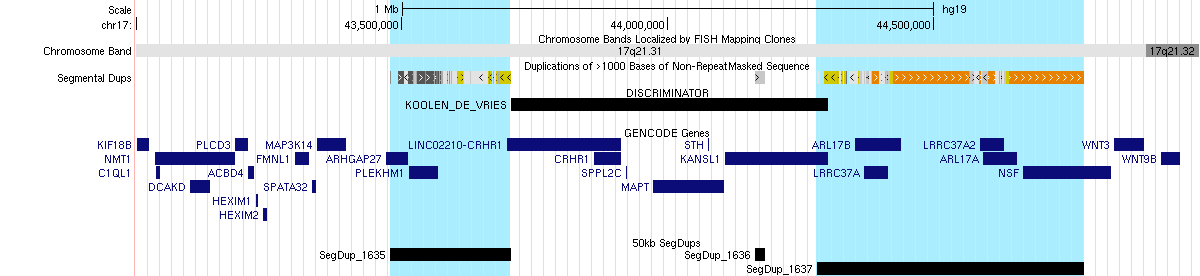

A

B

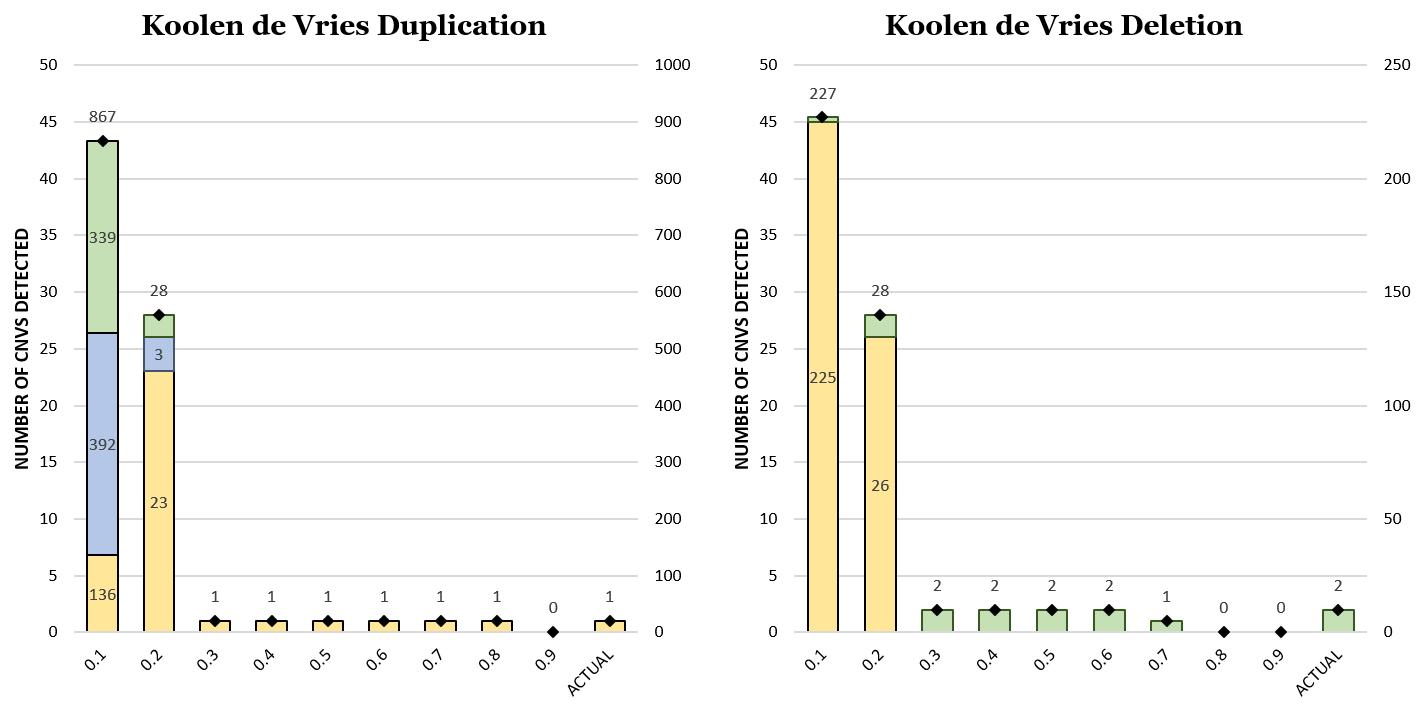

**Supplemental Figure 23: Number of Primary CNVs Identified by DISCRIMINATOR for Each Jaccard Value (22q11.2)**

**.** (A) UCSC Genome Browser View (http://genome.ucsc.edu) of the 22q11.2 region (chr22:18,000,000-26,000,000; hg19). The different DISCRIMINATOR regions for the 22q11.2 Primary CNV Intervals are shown as black bars, genes in this region are shown by dark blue bars, and the breakpoint regions are labeled and highlighted in blue. Note that while there are individual Primary CNV Intervals for the distal CNV regions, these are collapsed into the "DGS/VCFS Distal" Primary CNV. (B) The number of Primary CNVs called within each cohort for each Jaccard threshold value (0-1-0.9) and Primary CNV within the 22q11.2 locus for gain events. The true number of primary CNVs is depicted on the far right of each graph. CNVs identified in each cohort are color coded (Cohort #1: yellow, Cohort #2: blue, and Cohort #3: green). (C) The number of Primary CNVs called within each cohort for each Jaccard threshold value (0.3-0.5) for the 22q11.2 Distal Microdeletion. The number of Primary CNVs identified for remainder of the Primary CNV intervals in this region was unchanged across the 0.3-0.5 Jaccard threshold range.

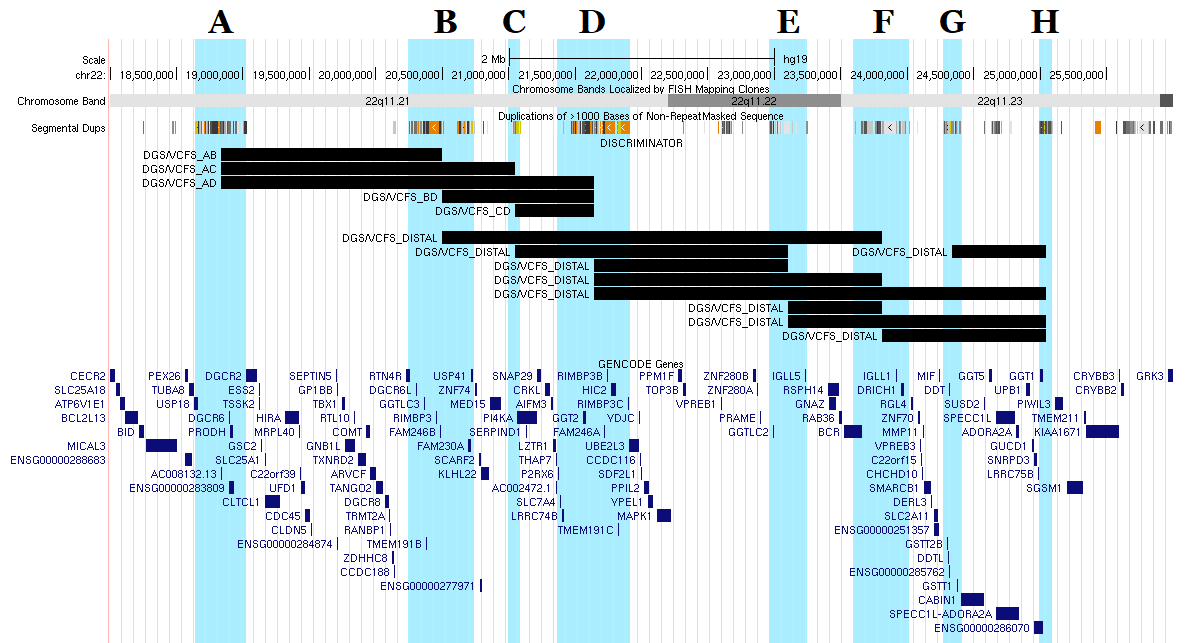

A

B

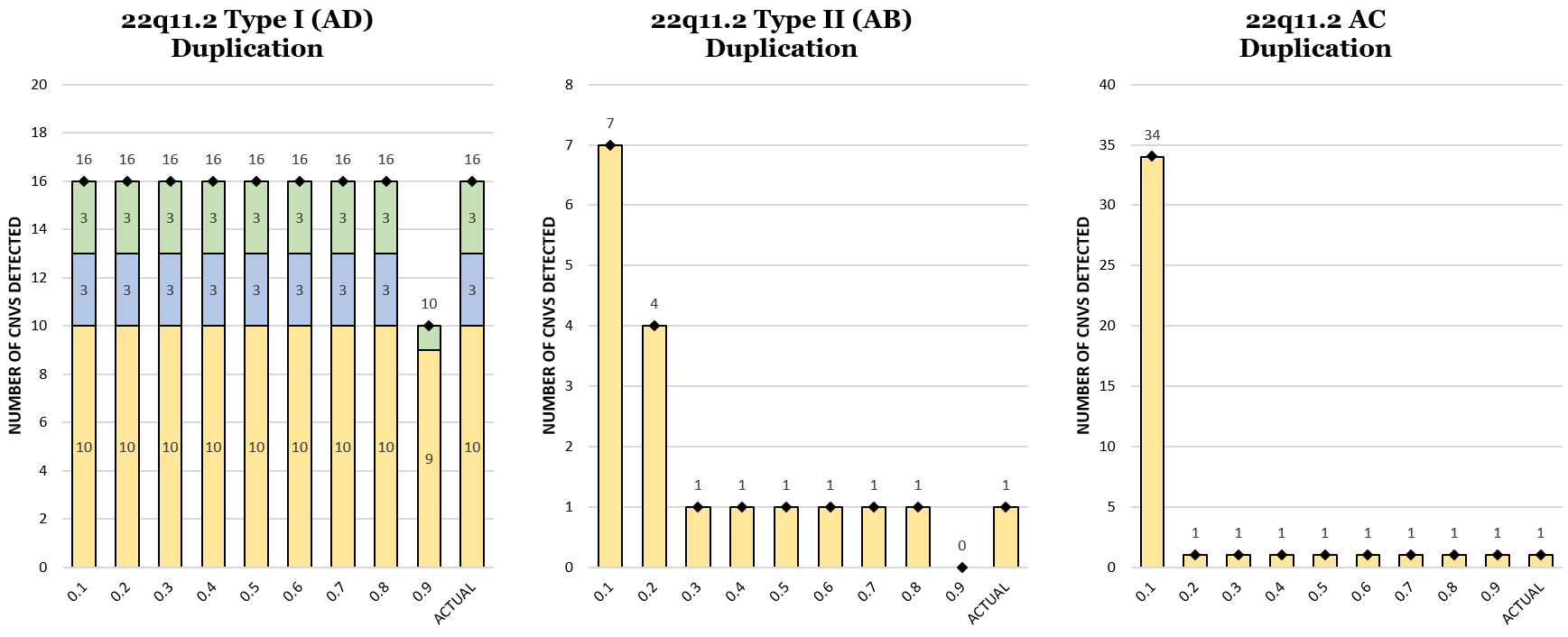

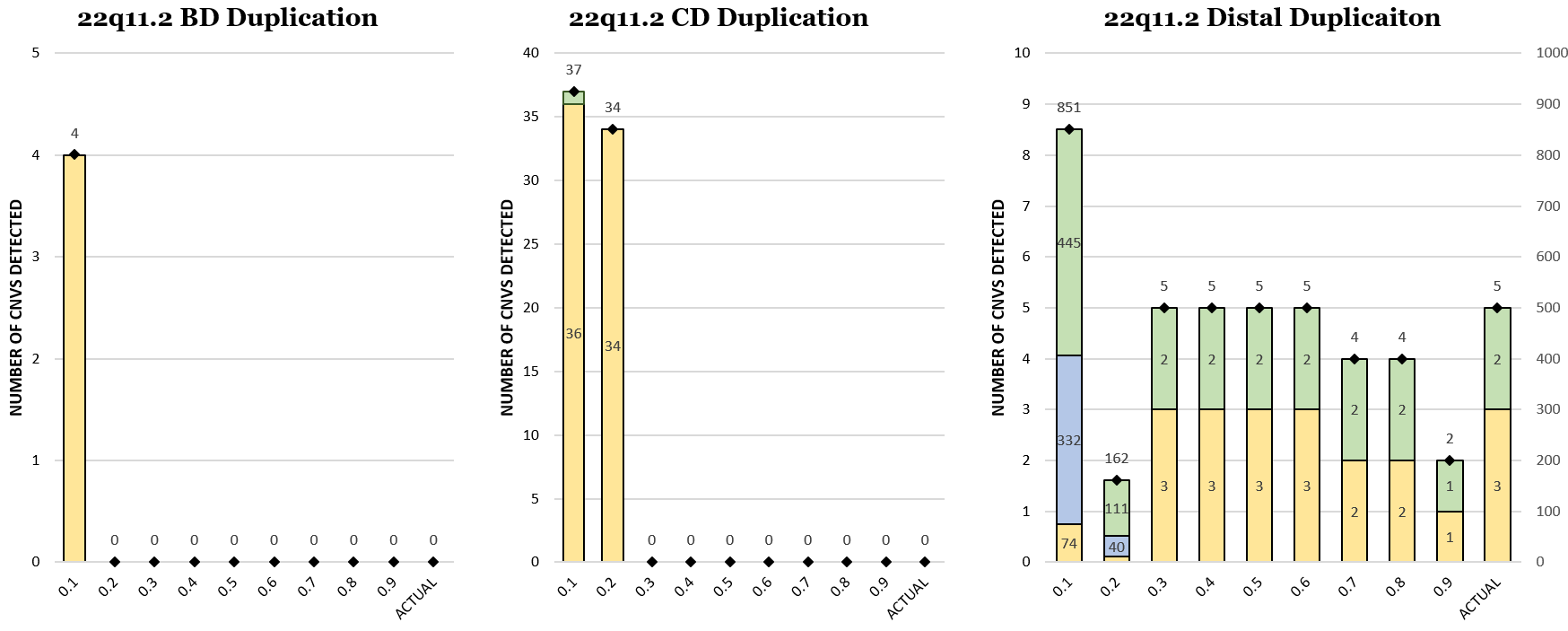

B

C

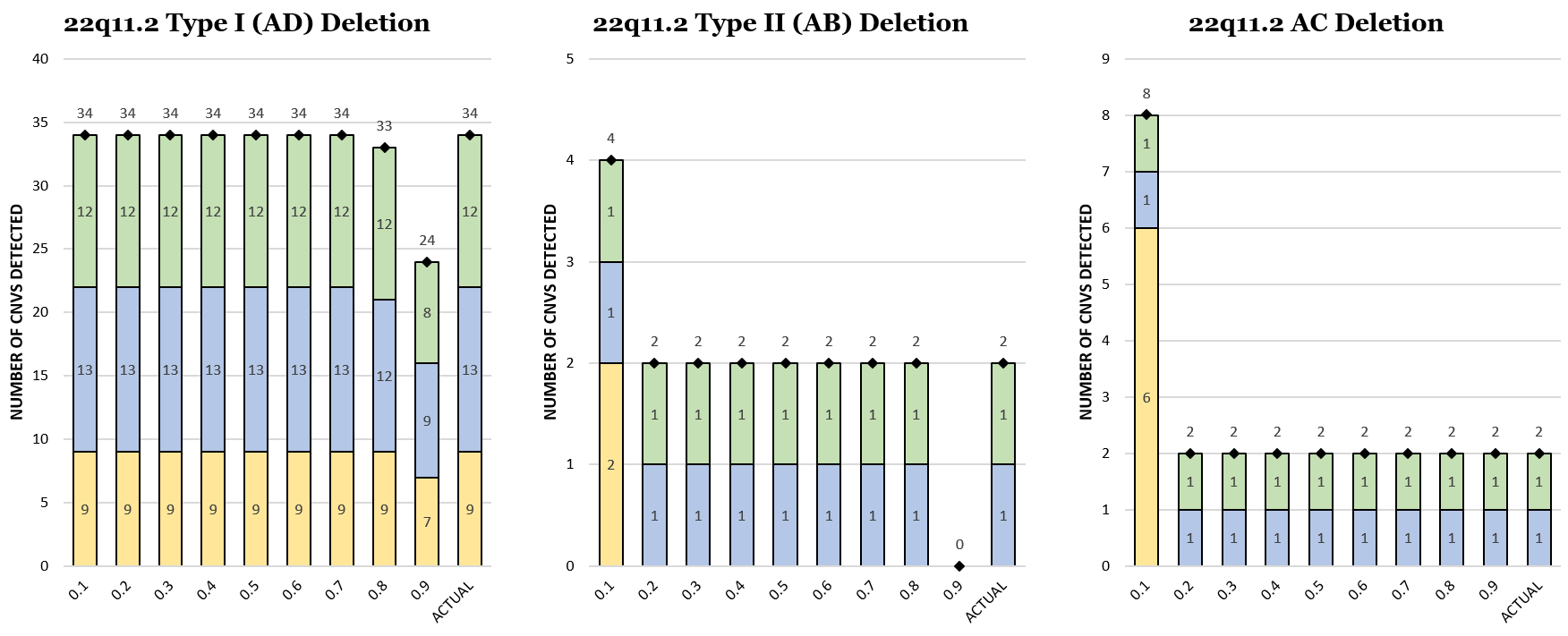

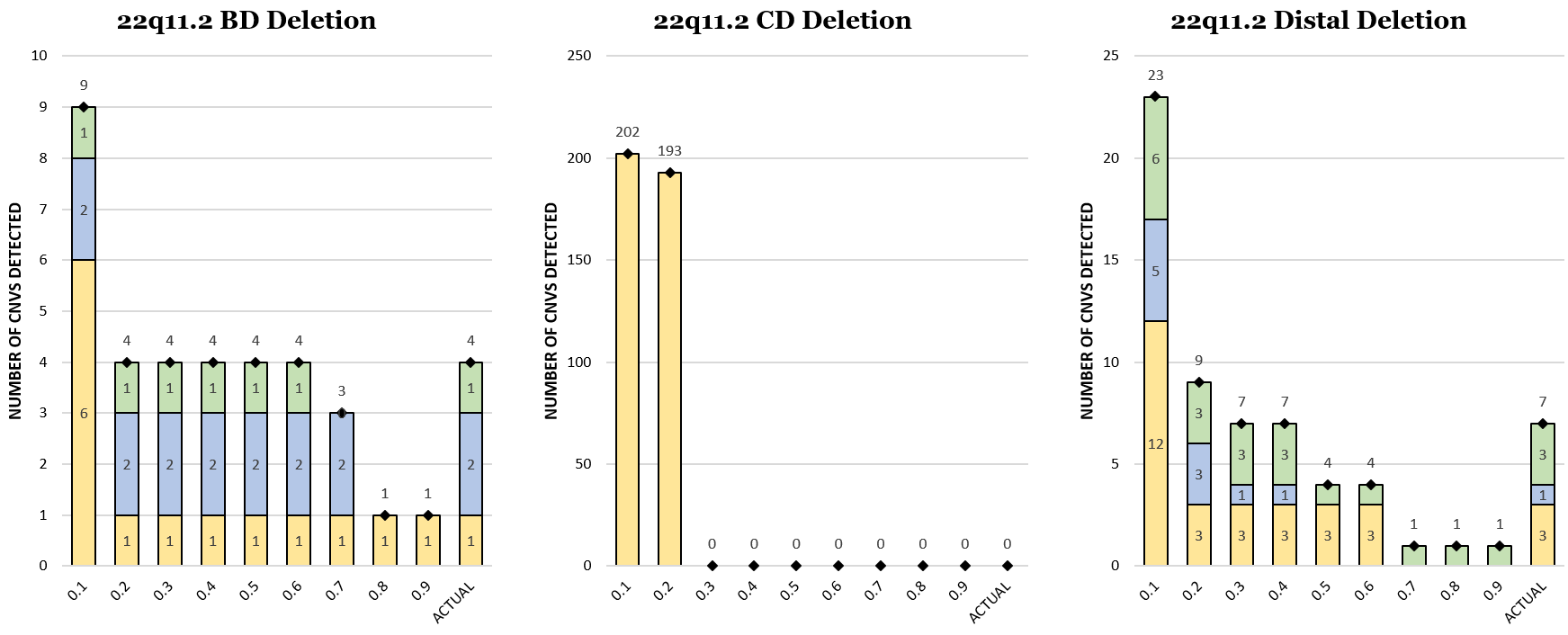

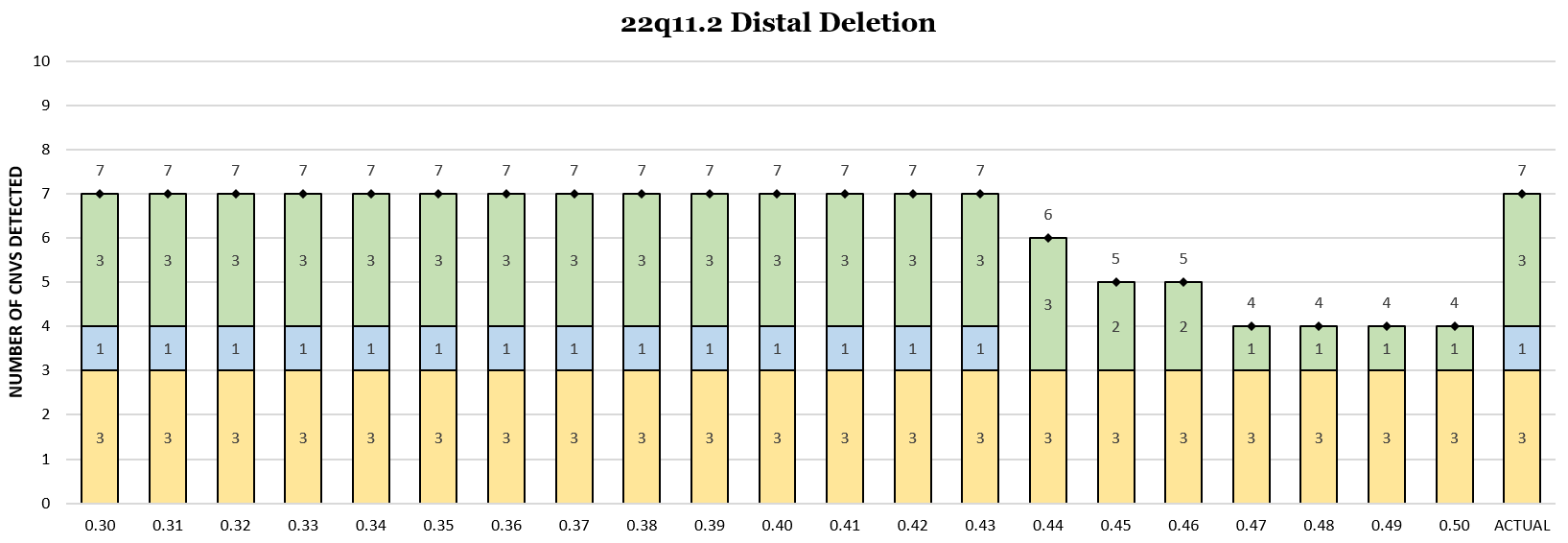

D

**Supplemental Figure 24: Number of Primary CNVs Identified by DISCRIMINATOR for Each Jaccard Value (Xp22.33)**

**.** (A) UCSC Genome Browser View (http://genome.ucsc.edu) of the Xp22.33 region (chrX:240,000-1,200,000; hg19). The different DISCRIMINATOR regions for the SHOX Primary CNV Intervals are shown as black bars and genes in this region are shown by dark blue bars. (B) The number of Primary CNVs called within each cohort for each Jaccard threshold value (0-1-0.9) and Primary CNV within the Xp22.33 locus for both gain and loss events. The true number of primary CNVs is depicted on the far right of each graph. CNVs identified in each cohort are color coded (Cohort #1: yellow, Cohort #2: blue, and Cohort #3: green). (C) The number of Primary CNVs called within each cohort for each Jaccard threshold value (0.3-0.5) for the SHOX Microduplication.

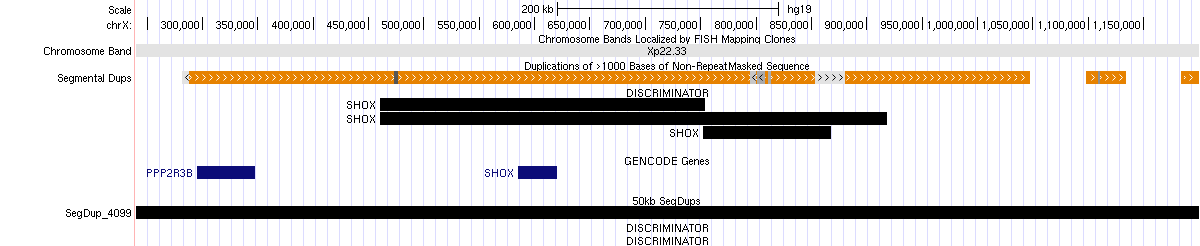

A

B

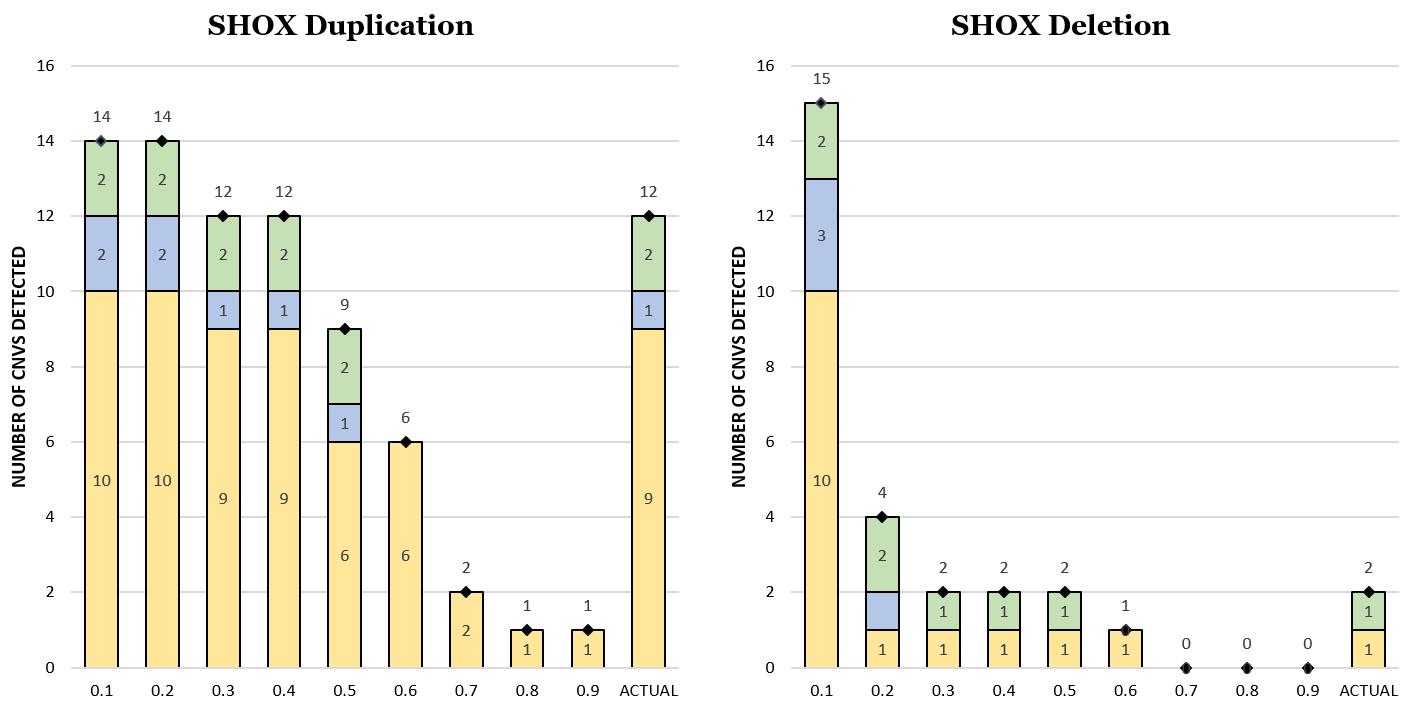

C

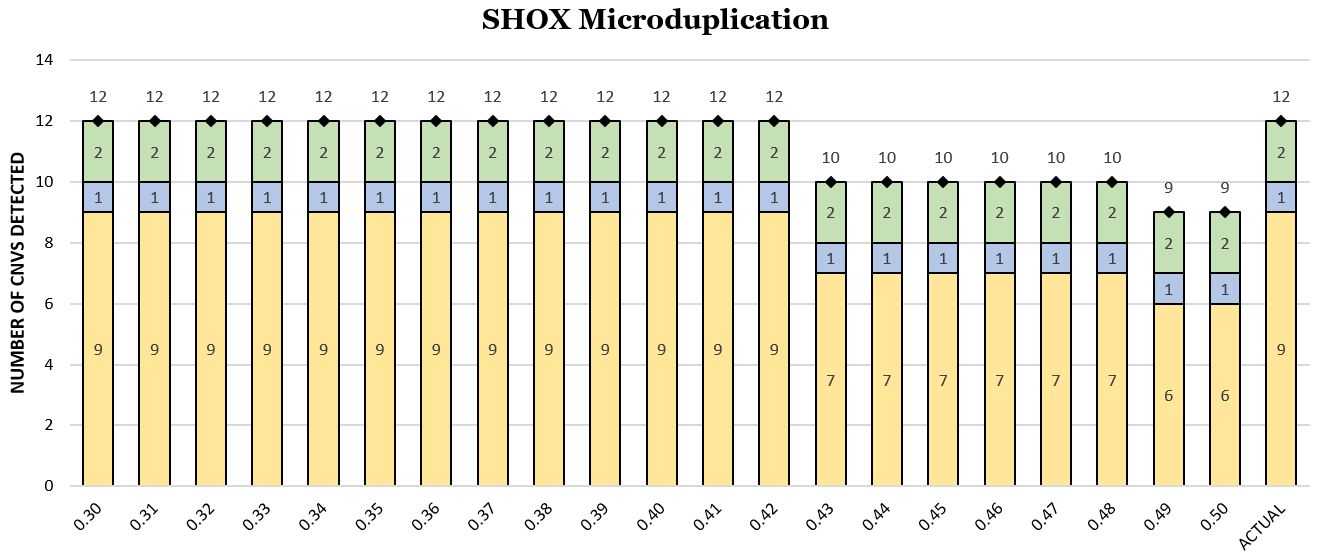

**Supplemental Figure 25: Number of Primary CNVs Identified by DISCRIMINATOR for Each Jaccard Value (Xp22.31)**

**.** (A) UCSC Genome Browser View (http://genome.ucsc.edu) of the Xp22.31 region (chrX:6,200,000-8,300,000; hg19). The different DISCRIMINATOR regions for the STS Primary CNV Intervals are shown as black bars and genes in this region are shown by dark blue bars. (B) The number of Primary CNVs called within each cohort for each Jaccard threshold value (0-1-0.9) and Primary CNV within the Xp22.31 locus for both gain and loss events. The true number of primary CNVs is depicted on the far right of each graph. CNVs identified in each cohort are color coded (Cohort #1: yellow, Cohort #2: blue, and Cohort #3: green). (C) The number of Primary CNVs called within each cohort for each Jaccard threshold value (0.3-0.5) for the STS Microduplication.

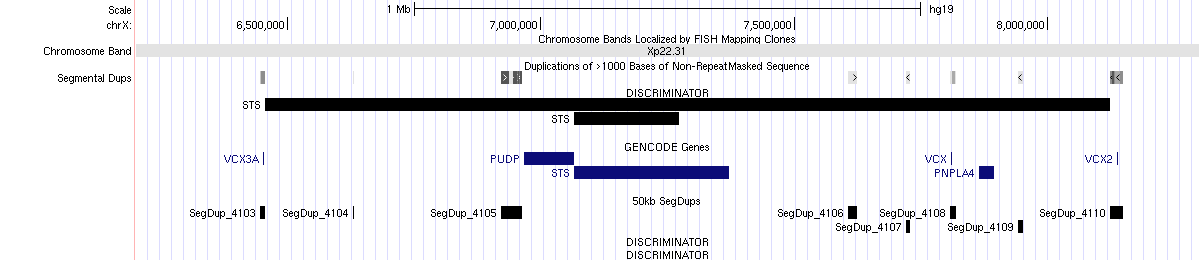

A

B

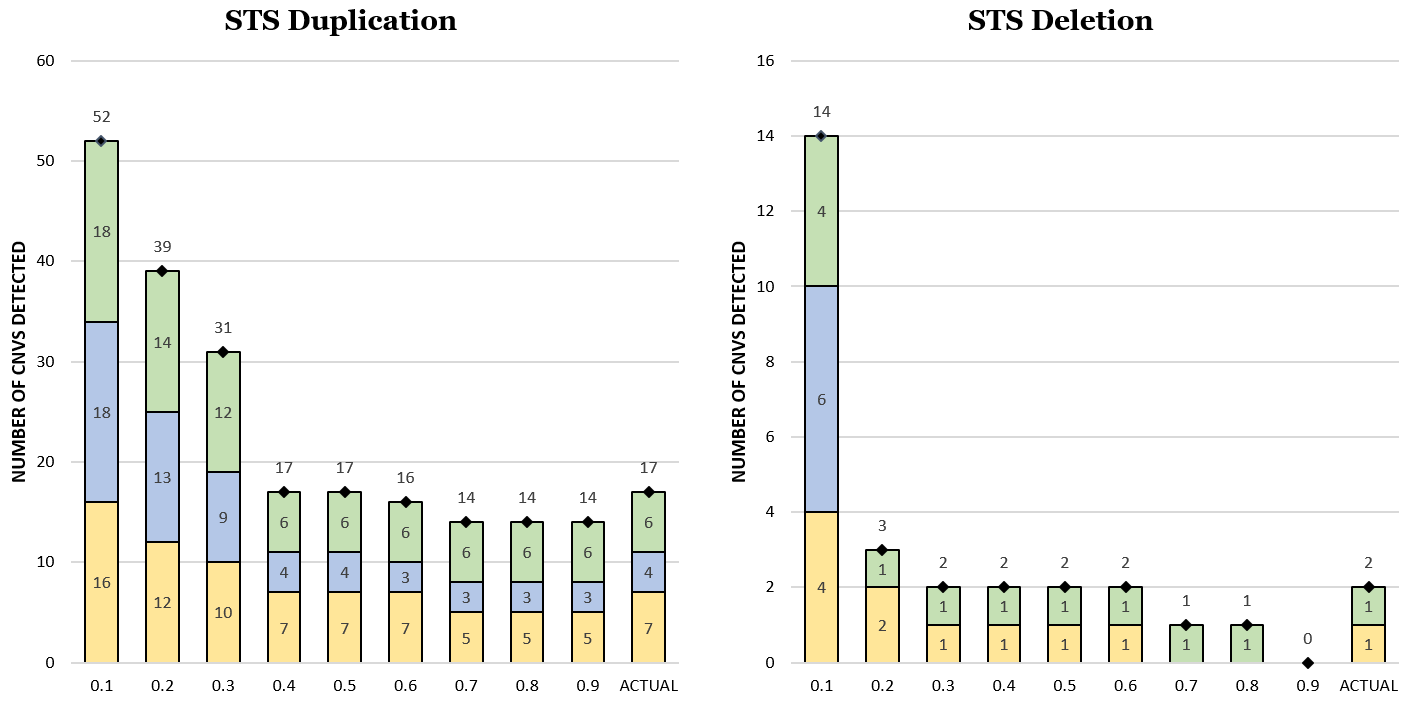

C

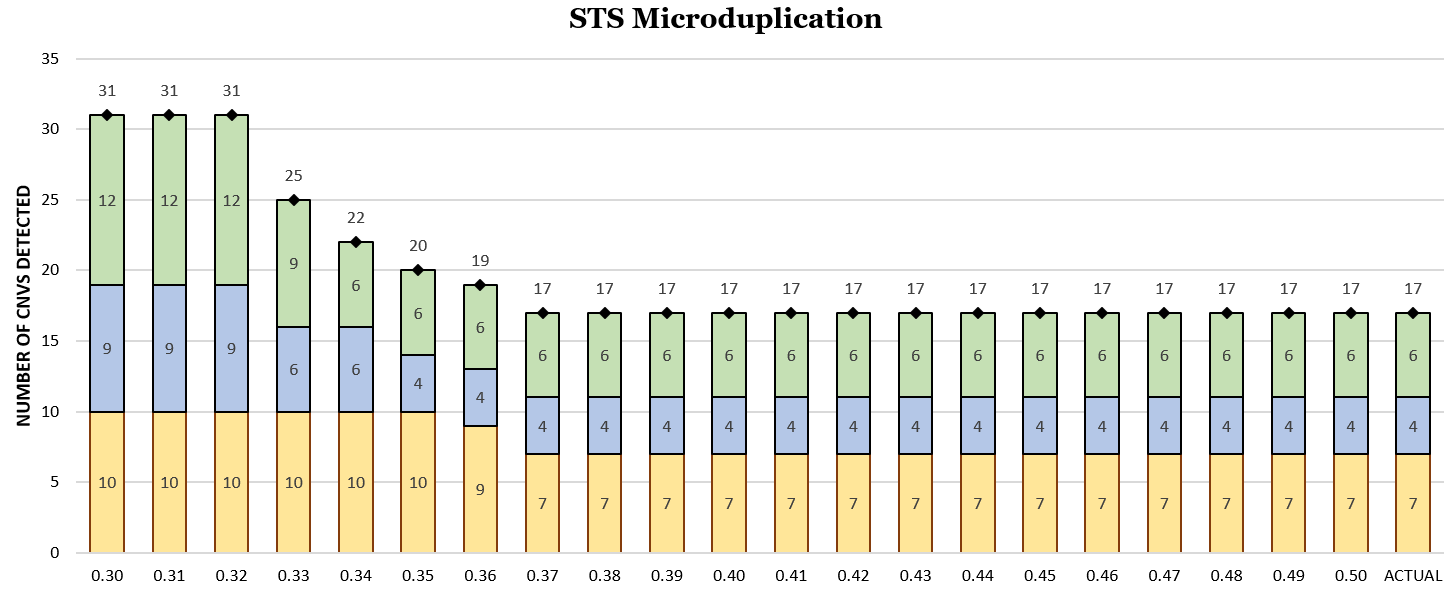

10.	Wetzel, A.S. & Darbro, B.W. A Comprehensive List of Human Microdeletion and Microduplication Syndromes. *BMC Genomic Data* (In Press).
